# Genome-wide association study and multi-ancestry meta-analysis identify common variants associated with carotid artery intima-media thickness

**DOI:** 10.1101/2025.04.11.25325582

**Authors:** Devendra Meena, Jian Huang, Marjan Zare, Natalie R. Hasbani, BOUA Palwendé Romuald, Rima Mustafa, Sander W van der Laan, Huichun Xu, James G. Terry, Joshua C. Bis, Deepti Jain, Nicholette D. Palmer, Nancy Heard-Costa, Yuan-I Min, Xiuqing Guo, Jie Yao, Kent D. Taylor, Jingyi Tan, Juan Peralta, Alexandre C. Pereira, Alyna Khan, Ananyo Choudhury, Anne B. Newman, Anny H. Xiang, Aroon Hingorani, Barry I. Freedman, Christopher J. O’Donnell, Claudia Giambartolomei, David M. Herrington, David R. Jacobs, Derek Klarin, Fei Fei Wang, Gerardo Heiss, HarshaVardhan Doddapaneni, Howard N. Hodis, Jai Broome, James G. Wilson, Jean-Tristan Brandenburg, John Blangero, Jose E Krieger, Josh D. Smith, Karine A. Viaud-Martinez, Kathleen A. Ryan, Leslie A. Lange, May E. Montasser, Michael C. Mahaney, Michal Mokry, Myriam Fornage, Patricia Munroe, Richard A. Gibbs, Russell P. Tracy, Ryan W. Kim, Scott M. Damrauer, Stephen S. Rich, Willa A. Hsueh, Yii-Der Ida Chen, NHLBI Trans-Omics for Precision Medicine (TOPMed) Consortium, The Million Veteran Program (MVP), TOPMed Atherosclerosis Working Group, Alanna C. Morrison, Braxton D. Mitchell, John Jeffrey Carr, Bruce M. Psaty, Donald W. Bowden, Ramachandran S. Vasan, Adolfo Correa, Wendy S. Post, Mark O. Goodarzi, Leslie J. Raffel, Joanne E. Curran, Michele Ramsay, Jerome I. Rotter, Paul Elliott, Nora Franceschini, Paul S. de Vries, Ioanna Tzoulaki, Abbas Dehghan

## Abstract

Carotid artery intima-media thickness (cIMT) is a measurement of subclinical atherosclerosis that predicts future cardiovascular events, including stroke and myocardial infarction. Genome-wide association studies (GWAS) have identified only a fraction of the genetic variants associated with cIMT. We performed the largest GWAS for cIMT involving up to 131,000 individuals. For the first time, we meta-analysed a diverse range of ancestries including populations with African, Asian (Chinese), Brazilian, European, and Hispanic ancestries. Our study identified 59 independent loci (53 loci from the multi-ancestry single variant analysis of which 19 are novel, P<5x10^-8^; 6 novel in gene-based analysis from single variant analysis, P=2.6x10^-6^, 2 novel in meta-regression) associated with cIMT. Gene-based, tissue-expression and gene-set enrichment analyses revealed novel genes of potential interest and highlighted significant relationships between vascular tissues (aorta, coronary and tibial arteries) and genetic associations. We found that circulatory levels of seven proteins, including ACAN, BCAM, DUT, ERI1, APOE, FN1, and GLRX were potentially causally associated with cIMT levels. We found a strong genome-wide correlation between cIMT and various cardiometabolic, smoking phenotypes, and lipid traits. Using Mendelian randomisation, our analyses provide robust evidence for causal associations between cIMT and several clinically relevant traits, including lipids, blood pressure, and waist circumference. Our study extends our genetic knowledge of atherosclerosis and highlights potential causal relations between risk factors, atherosclerosis and clinical diagnoses.

## Introduction

Carotid artery intima-media thickness (cIMT), a measurement of subclinical atherosclerosis, is correlated with change in coronary artery disease assessed by quantitative coronary angiography^1^ and is a strong predictor of future clinical cardiovascular events^2^ that can be measured non-invasively by B-mode ultrasound imaging of the carotid arteries^3^. cIMT is significantly heritable with study-specific heritability estimates ranging from 35-60%, suggesting a substantial genetic component underlying individual differences in cIMT measurements^4–7^.

To date, genome-wide association studies (GWAS) have identified at least 35 loci robustly associated with cIMT (P<5×10^−8^)^8–14^. The largest GWAS meta-analysis of cIMT to date (comprising up to 100,253 individuals of European ancestry) came from a meta-analysis combining data from the UK Biobank (UKBB), the Cohorts for Heart and Aging Research in Genomic Epidemiology (CHARGE), and the University College London-Edinburgh-Bristol (UCLEB) consortia. However, most previous GWAS of cIMT focused on individuals of European ancestry only.

Here, we conduct the first multi-ancestry GWAS meta-analysis of cIMT that includes individuals of European, African, Hispanic, Asian (Chinese), and Brazilian populations from the UKBB, CHARGE-UCLEB consortium^9^; Trans-Omics for Precision Medicine (TOPMed) Program; the Africa Wits-INDEPTH partnership for Genomic Studies (AWI-Gen), the Mexican-American Coronary Artery Disease (MACAD); the Hypertension-Insulin Resistance Family Study (HTN-IR); and the Baependi Heart Study (BHS).

## Results

A European- and multi-ancestry GWAS meta-analysis of cIMT was conducted, aggregating data from seven consortia/studies (**Supplementary** Fig. 1) that identified 51 loci, applying stringent quality control filters before meta-analysis. The genomic inflation factor showed little evidence for population stratification (λGC range 0.95–1.09) (**ST 1–2**).

### European- and multi-ancestry GWAS meta-analysis

A European-only meta-analysis was first performed, combining UKBB (IMT mean maximum) with CHARGE-UCLEB cIMT GWAS summary statistics and identified 39 loci at P<5×10^−8^ (**ST 3**). A subsequent fixed-effects inverse variance-weighted multi-ancestry meta-analysis included non-overlapping samples from UKBB and CHARGE-UCLEB, TOPMed, AWI-Gen, MACAD, HTN-IR, and BHS (**Supplementary** Figs. 2-3), identifying an additional twelve loci, taking the total to 51, of which 17 were novel (**Fig. 1, ST 4**). A meta-regression was applied using MR-MEGA^15^ to account for the heterogeneity in allelic effects between the ancestral groups, identifying two additional loci on chromosome 7 near *CPVL* (rs1522919, PMR- MEGA=2.5×10^−8^) and chromosome 15 near *TMC3/STARD5 (*rs75789774, PMR-MEGA=3×10^−8^). We replicated all 35 known signals from published GWAS at a p-value of 0.05 (**ST4a**).

**Figure 1.**
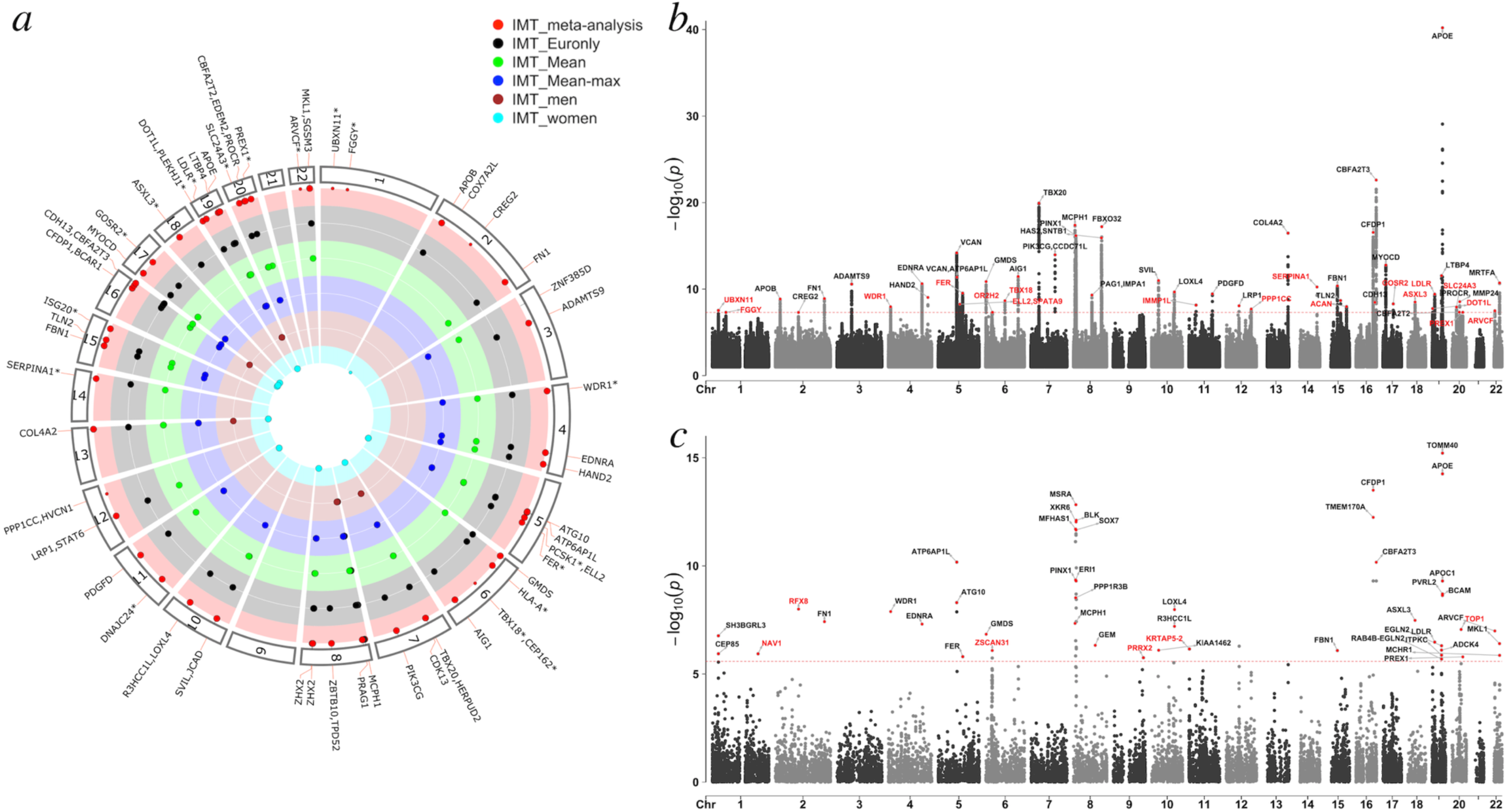
Manhattan and Fuji plots of common variants associated with cIMT. **(a)** Fuji plot showing independent loci associated with cIMT. The colour-coded circles (from inside to outside) represent GWAS of 1) IMTmean-max GWAS in women in UKBB; 2) IMTmean-max GWAS in men in UKBB; 3) IMTmean in UKBB; 4) IMT mean-max in UKBB and 5) IMTmean- max multi-ancestry GWAS meta-analysis. Every dot represents a locus associated with cIMT; large dots represent cross-trait loci, while small dots represent trait-specific loci. Genes with an asterisk denote novel cIMT loci; **(b)** Manhattan plot for the multi-ancestry GWAS on cIMT showing the negative log10 transformed P value for each SNP on the y axis and the base-pair position of the SNPs on each chromosome on the x-axis. The genome-wide-significance threshold (P<5×10^−8^) is represented as the horizontal red line. The 51 cIMT loci are annotated; red and black dots refer to novel and known loci, respectively; and **(c)** Gene-based analysis: Manhattan plot showing the genes associated with cIMT in the gene-based analysis. The y- axis shows the negative log10 transformed P value of the gene-based test computed in MAGMA, and the x-axis shows the genomic position on each chromosome. The red line indicates the Bonferroni-corrected threshold for genome-wide significance (P=2.6x10^-6^ (0.05/19,220 genes). The novel signals from the gene-based test are highlighted in red.

In a conditional analysis at the 51 loci, we found further independent associations at four loci including *SVIL* (rs2369339, rs9337951), *MYOCD* (rs6502187, rs7504018), *COL4A2* (rs9515203, rs4773141), and *APOE* (rs7412, rs429358) (**ST4**). We have generated regional plots (**Supplementary** Figure 4), forest plots for the most significant SNPs at each locus (**Supplementary** Figure 5) and Circos plots to show eQTL and chromatin interaction in the identified loci (**Supplementary** Figure 6).

### Sex-specific GWAS in the UKBB

A sex-specific cIMT GWAS was conducted in the UKBB to detect sex-related genetic differences. The analysis identified 17 loci (11 for women and 6 for men; **Supplementary** Fig 7, **ST 5-6**), of which three were not detected in the main GWAS (**Supplementary** Fig. 7, **ST 7-8**). Two loci were significant only in women including rs4953207 (β=0.021, P=4×10^−8^, near *COX7A2L*, p for interaction = 0.03) and rs746610273 (β=0.027, P=3.7×10^−8^ *near CDK13*, p for interaction = 0.01). The effect estimates and p-values from the UKBB sex-specific GWAS showed a moderate correlation (r=.78) (**Supplementary** Fig 8).

### Functional annotation and gene mapping of cIMT loci

Functional annotation of genome-wide significant single nucleotide polymorphisms (SNPs) using FUMA showed that the vast majority (∼84%) of these lies within intronic or intergenic regions (**ST 9**). The lead SNPs at *APOE* (rs7412) and *LTBP4* (rs34093919) on chromosome 19 were exonic, non-synonymous and had the highest observed probability of a deleterious protein effect (Combined Annotation Dependent Depletion (CADD) score >25), indicating a probable deleterious impact of these variants on gene function. The remaining 10 exonic non-synonymous SNPs at *FN1*, *MCPH1*, *PPP1CC*, *SERPINA1*, *CBFA2T3* (rs12930980 and rs12924185), *APOE* (rs28399653, rs28399654, rs429358) and *MKL1* were in high linkage disequilibrium (LD) with either a lead SNP or with one of the independent significant SNPs (r^2^>0.6). Out of 246 genes that were mapped to independent significant SNPs or their proxies (r^2^ >0.6), gene mapping prioritised 106 genes based on *genomic position*, 47 based on expression quantitative loci (*eQTLs*), and 176 based on *3D chromatin interaction* mapping (**Fig. 2d**, **ST 10**).

**Figure 2.**
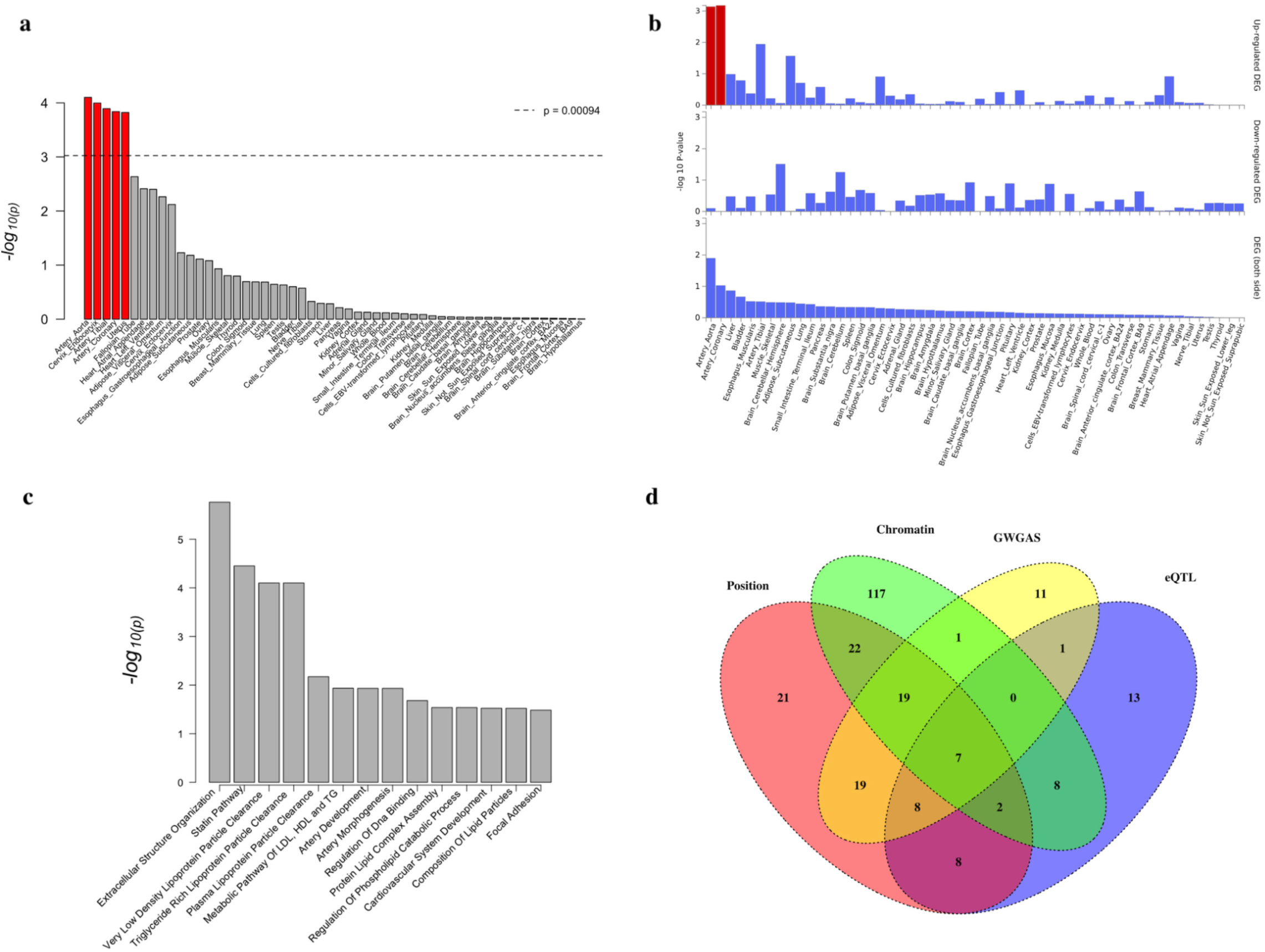
Functional annotation of cIMT-associated variants. **a)** MAGMA tissue expression analysis using gene expression per tissue based on GTEx version 8 data for 53 specific tissue types. The horizontal dotted line indicates the Bonferroni-corrected significance threshold (P = 0.05/53; -log10 transformed). Significant tissues are shown in red. **(b)** Differential expression gene analysis of prioritized genes across GTEx version 8 30 general tissue types. Genes with P-value ≤ 0.05 after Bonferroni correction and absolute log fold change ≥ 0.58 were defined as differentially expressed genes in a given tissue compared to others. Significantly enriched DEG (Bonferroni corrected P-value ≤ 0.05) are highlighted in red. **(c)** Gene-set and pathway enrichment analysis; and **(d)** Venn diagram showing overlap of genes implicated by positional mapping, chromatin interactions, GWGAS, and eQTL mapping. GWGAS, genome-wide gene-based association study; eQTL, Expression quantitative trait loci.

### Statistical fine-mapping (Credible-set analysis)

Bayesian fine-mapping was performed using summary statistics from the European-only GWAS and multi-ancestry meta-analysis to identify potential causal variants at the identified loci. At 3 loci (*COL4A2*, *LTBP4, APOE*), a single variant was highlighted as the likely single causal SNP (**ST 11**). For 22 out of 39 loci, the multi-ancestry meta-analysis results yielded smaller 95% credible sets compared to the European-only (**ST 11a**, **Supplementary** Fig. 9), with the highest reduction in the number of variants at *ZBTB10*, *ZHX2*, and *CFDP1* loci.

### Genome-wide gene-based association analysis

A genome-wide gene-based association study (GWGAS) was conducted on the summary statistics from a multi-ancestry meta-analysis using MAGMA in FUMA. This identified 66 protein-coding genes significantly associated with cIMT (Bonferroni-corrected P-value < 2.6x10^-6^ (0.05/19,220 genes)), sixty of which were also included in the 246 prioritised genes mapped to 51 loci in the single variant analysis (**Fig. 1c**, **ST12**). The most significant association was with *TOMM40* (P=6.1×10^-16^) followed by *APOE* (P=5.5×10^-15^) and *CFDP1* (P=3.1× 10^-14^).

### Rare variant analysis for in TOPMed

Gene-based rare variant analyses on whole genome sequence data from 24,953 participants across nine TOPMed studies (five ancestry groups) using Efficient Variant-Set Mixed Model Association Test (SMMAT) revealed 56 significant genes (Supplementary document) with 10 genes showing nominal significance (P<0.05), and MSRA consistently meeting significance across all aggregation strategies (Supplementary document, **ST 12a**).

### Gene-set enrichment and Tissue expression (gene-property) analysis

Gene-set enrichment and tissue expression analyses were conducted on 246 prioritised genes in sets of genes defined by MsigDB v7.0, WikiPathways, and the NHGRI-EBI GWAS catalog to link multi-ancestry GWAS findings to biological functions. The strongest enrichment was found for NHGRI-EBI GWAS catalog-reported genes for a general factor of neuroticism (P=5.1×10^-18^) followed by cIMT (P=3.4×10^-13^) (**Fig. 2c**, **ST13**).

Differentially expressed gene (DEG) analysis across 30 tissue types showed a significant upregulation of these genes in blood vessel tissues, including coronary and aorta arteries (P<0.03) (**Fig. 2b**, **ST13a**). Tissue expression analysis found that the genes identified by gene-based analysis were significantly enriched for expression in blood vessel tissues: tibial artery (P=1.3×10^-3^); coronary artery (P=1.4×10^-3^); and aorta artery (P=7.9×10^-5^) (**Fig. 2a**, **ST13b**).

### Genetic correlation and heritability estimate of cIMT-associated variants

The SNP-based heritability (h^2^) of the European-only cIMT-associated variants was 0.1095 (se=0.001). A genome-wide genetic correlation (rg) was estimated between the European-only IMT GWAS and 53 other complex trait/disease categories using LD score regression (http://ldsc.broadinstitute.org/ldhub/). After adjusting for multiple testing (P=0.00094 (0.05/53)), a significant genetic correlation was found with ten traits/diseases (**ST14**) six of which had rg values stronger than 0.2, including type 2 diabetes (T2D; rg=0.29), waist circumference (WC; rg=0.24), overweight (rg=0.23), waist-to-hip ratio (rg=0.22), obesity (rg=0.22) and body mass index (BMI; rg=0.21).

### Association with clinical outcomes

Lookups of all independent SNPs (r^2^<0.6, n=399) in publicly available GWAS summary statistics of various diseases and cardiovascular risk factors relevant to cIMT were performed. We found that 41 loci were related to at least one of the 15 traits and disease categories, including general factors of neuroticism, coronary artery disease (CAD), immune-, blood- and lipid-related traits (**Fig. 3a**, **ST15**). Blood pressure (BP; systolic blood pressure (SBP) and diastolic blood pressure (DBP)) had the largest number of overlapping genes with 28 genes associated with both BP and cIMT. We also identified 10 loci for CAD, 8 loci for carotid plaque, 4 for lipid levels and peripheral artery disease (PAD), 2 for ischemic stroke (AIS), 2 for any stroke (AS), 1 for arterial stiffness index (ASI), and 1 for Alzheimer’s disease (AD) (**Figs. 3b-3c**).

**Figure 3.**
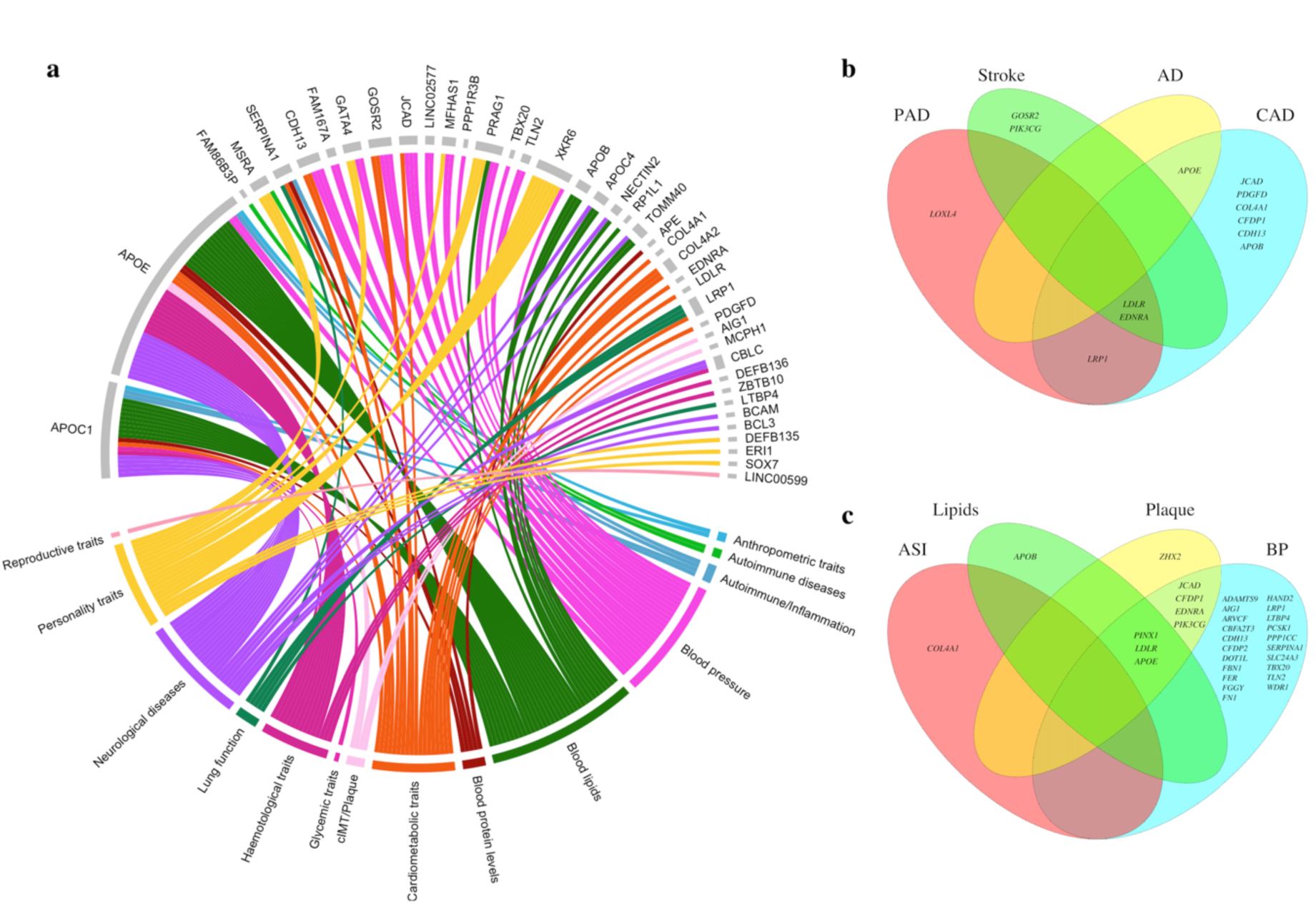
Association of the identified cIMT SNPs with clinical traits and diseases. **(a)** Look up in GWAS catalog **(b) Look up in published GWAS on cardiovascular** diseases, and **(c) Look up in published GWAS on cardiovascular** traits. PAD, peripheral arterial disease; AD, Alzheimer’s disease; CAD, coronary artery disease; Plaque, carotid plaque; ASI, arterial stiffness index; BP, blood pressure.

### Mendelian randomisation

A bi-directional MR analysis was conducted on cIMT and various clinically relevant traits and diseases (**ST16**). We found supporting evidence for a positive effect of low-density lipoprotein cholesterol (LDL; βIVW =0.013, P=7.4×10^-10^), BMI (βIVW=0.018, P=8.2×10^-6^), waist circumference (βIVW =0.023, P=6.2×10^-7^), SBP (βIVW =0.002, P=9.1×10^-41^), DBP (βIVW =0.001, P=2.1×10^-13^), pulse pressure (PP; βIVW =0.0046, P=2.0×10^-74^), and life-time smoking (βIVW=0.026, P=4.1×10^-4^) and an inverse effect of high-density lipoprotein cholesterol (HDL; βIVW =-0.007, P=0.001) on cIMT (**Fig. 4a**, **ST17**). MR-Egger did not suggest a directional pleiotropic effect.

**Figure 4.**
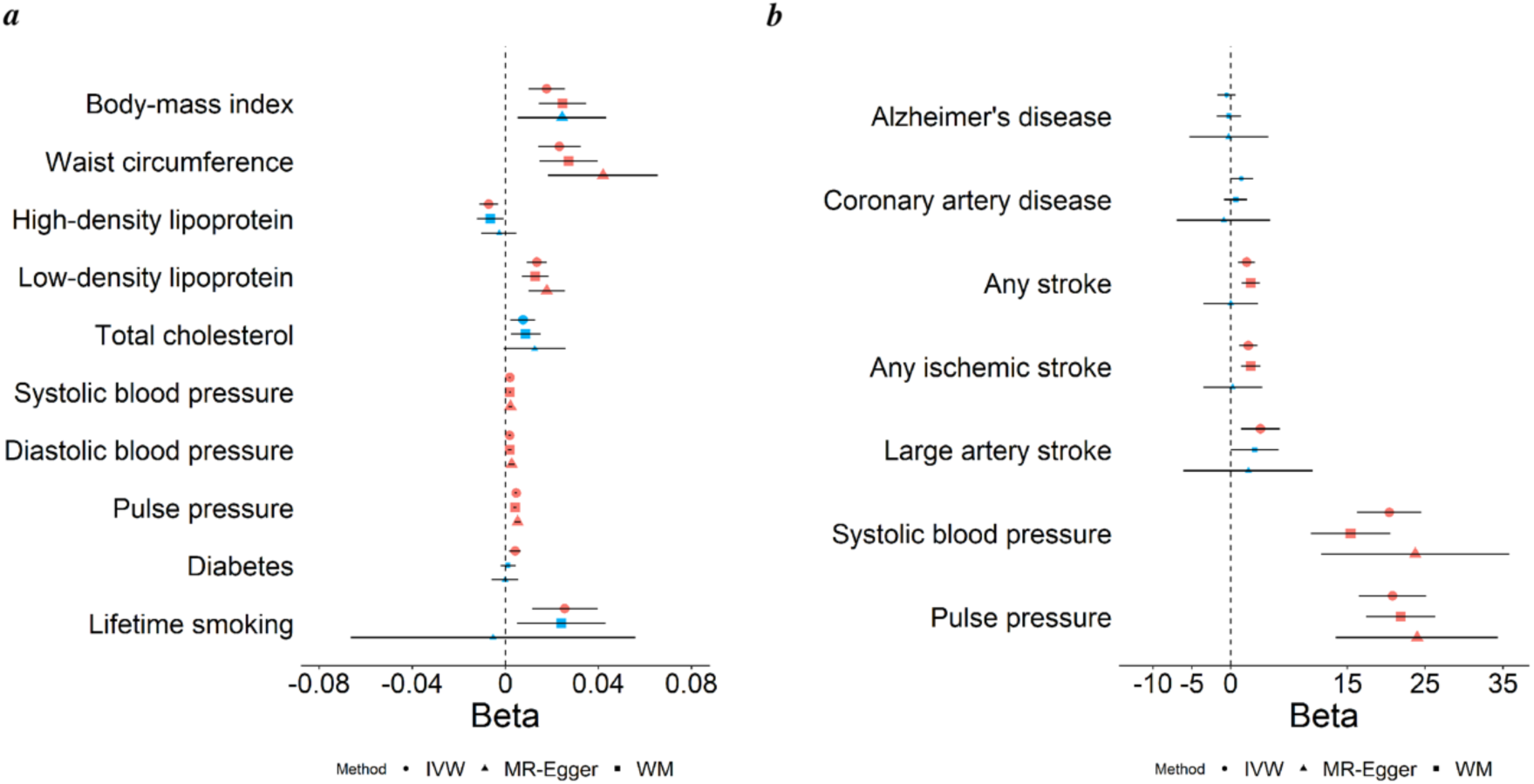
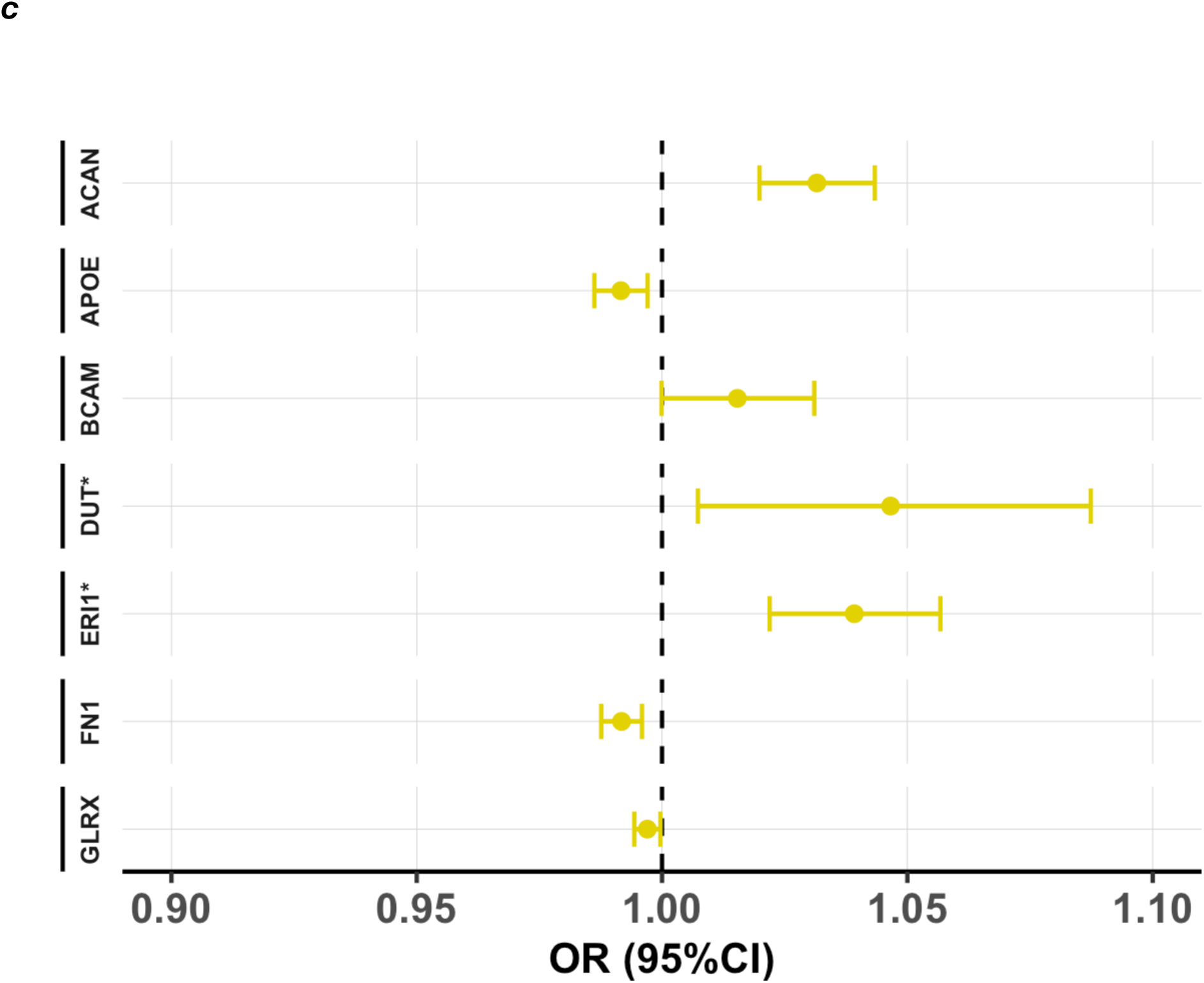
Forest plot of the IVW, WM and MR-Egger estimates. **(a)** Effect of cardiovascular risk factors on cIMT, **(b)** Effect of cIMT on cardiovascular traits and diseases and c) Forest plot for the effect estimates of selected plasma proteins on cIMT. *Represents proteins with Wald ratio (causal estimate obtained for a single genetic variant). MR, Mendelian randomization; IVW, inverse-variance weighted; WM, weighted median.

Conversely, cIMT was significantly associated with SBP (βIVW =20.4, P=3.2×10^-22^), PP (βIVW =20.8, P=5.4×10^-21^), risk of any stroke (ORIVW=7.9, P=2.3×10^-4^), ischemic stroke (ORIVW =9.6, P=2.0×10^-4^), and large artery stroke (LAS; ORIVW=46.1, P=0.003) (**Fig. 4b**, **ST18**) with support from at least one robust method to rule out pleiotropic effects.

Given the bidirectional association observed between SBP and cIMT, a sensitivity analysis was performed excluding shared genetic instruments for cIMT and SBP, showing a consistent association between SBP and cIMT supported by WM, and MR-Egger (βIVW=0.0020, P=1.1×10^-40^) (**ST19**). The reverse direction showed a slight decrease in magnitude (βIVW=15.1, P=1.9×10^-14^), without supporting evidence by MR-Egger and potential evidence of horizontal pleiotropy (P=0.042) (**ST19**). The MR-Steiger estimates suggested a bidirectional relationship between cIMT and SBP with a stronger causal effect of SBP on cIMT (p=3.7×10^-271^) compared to the effect of cIMT on SBP (p=1.5×10^-138^) (**ST19a**).

We used data from the UK Biobank in a framework of Mendelian randomisation (MR) analysis to examine whether the protein products of the identified genes are causally related to cIMT. Of the 246 genes prioritised by FUMA, 39 genes were found to be coding for a protein that was measured by Olink assay and were carried forward for performing cis-MR. Our cis-MR revealed that genetically predicted ACAN, BCAM, DUT, and ERI1 levels showed a positive association while APOE, FN1, and GLRX showed a negative association with cIMT levels (**ST20; Fig. 4c)**

### cIMT loci associate with cell-type specific plaque expression and morphology

We assessed the cell-type specific expression of the 54 nearest protein-coding genes at each of 51 loci in atherosclerotic plaques using the single-cell RNA sequencing data from the Athero-Express Biobank study^16^ that included 35 patients undergoing carotid endarterectomy^17^. Of note was the cell-type-specific expression of the arterial wall: *FN1*, *LRP1*, *COL4A2*, *FBN1*, *CDH13*, *LTBP4* in ACTA2+ smooth muscle cells, and *SOX7*, *JCAD*, *PDGFD*, *CDH13*, *LTBP4*, *COL4A2* in endothelial cells. We also observed inflammatory cell-specific expression of *TBD52*, *CBFA2T3*, and *GOSR2* in CD79A+ B-cells, *WDR1*, *PPP1CC*, *ISG20* in T-cells, *SERPINA1*, *APOE*, *LRP1* in macrophages, *SLC24A3* in mast-cells, and *PREX1* in natural killer cells (**Figure 5**). These data are also available through www.plaqview.com together with other plaque-derived scRNAseq datasets^18^.

**Figure 5.**
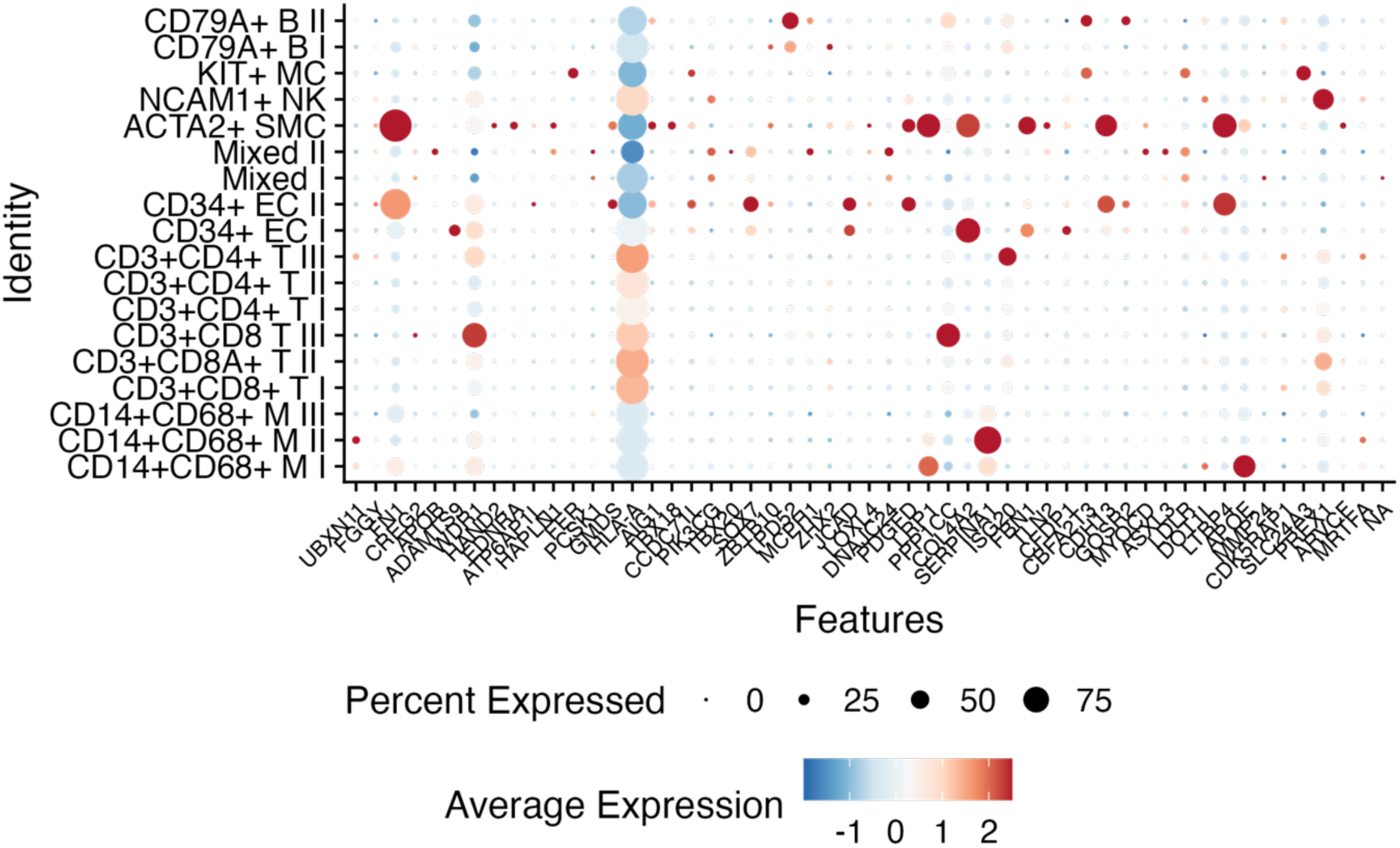
Cell-type specific expression in carotid plaques of candidate genes. The y-axis shows cell types as previously described ^16^. The size of the dot shows the percentage of cells expressing the respective genes, and the colour indicates the average total expression in that cell-types.

We tested the 51 lead variants for association with plaque histological markers in 1500 patients from the Athero-Express Biobank. Of note was the significant association of the *APOE* locus with plaque fat content, and positive correlation with collagen content, calcification, and intraplaque vessel density at the *CFDP1*, *FGGY*, and *MCPH1* loci, respectively (**ST21**).

## Discussion

We performed a large-scale multi-ancestry GWAS meta-analysis of cIMT, aggregating data from seven studies or consortia, comprising more than ∼130,000 participants. The meta-analysis identified 53 cIMT-associated loci for maximum mean cIMT, of which 19 were novel. An additional two loci were identified when we accounted for heterogeneity in allelic effects between ancestral groups. A sex-specific GWAS using the UK Biobank, identified 17 loci, 11 for women and 6 for men, with 14 overlapping with sex-combined GWAS. We highlighted 4 potentially causal genes that are highly expressed in the arterial wall of smooth muscle, endothelial cells, and inflammatory cells, and show evidence for locus-specific effects on plaque morphology. We found that circulatory levels of protein products of seven identified genes, including ACAN, BCAM, DUT, ERI1, APOE, FN1, and GLRX were potentially causally associated with cIMT levels. We found a considerable genetic correlation between anthropometric measures, glycaemic homeostasis, and CAD, however less with lipids (except for HDL-cholesterol). Mendelian randomisation analyses showed significant associations with pulse pressure, risk of any stroke, ischemic stroke, and large artery stroke as well as a strong and potentially bidirectional causal relationship between SBP and cIMT.

Our study is a large multi-ancestry GWAS meta-analysis for cIMT performed in individuals from five diverse populations including, African, Asian (Chinese), Brazilian, European, and Hispanic. Our findings further support the efficiency of conducting multi-ancestry GWAS. First, by using a Bayesian fine-mapping approach in European-only and multi-ancestry GWAS, we showed that multi-ancestry meta-analysis resulted in considerable refinement of causal variants (due to less extensive linkage disequilibrium) at 22 loci associated with cIMT. Second, the identification of 12 additional loci associated with cIMT from the multi-ancestry meta-analysis emphasises the importance of aggregating GWAS results from diverse populations to increase sample size and statistical power which is in line with previous research^19^.

Observational research has linked modifiable cardiovascular risk factors with cIMT but whether the links are causal is unclear. Here for the first time, using a Mendelian randomisation framework, we present evidence for the causal role of anthropometric measures, blood pressure, smoking and dyslipidaemia in subclinical atherosclerosis. We provide strong evidence of a causative effect of higher waist circumference on higher cIMT suggesting that central fat distribution (abdominal obesity) is a causal risk factor for cIMT. Waist circumference is associated with subclinical atherosclerosis^20^, and it may influence changes in cIMT, possibly via its effect on insulin resistance and glucose levels^21,22^. Our MR results are consistent with the results from epidemiological studies which showed that HDL-C is inversely associated with cIMT^23^, while increased levels of total cholesterol and LDL-C are positively associated with cIMT.

In bidirectional two-sample MR analyses, we found causal associations between SBP and cIMT, and vice versa. These associations remained robust in sensitivity analyses as well as after excluding genetic variants associated with both cIMT and SBP. Studies have suggested that elevated blood pressure results in increased cIMT possibly by the vascular damage caused by local shearing force in this condition^24^. On the other hand, increased cIMT may result in increased blood pressure by a possible mechanism leading to vascular endothelial dysfunction^24^ and arterial stiffness.

Several studies have linked cardiovascular diseases with AD, possibly due to shared risk factors between these two diseases, especially APOE which is a known gene for cIMT and AD. Moreover, there is growing evidence to suggest that vascular dysfunction plays a central role in the development of AD. Thus, we did a lookup of the cIMT-associated variants in AD GWAS and investigated the potential causal role of cIMT in AD using Mendelian randomisation analysis. Although previous research has repeatedly implicated vascular factors induced by cardiovascular diseases in AD pathology^25^, our MR analysis did not find any causal association between cIMT and AD, indicating that factors such as atherosclerosis might not be materially involved in the pathophysiology of AD. *APOE* is a pleiotropic genetic locus and could be linked to cardiovascular diseases and AD through different pathways.

Cross-sectional studies have shown higher cIMT in men compared to women; however, the contribution of genetic factors to these differences is largely unknown. We ran a sex-stratified analysis and identified two novel loci (*COX7A2L* and *CDK13*) that were genome-wide significant in women but not in men. *COX7A2L*, an estrogen-responsive gene, regulates energy metabolism, its deficiency has been shown to lower blood glucose levels^26^, and it has also been associated with thrombosis (P =2 x10^-7^)^27^. *De novo* mutations in *CDK13* have been associated with congenital heart defects^28^. Our sex-specific GWAS had a fairly balanced gender breakdown (48% men versus 52% women), similar to that of previous UKBB cIMT GWAS, so it is less likely that the lack of association in men is due to a lack of statistical power. Previous studies have suggested that the sex difference in cIMT could be explained by differences in cardiovascular risk factors, including obesity and blood pressure^29^.

However, we found two loci that were only significant in women, and we showed that the genetic effects at the *APOE* locus are stronger in men (β= 0.030) compared to women (β= 0.018). This might indicate that the sex differences in cIMT levels are not fully driven by risk factor distribution but also have roots in genes.

The strength of our study is the large sample size and multi-ancestry meta-analysis, which has enabled us to significantly enhance the number of known loci. We further searched for the role of rare variants in the genetic architecture of cIMT using whole-genome sequence data from TOPMed and applied gene-based methods. We applied various methods to study the functional characteristics of the identified genetic loci. Finally, we used various methods including genetic correlation, locus lookups, colocalization, and Mendelian randomisation to study the link between cIMT and cardiovascular risk factors and clinical outcomes. Several limitations should be mentioned. First, this study has collected all available studies in the discovery panel to maximise the statistical power and therefore findings are not replicated in independent samples. Second, the cIMT measurement protocols were slightly different across participating studies, however, any systematic differences in calibration should have been taken care of after variable transformations. Moreover, the findings are not driven by a single study. Third, the majority of the samples analysed in the current study were of European ancestry, which highlights the need for further genome-wide scans in populations other than those of European ancestry.

In conclusion, we report the largest multi-ancestry GWAS meta-analysis of cIMT. These findings extend our understanding of the genetic architecture of cIMT. We have comprehensively studied the link between cardiovascular risk factors, cIMT, and complex disorders. We report that systolic blood pressure and cIMT could have a bidirectional synergic effect on each other.

## Methods

### Study population

#### UKBB sample

The UK Biobank (UKBB), a large-scale prospective epidemiological cohort, recruited ∼500,000 individuals aged 40-69 years from across the United Kingdom between 2006 and 2010. In 2014, UKBB began a pilot phase for imaging modalities relevant to cardiovascular research, including ultrasound of the carotid arteries in 100,000 participants^30^. Carotid intima-media thickness (cIMT) phenotyping was performed at one of the UKBB imaging centres. A detailed description of carotid ultrasound protocol (UKBB Resource 511, https://biobank.ctsu.ox.ac.uk/crystal/refer.cgi?id=511) is provided elsewhere^31^. After performing quality control on phenotypic data, 42,449 cIMT measurements were available for further analysis. We use natural log-transformed values of the mean of the maximum (IMTmean-max) and mean of the mean (IMT mean) cIMT for normality. A detailed description of genotyping measures, imputation and central QC performed by the UKBB can be found elsewhere^32^. The Genome-Wide Association Study (GWAS) of cIMT was performed using a linear mixed non-infinitesimal model as implemented in BOLT-LMM^33^, adjusted for age, sex, genotyping centre and first four principal components. Strict quality control filters were applied on genotypic as well as genome-wide imputed data (**Supplementary Tables 1-2**).

#### CHARGE-UCLEB

We obtained summary statistics from a previous GWAS meta-analysis of cIMT reported by the CHARGE-UCLEB consortia^9^. This GWAS meta-analysis analysed ∼9.5 million non-sex-linked SNPs in up to 71,128 individuals of European ancestry from 31 studies ^8,9^. Each participating study excluded SNPs and samples based on standard quality measures (**ST 1-2**). cIMT was defined by the mean of the maximum of several common carotid artery measurements, measured at the far wall or the near wall.

#### TOPMed

Whole genome sequencing (WGS) data from the Trans-Omics for Precision Medicine (TOPMed) program was used, involving 24,953 individuals from 9 different studies and 4 different ancestry groups. cIMT was measured using carotid ultrasound measurements with two primary measures of IMT were presented: 1) IMTmean, and 2) IMTmean-max. Phenotype harmonization was performed by the TOPMed Data Coordinating Centre. A GWAS of cIMT was performed using the software package ‘genesis’^34^ adjusting for age, sex, study and 11 principal components (PCs) (**ST 1-2**).

#### AWI-Gen

AWI-Gen, a population-based cross-sectional study, included populations from six sub-Saharan African sites in four countries (Burkina Faso, Ghana, Kenya and South Africa)^35^. Over 12,000 sub-Saharan Africans, aged from 40 to 60 years, were enrolled from rural and urban settings from 2012 to 2016 ^35,36^. Ultrasound scans were performed to assess the right and left cIMT (two measurements: IMTmean-max and IMTmean) using dual B-mode ultrasound images of the left and right common carotid arteries. Samples were genotyped using the H3Africa genotyping array on ∼2.3 million SNPs. A GWAS of cIMT was performed using fastGWA^37^ software, adjusting for age, sex, study site and first four PCs (**ST 1-2**).

#### MACAD and HTN-INR

The MACAD and HTN-IR studies were designed to examine the genetic basis of coronary artery disease and insulin resistance in Mexican-Americans using a family-based design. cIMT was measured using standardized procedures and technology developed specifically for longitudinal measurement of atherosclerosis^38^ (Patents 2005, 2006, 2011). The coefficient of variation of repeated cIMT measurements is typically <3% and often approaches 1%. Samples with call rates >0.98, SNPs with call rates >0.99, MAF >0.001, and HWE (P<1×10^-6^) passed laboratory quality control ^39^. Pedigrees were examined for consistency of stated family structure. Software package ‘Rvtests’^40^ was used for cIMT genetic association analysis adjusting for age, sex and admixture proportions (**ST 1-2**).

#### BHS

The Baependi Heart Study is an epidemiological study in Baependi, a rural city in Minas Gerais State, Brazil. We used a cross-sectional analysis of data collected at the second evaluation visit (from 2010 to 2015) on subjects that underwent carotid ultrasonography. Individuals with angina, infarction, cardiac insufficiency, and revascularization were removed from our analysis. Imputation was performed using the Haplotype Reference Consortium Michigan Imputation Server using the TOPMed reference haplotype panel as a reference.

After imputation markers were kept if R^2^>0.3, MAF>0.01 and HWE p-value >1×10^−20^. The cIMT GWAS was performed using FaST-LMM ^41^ adjusting for age, sex and the first four PCs (**ST 1-2**).

### Pre-meta-analysis quality control (QC) and Statistical analyses

Pre-meta-analysis QC on UKBB and CHARGE-UCLEB, TOPMed, AWI-Gen, HTN-IR, MACAD, and BHS summary-level data was performed using the R software package ‘EasyQC’^42^. We used Haplotype Reference Consortium (HRC)^43^ for UKBB and the 1000 Genomes phase 1 version 3 for CHARGE-UCLEB. We excluded markers based on missing alleles, allele frequency, effect size estimates and their associated standard errors. After performing central QC on all participating cohorts, ∼16M autosomal markers were available for meta-analysis. Meta-analysis of summary statistics was performed using the inverse-variance fixed-effects method as implemented in the software METAL^44^. To account for heterogeneous loci, we also implemented a meta-regression approach using MR-MEGA v0.2^15^ (https://genomics.ut.ee/en/tools). Principal components generated using MR-MEGA are shown in Supplementary Figure 10.

### Functional annotation of cIMT-associated variants

We used the web-based tool Functional Mapping and Annotation (FUMA) v1.3.6^45^ for annotating the cIMT-associated variants identified in the multi-ancestry meta-analysis. Independent significant SNPs with P <5×10^−8^ in the meta-analysis, and in moderate linkage disequilibrium (LD) (r^2^<0.6) within a 500 kb window were used to define a locus. These ‘independent significant SNPs’ were further narrowed down to ‘lead SNPs’ having r^2^ <0.1 with each other ^45^. The most likely causal genes associated with cIMT were identified using positional mapping, eQTL mapping, and chromatin interaction mapping, prioritising 246 genes. Tissue expression analysis employed the results originating from the gene-based genome-wide association analysis using MAGMA.

<u>Gene-based analysis</u>: The gene-based genome-wide association analysis was performed using MAGMA v1.07^46^ as implemented in FUMA. This analysis used GWAS-identified SNPs and mapped them to protein-coding genes. A principal components regression model computed gene-level p-value (F-test)^46^ and multiple testing was addressed using Bonferroni correction (α = 2.6×10^-6^ = 0.05/19220).

<u>Gene-set and tissue expression analysis</u>: A gene-set analysis was performed using MAGMA’s competitive testing as implemented in FUMA to identify key biological pathways. This analysis tests whether genes in a gene-set are more strongly associated with a phenotype of interest than other genes. A gene-property analysis was also applied, testing for the relationship between highly expressed genes in specific tissue types and genetic associations.

### Conditional analysis using COJO

A stepwise model selection procedure was performed using the COndition and JOint analysis tool implemented in the Genome-wide Complex Trait Analysis software^47^ (COJO-GCTA; cojo-slct) to select independently associated SNPs. For each locus where we observed multiple independent signals, the lead SNP was considered the first independent signal and a conditional analysis was performed by selecting SNPs within 500 kb of the lead SNP, using the 1000 genomes data as reference panel (N=503). Conditional analysis included SNPs with MAF > 0.01 and a p-value <= 5×10^-8^.

### Statistical fine-mapping

We performed Bayesian fine-mapping by constructing 95% credible sets for each locus identified in the European-only GWAS and compared it with the multi-ethnic meta-analysis. Index SNPs were selected for all 39 loci identified in the European-only GWAS and defined regions for fine-mapping by extracting all SNPs within a 50 kb upstream/downstream window around each index SNP. For each SNP within a given region, Bayes factor (BF) was calculated based on GWAS p-values and the posterior probability that the SNP is causal (SNP’s BF / summation of the BFs in the region)^48^. We generated 95% credible sets by summing the posterior probabilities until the cumulative value exceeded 95% of the total cumulative posterior probability for all SNPs in the region.

### Estimating heritability and genetic correlation with other traits

Genome-wide genetic correlations between cIMT GWAS (European-only) summary statistics and various cardiometabolic, anthropometric, and glycemic traits and diseases were evaluated to investigate the shared genetic architecture. Genetic correlation analysis was performed through the web-platform LDHub^49^. Statistical significance for the genetic correlation analysis was defined at P=0.00094 (0.05/53) after correction for the number of evaluated associations. We used linkage disequilibrium score regression to estimate the SNP-based heritability. The summary level data from CHARGE-UCLEB and UK Biobank was used for this analysis, restricted to HapMap3 SNPs as these are well-imputed in most studies.

### Two-sample univariable Mendelian randomization

We performed two-sample Mendelian randomisation (MR) using ‘TwoSampleMR’ package to investigate both the causal role of various risk factors on cIMT and the causal role of cIMT on various outcomes. For MR analysis, we used the genetic association for cIMT from the European-only meta-analysis in this study. The details of risk factors and outcomes are available in Supplementary Table 21, including sources of genetic association and sample size of the corresponding GWAS. For each MR analysis, we selected SNPs associated with the exposure of interest at P-value <5×10^-8^, with an F-statistic >10, and MAF > 5%. The genetic associations of the same SNPs with the outcome of interest were obtained from the corresponding outcome GWAS (**ST18**). The independent (r^2^<0.001) harmonised SNPs were used as genetic instruments for the MR analysis.

SNP-specific effects for an exposure-outcome pair were estimated using the Wald ratio. We used the multiplicative random-effects inverse-variance weighted (IVW) method to pool the SNP-specific estimates for the MR effect estimates. We also used two sensitivity methods, namely weighted median, and MR-Egger regression^50^. In all analyses, we excluded potential outlier SNPs identified by MR-PRESSO^51^. Specifically for the bidirectional analysis for cIMT and SBP, we performed a sensitivity analysis excluding common SNPs associated with both traits. We excluded 43,514 common SNPs and their proxies (r^2^≥0.001). We also used MR-Steiger^52^ to investigate the direction of causality between cIMT and SBP.

We accounted for multiple comparisons using Bonferroni correction (α = 0.05/13 (N risk factors and outcomes) =0.004). For cis-MR on circulatory proteins, we used independent (r2=0.001) protein quantitative trait loci (pQTL) (p-value < 5×10^-8^) from Sun et al 2023^53^ within +/-500kb of the coding gene.

### Single-cell expression in carotid plaque

The Athero-Express Biobank Study (www.atheroexpress.nl) in Utrecht, the Netherlands collected atherosclerotic plaques during surgery blood and plaques from patients undergoing carotid endarterectomy and stored at - 80°C ^16,54^. The plaques were analysed to quantitatively score the number of macrophages (CD68) and smooth muscle cells (SMCs, α-actin), as a percentage of the microscope field area by computerized analysis. Vessel density (CD34) was assessed as the average number per 3 hotspots. Intraplaque haemorrhage (IPH), and intraplaque fat, were scored using hematoxylin and eosin staining (HE). Calcification (using HE), and collagen (picrosirius red) were scored binary ^55^.

DNA was isolated for genetic analysis of these plaque characteristics ^56^. The study was conducted in three separate experiments: Athero-Express Genomics Study 1 (AEGS1) including 891 patients, the Athero-Express Genomics Study 2 (AEGS2) including 954 patients, and the Athero-Express Genomics Study 3 (AEGS3) including 658 patients. All experiments were carried out according to OECD standards and we adhered to community standard quality control and assurance (QCA) procedures of the genotype data as described before^56,57^. After QCA these comprise 890 samples and 407,712 SNPs in AEGS1, 869 samples and 534,508 SNPs in AEGS2, and 649,954 samples and 534,508 SNPs in AEGS3 remained. Missing genotypes were imputed with 1000G phase 3, version 5 and HRC release 1.1 as a reference using the Michigan Imputation Server^58^. These results were further integrated using QCTOOL v2, where HRC imputed variants are given precedence over 1000G phase 3 imputed variants.

The association of 51 independent loci with plaque characteristics was analysed adjusted for age, sex and principal components. Continuous variables were inverse-rank normal transformed. Codes and raw results from these analyses are found here: https://github.com/CirculatoryHealth/CHARGE_CIMT.

For single-cell RNA sequencing, we collected carotid plaques from 35 individuals, isolated and processed RNA as described before^59^, while employing the CEL-seq2 method^60^.

## Contributors

D.M., J.G.T., J.C.B., N.D.P., N.H.C., Y.I.M., G.H., D.R.J.J., M.F., A.B.N., J.B., A.K., F.F.W., B.I.F., L.A.L., J.G.W., D.M.H., S.S.R., W.A.H., A.H.X., H.N.H., M.C.M., J.B., M.R., B.P.R., A.C.M., B.D.M., J.J.C., B.M.P., D.W.B., R.S.V., A.C., J.I.R., W.S.P., M.O.G., L.J.R., A.C.P., J.E.C., P.S.V., A.D., I.T. contributed to the phenotype acquisition or harmonization including study design or funding of contributing studies. D.M., D.J., N.H.C., K.D.T., J.G.W., S.S.R., Y.D.I.C., M.C.M., J.B., J.D.S., R.A.G., H.V.D., K.A.V.M., M.R., B.P.R., B.D.M., A.C., J.I.R., A.C.P., J.E.C., A.D., I.T. contributed to acquisition of genotyping data or quality control. D.M., J.H., N.R.H., D.J., X.G., J.Y., J.T., M.R., B.P.R., A.C.P., P.S.V., A.D., I.T., S.W.L. contributed to statistical analysis of the data. D.M., J.H., D.J., M.R., B.P.R., P.S.V., A.D., I.T., S.W.L. contributed to the drafting of the manuscript. All authors contributed to the critical revision of the manuscript.

## Declaration of interest

C.J.O. is a full-time employee at Novartis Institute of Biomedical Research. S.W.L. has received Roche funding for unrelated work. R.W.K. is an employee at Psomagen Inc. B.M.P. serves on the steering committee of the Yale Open Data Access Project funded by Johnson & Johnson. M.E.M. receives funding from Regeneron Pharmaceuticals Inc. unrelated to this project. KAV-M is an employee at Illumina Inc. All remaining authors declare no competing interests.

## Funding

This work is supported by the UK Dementia Research Institute at Imperial College, which receives its funding from UK DRI Ltd., funded by the UK Medical Research Council (MRC), Alzheimer’s Society and Alzheimer’s Research UK. A.D. is funded by a Wellcome Trust seed award (206046/Z/17/Z). R.M. is funded by the President’s PhD Scholarship from Imperial College London. S.W.L. is funded through EU H2020 TO_AITION (grant number:848146).

## Correspondence

Dr Abbas Dehghan, MD, PhD, Department of Epidemiology and Biostatistics, Imperial College London School of Public Health, London, UK.

## Data availability

The summary statistics of the cIMT European-only and multi-ancestry GWAS will be made available for download upon publication via the database of Genotypes and Phenotypes (dbGaP) and/or GWAS catalog (https://www.ebi.ac.uk/gwas/).

## Code availability

The following software packages were used for data analysis: R (https://www.r-project.org) version 3.6, EasyQC version 9.2, Plink1.9 (https://www.cog-genomics.org/plink/1.9); Plink2 (https://www.cog-genomics.org/plink/2.0); BOLT-LMM version v2.3.6, METAL (version 2011-03-25), FUMA version v1.3.6a (https://fuma.ctglab.nl), MAGMA version v1.08 (https://ctg.cncr.nl/software/magma), and LDHub (http://ldsc.broadinstitute.org/ldhub/), MR- MEGA v0.215 (https://genomics.ut.ee/en/tools).

## Acknowledgements

The UK Biobank data used in this work were obtained from UK Biobank Resource under application number 236. We are grateful to UK Biobank for making this data available, as well as to the generous participants who contributed their time to make this resource possible. Infrastructure for the CHARGE consortium is supported in part by the National Heart, Lung, and Blood Institute grant R01HL105756. TOPMed analyses were funded by the National Heart, Lung and Blood Institute (NHLBI) grant number R01HL146860. Paul S. de Vries and Natalie R. Hasbani were additionally supported by American Heart Association grant number 18CDA34110116. Molecular data for the Trans-Omics in Precision Medicine (TOPMed) program was supported by the NHLBI. See the TOPMed Omics Support Table (Supplementary Table 23) for study-specific omics support information. Core support including centralized genomic read mapping and genotype calling, along with variant quality metrics and filtering were provided by the TOPMed Informatics Research Center (3R01HL- 117626-02S1; contract HHSN268201800002I). Core support including phenotype harmonization, data management, sample-identity QC, and general program coordination was provided by the TOPMed Data Coordinating Center (R01HL-120393; U01HL-120393; contract HHSN268201800001I). Study-specific funding is detailed in the Supplementary texts. We thank L. Adrienne Cupples for her helpful comments that improved the manuscript. We thank Deborah A. Nickerson for her helpful contributions. We gratefully acknowledge the studies and participants who provided biological samples and data for TOPMed. Study-specific acknowledgements can be found in the supplementary document. We thank Rui P. Climaco for his help in producing Figure 5 in the main text. We are thankful for the support of the Netherlands CardioVascular Research Initiative of the Netherlands Heart Foundation (CVON 2011/B019 and CVON 2017-20: Generating the best evidence-based pharmaceutical targets for atherosclerosis [GENIUS I&II]), the ERA-CVD program ‘druggable-MI-targets’ (grant number: 01KL1802), and the Leducq Foundation ‘PlaqOmics’.

## Notes

### Author Declarations

The data used for this project was partly aggregate/summary data (from previous GWAS on cIMT) and partly individual level (UK biobank).

