## Supplementary figures for "Genome-wide association study and multi-ancestry meta-analysis identify common variants associated with carotid artery intima-media thickness"

**Supplementary Figure 1.**
Overall study design.......................................................................................................2

**Supplementary Figure 2.**
Manhattan plots of cIMT GWAS by study...........................................................................3

**Supplementary Figure 3.**
Quantile-Quantile plots of cIMT GWAS by study……….....................................................4

**Supplementary Figure 4.**
Regional plots of 51 genome-wide significant loci associated with cIMT…………………5-17

**Supplementary Figure 5.**
**Forest plots of 51 loci significantly associated with cIMT** …………………………18-30

**Supplementary Figure 6.**
Circos plots showing cIMT genomic risk loci and genes prioritised by eQTL and chromatin interaction …………………………………………………………………………………31-37

**Supplementary Figure 7.**
Miami plots for GWAS of cIMT in UK Biobank…………………………………………………..38

**Supplementary Figure 8.**
Comparison of effect size estimates and p-values from the UKBB $IMT_{mean}$ and $_{IMTmean-max}$ and sex-stratified analysis…………………………………………………......................39

**Supplementary Figure 9.**
Barplot presenting the number of SNPs in 95% credible sets at 39 loci identified in the European-only GWAS versus multi-ancestry …………………………………………………...40

**Supplementary Figure 1. Overall study design.** UKB, UK Biobank; CHARGE, Cohorts for Heart and Aging Research in Genomic Epidemiology; UCLEB, University College London-Edinburgh-Bristol consortia; TOPMed, Trans-Omics for Precision Medicine Program; AWI-Gen, Africa Wits-INDEPTH partnership for Genomic Studies; MACAD, Mexican-American Coronary Artery Disease; HTN-IR, the Hypertension-Insulin Resistance Family Study; BHS, Baependi Heart Study; GWAS, genome-wide association study; SNPs, single nucleotide polymorphisms; n, sample size; cIMT, carotid artery intima-media thickness; GWS, genome-wide significant; GWGAS, genome-wide gene-based analysis.

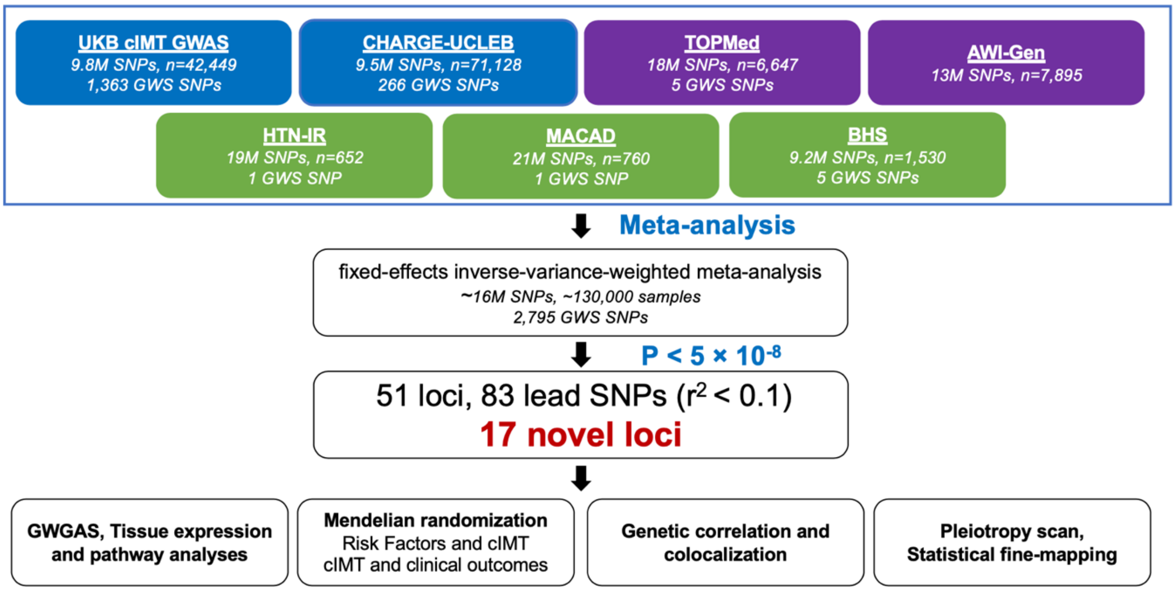

**Supplementary Figure 2**. Manhattan plots of the individual study GWAS results included in the multi-ancestry meta-analysis of cIMT in ~130,000 individuals. The plots show p-values (-$\log_{10}$) presented on the y-axis and chromosomal position on the x- axis for each participating study. The horizontal blue and red lines indicate the threshold for suggestive ($P<1\times10^{-5}$) and genome-wide ($P<5\times10^{-8}$) significance, respectively. CHARGE, Cohorts for Heart and Aging Research in Genomic Epidemiology; UCLEB, University College London-Edinburgh-Bristol consortia; TOPMed, Trans-Omics for Precision Medicine Program; AWI-Gen, Africa Wits-INDEPTH partnership for Genomic Studies; MACAD, Mexican-American Coronary Artery Disease; HTN-IR, the Hypertension-Insulin Resistance Family Study; BHS, Baependi Heart Study.

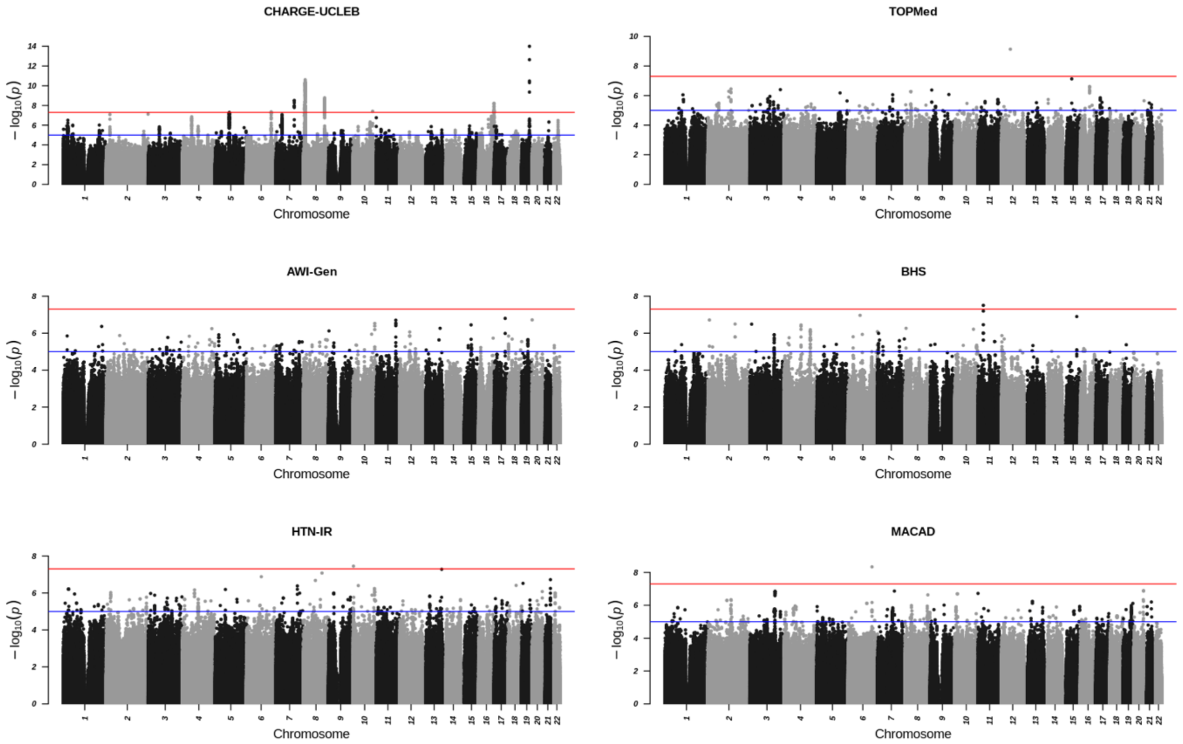

**Supplementary Figure 3.** QQ plots of the individual study GWAS results included in the multi-ancestry meta-analysis of cIMT in ~130,000 individuals. Observed -$\log_{10}$ transformed P-values of associations with cIMT measures are plotted against expected null P-values for all participating studies. CHARGE, Cohorts for Heart and Aging Research in Genomic Epidemiology; UCLEB, University College London-Edinburgh-Bristol consortia; TOPMed, Trans-Omics for Precision Medicine Program; AWI-Gen, Africa Wits-INDEPTH partnership for Genomic Studies; MACAD, Mexican-American Coronary Artery Disease study; HTN-IR, Hypertension-Insulin Resistance Family Study; BHS, Baependi Heart Study; λgc, genomic inflation factor.

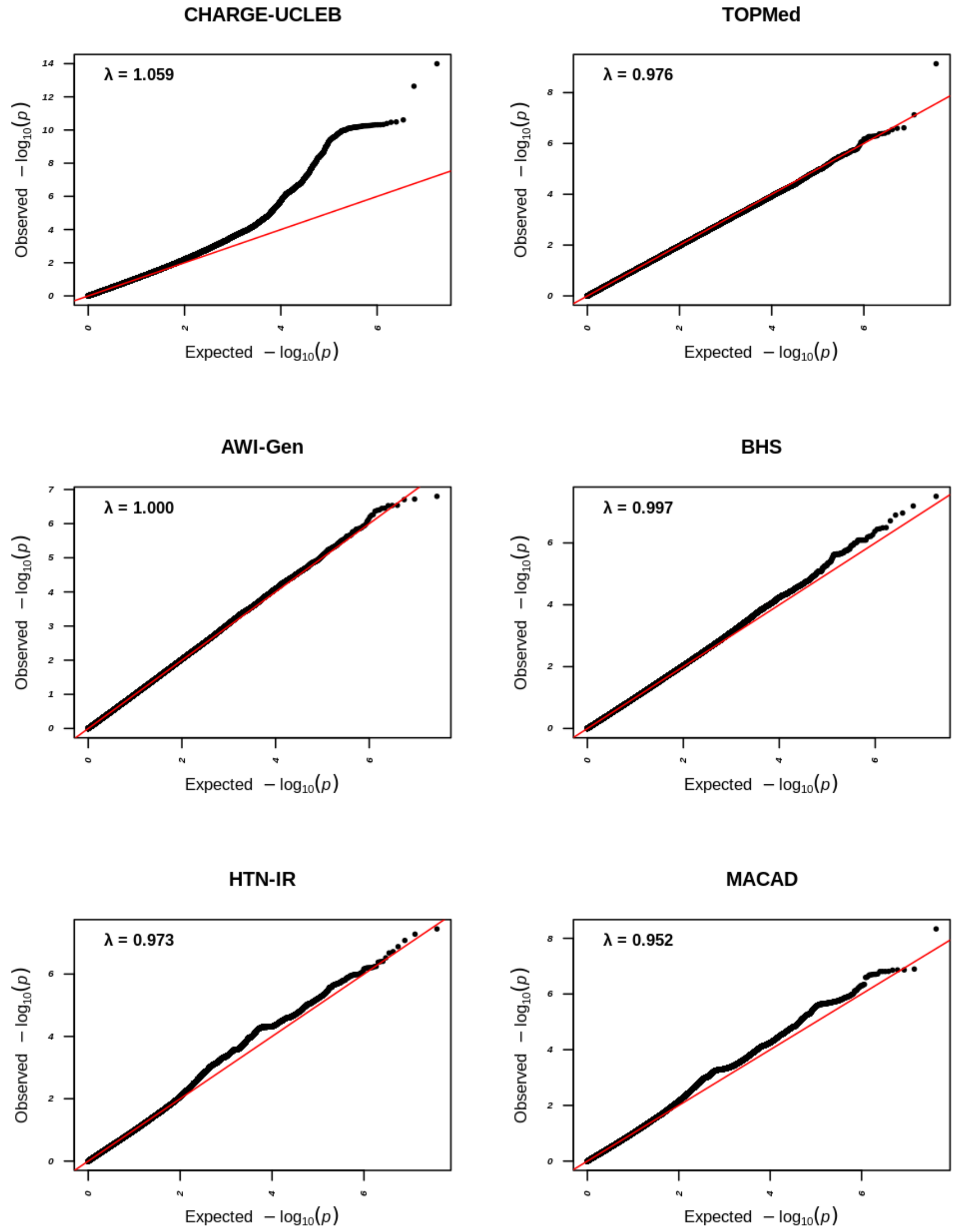

**Supplementary Figure 4.** Regional association plots for the 51 cIMT-associated loci from the multi-ancestry analysis. cIMT GWAS meta-analysis p-values (-$log_{10}$ transformed) were plotted against their genomic locations on chromosomes (starting from chromosome 1). The purple circle represents the strongest association. The colour-coded dots show the level of linkage disequilibrium with the peak SNP using 1000 genomes version 3 all populations. Genes are shown at the bottom.

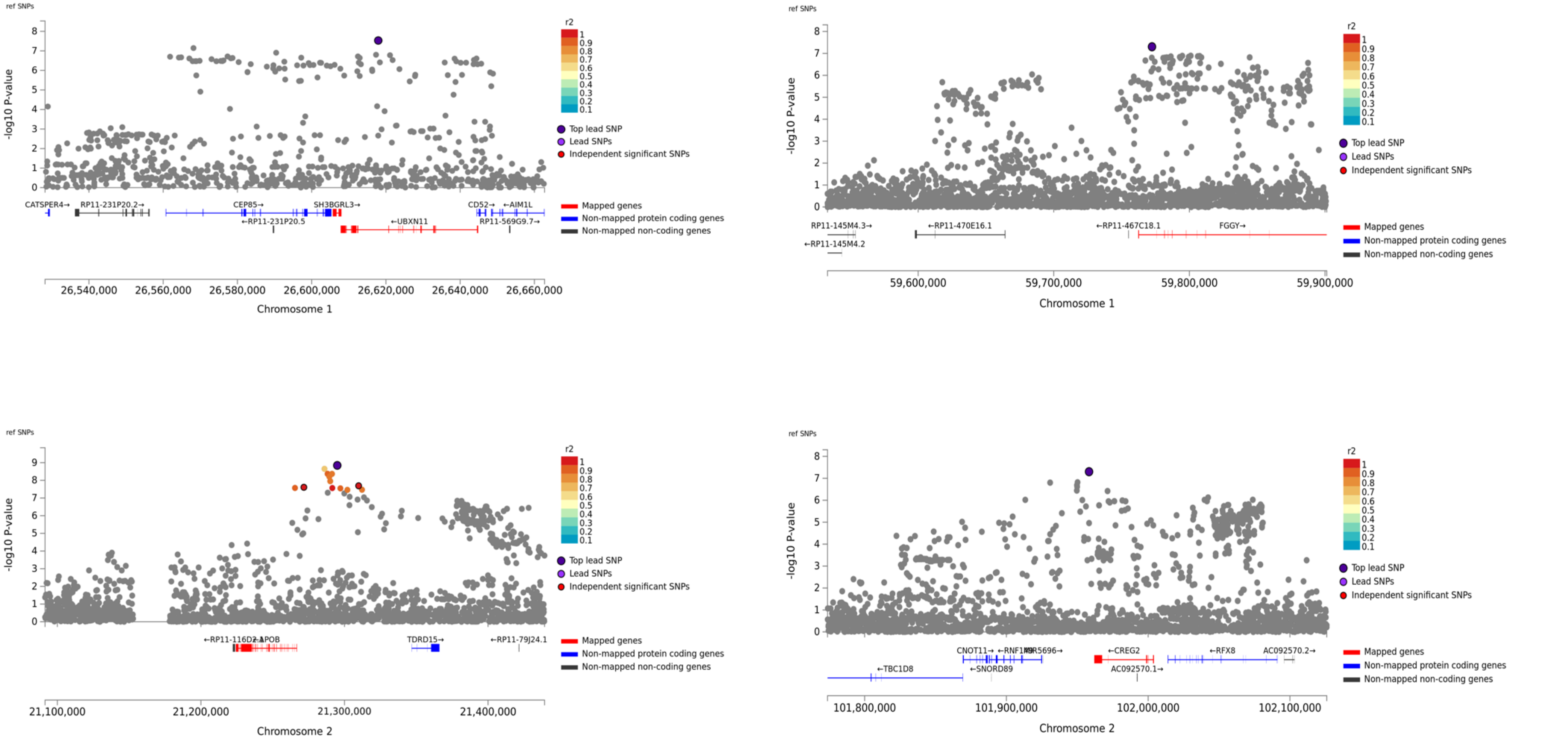

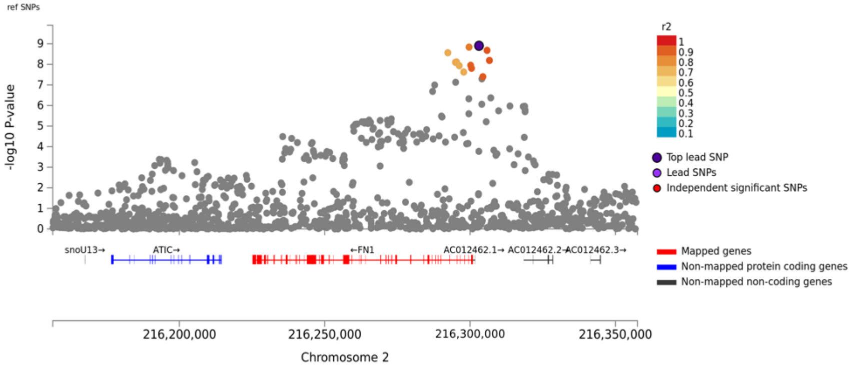
ref SNPs
-log10 P-value
r2
Top lead SNP
Lead SNPs
Independent significant SNPs
snoU13→
ATIC→
←FN1
AC012462.1→
AC012462.2→
AC012462.3→
Mapped genes
Non-mapped protein coding genes
Non-mapped non-coding genes
216,200,000
216,250,000
216,300,000
216,350,000
Chromosome 2

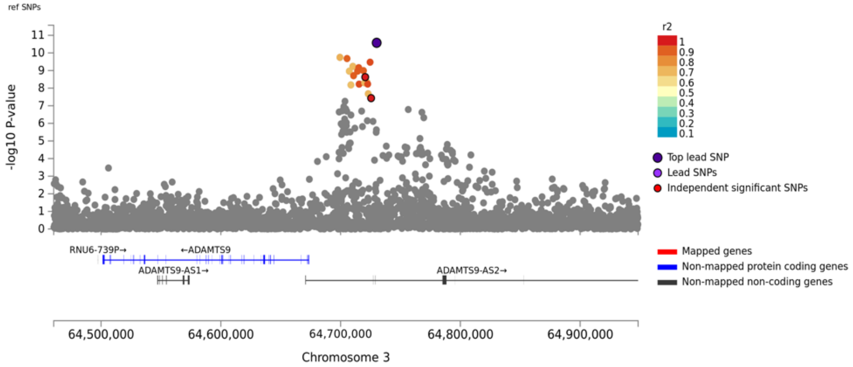
ref SNPs
-log10 P-value
r2
Top lead SNP
Lead SNPs
Independent significant SNPs
RNU6-739P→
←ADAMTS9
ADAMTS9-AS1→
ADAMTS9-AS2→
Mapped genes
Non-mapped protein coding genes
Non-mapped non-coding genes
64,500,000
64,600,000
64,700,000
64,800,000
64,900,000
Chromosome 3

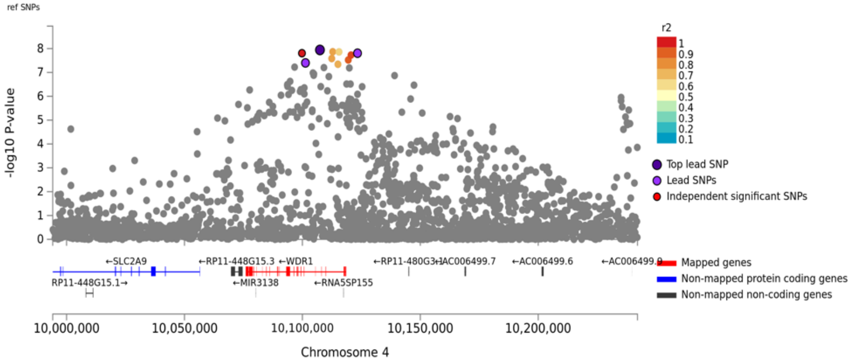
ref SNPs
-log10 P-value
r2
Top lead SNP
Lead SNPs
Independent significant SNPs
←SLC2A9
RP11-448G15.1→
←RP11-448G15.3
←WDR1
←MIR3138
←RNA5SP155
←RP11-480G3.1
AC006499.7
←AC006499.6
Mapped genes
Non-mapped protein coding genes
Non-mapped non-coding genes
10,000,000
10,050,000
10,100,000
10,150,000
10,200,000
Chromosome 4

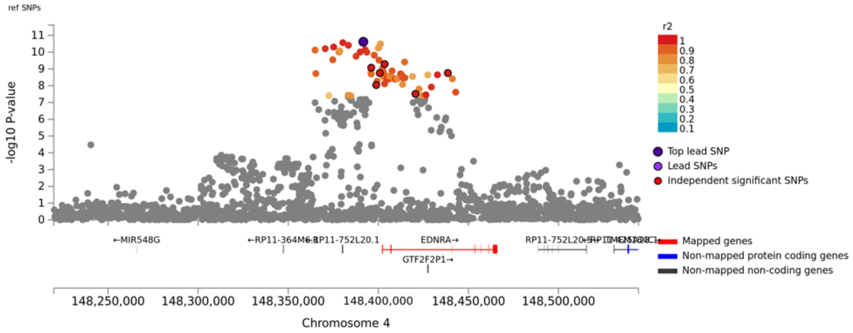
ref SNPs
-log10 P-value
r2
Top lead SNP
Lead SNPs
Independent significant SNPs
←MIR548G
RP11-752L20.1
EDNRA→
GTF2F2P1→
Mapped genes
Non-mapped protein coding genes
Non-mapped non-coding genes
148,250,000
148,300,000
148,350,000
148,400,000
148,450,000
148,500,000
Chromosome 4

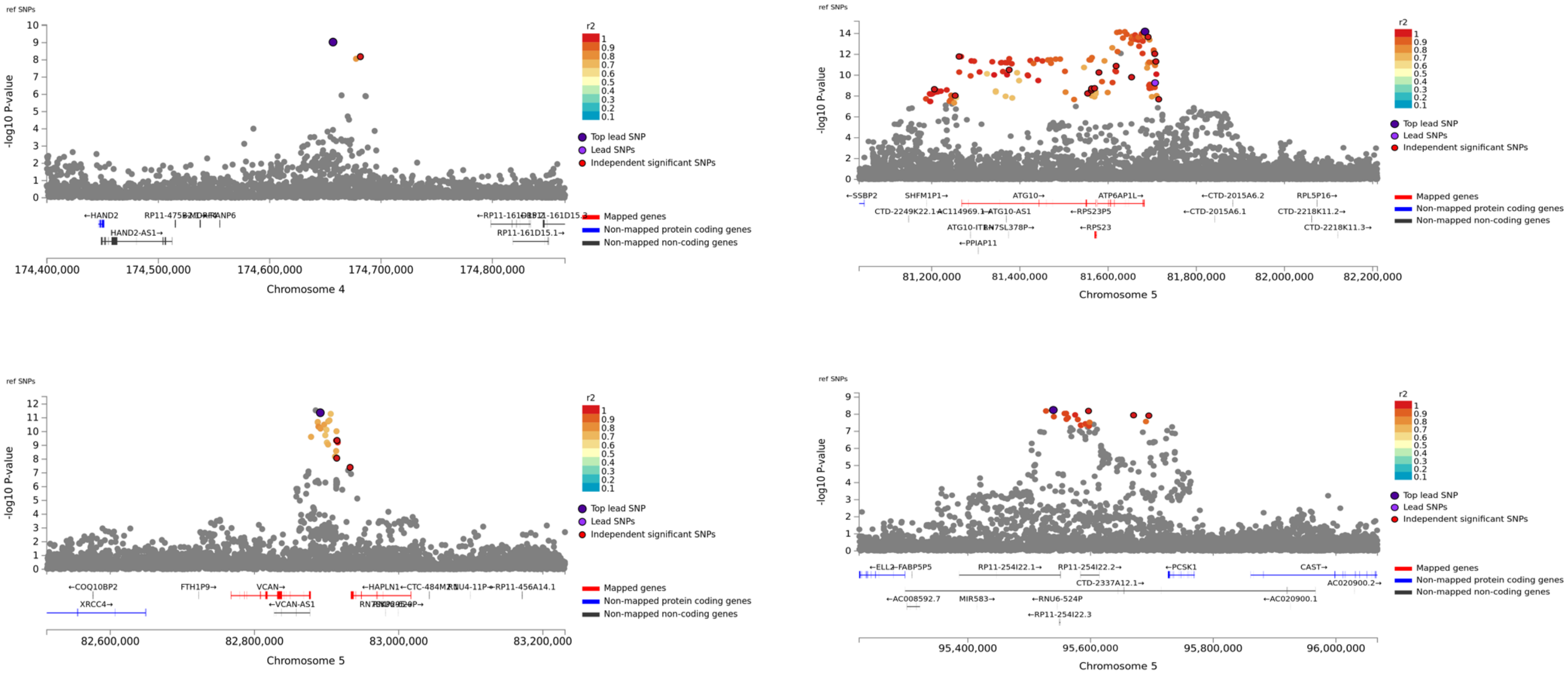

ref SNPs
-log10 P-value
r2
Top lead SNP
Lead SNPs
Independent significant SNPs
Mapped genes
Non-mapped protein coding genes
Non-mapped non-coding genes
Chromosome 4
Chromosome 5
Chromosome 5
Chromosome 5

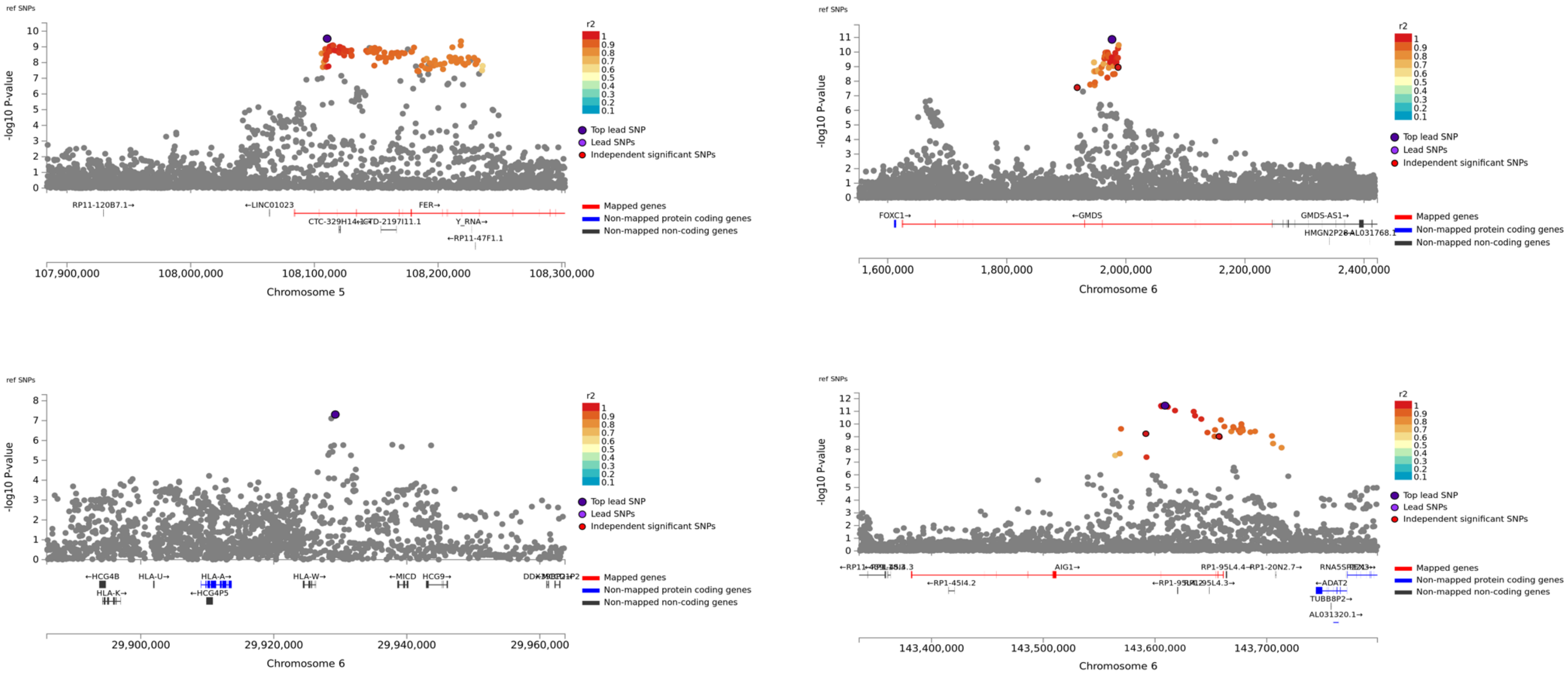

ref SNPs
-log10 P-value
r2
Top lead SNP
Lead SNPs
Independent significant SNPs
Mapped genes
Non-mapped protein coding genes
Non-mapped non-coding genes
RP11-120B7.1→
←LINC01023
FER→
Y_RNA→
←RP11-47F1.1
Chromosome 5
FOXC1→
←GMDS
GMDS-AS1→
Chromosome 6
←HCG4B
HLA-U→
HLA-A→
HLA-K→
←HCG4P5
HLA-W→
←MICD
HCG9→
Chromosome 6
AIG1→
←RP1-45I4.2
←ADAT2
TUBB8P2→
AL031320.1→
Chromosome 6

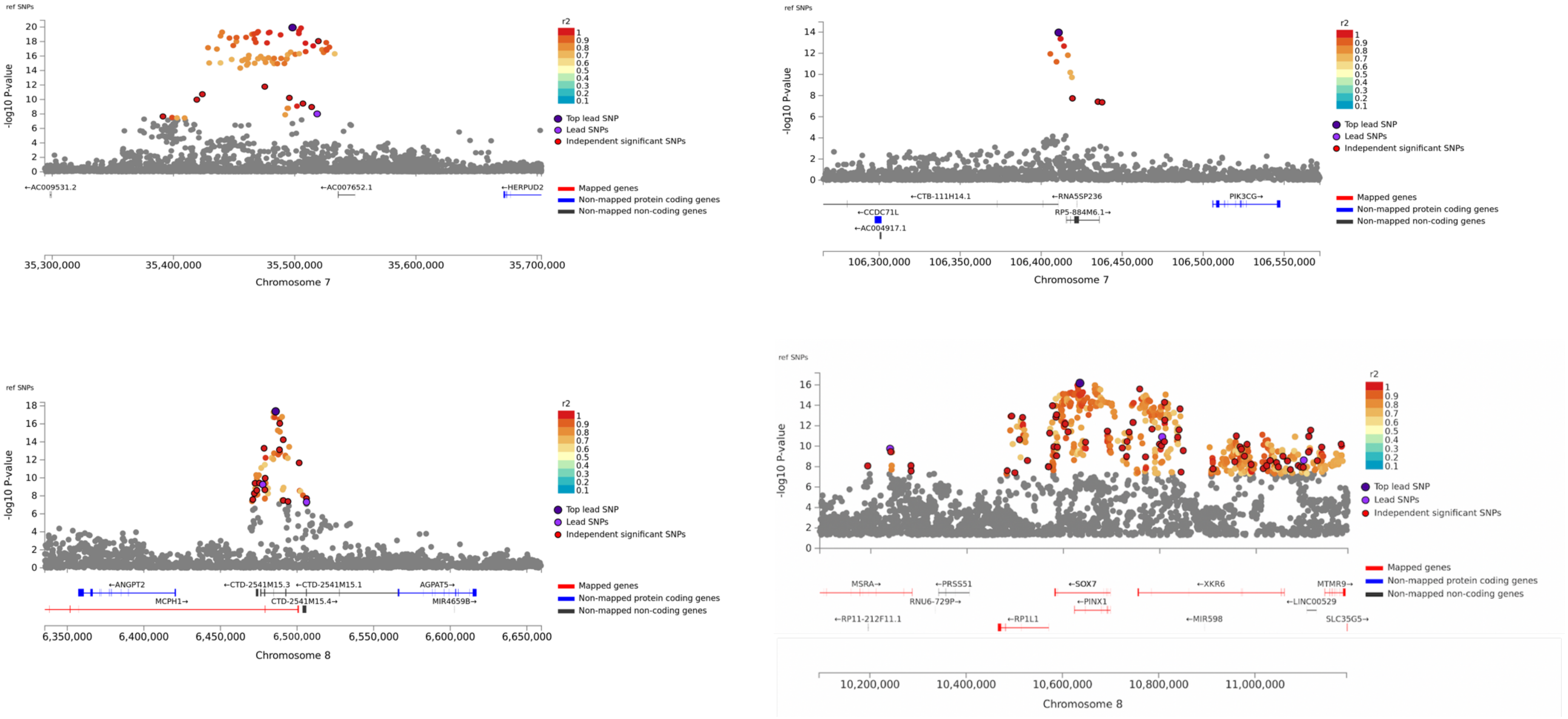

ref SNPs
-log10 P-value
r2
Top lead SNP
Lead SNPs
Independent significant SNPs
Mapped genes
Non-mapped protein coding genes
Non-mapped non-coding genes
←AC009531.2
←AC007652.1
←HERPUD2
35,300,000
35,400,000
35,500,000
35,600,000
35,700,000
Chromosome 7
←CTB-111H14.1
←RNASSP236
PIK3CG→
←CCDC71L
RP5-884M6.1→
←AC004917.1
106,300,000
106,350,000
106,400,000
106,450,000
106,500,000
106,550,000
Chromosome 7
←ANGPT2
←CTD-2541M15.3
←CTD-2541M15.1
AGPAT5→
MCPH1→
CTD-2541M15.4→
MIR4659B→
6,350,000
6,400,000
6,450,000
6,500,000
6,550,000
6,600,000
6,650,000
Chromosome 8
MSRA→
←PRSS51
←SOX7
←XKR6
MTMR9→
RNU6-729P→
←PINX1
←LINC00529
←RP11-212F11.1
←RP1L1
←MIR598
SLC35G5→
10,200,000
10,400,000
10,600,000
10,800,000
11,000,000
Chromosome 8

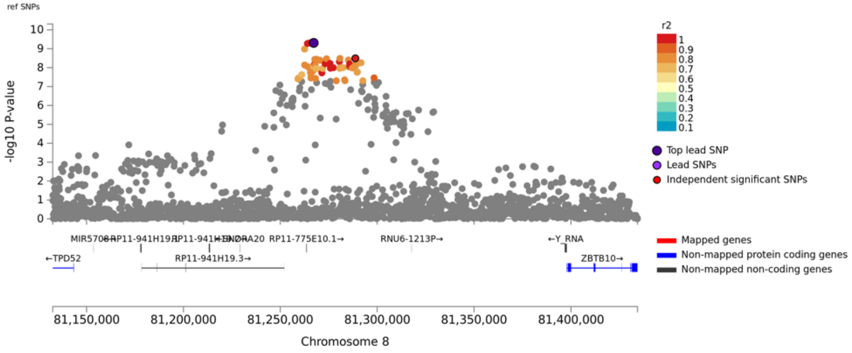

ref SNPs
-log10 P-value
r2
Top lead SNP
Lead SNPs
Independent significant SNPs
Mapped genes
Non-mapped protein coding genes
Non-mapped non-coding genes
←TPD52
RP11-941H19.3→
RP11-775E10.1→
RNU6-1213P→
←Y_RNA
ZBTB10→
81,150,000
81,200,000
81,250,000
81,300,000
81,350,000
81,400,000
Chromosome 8


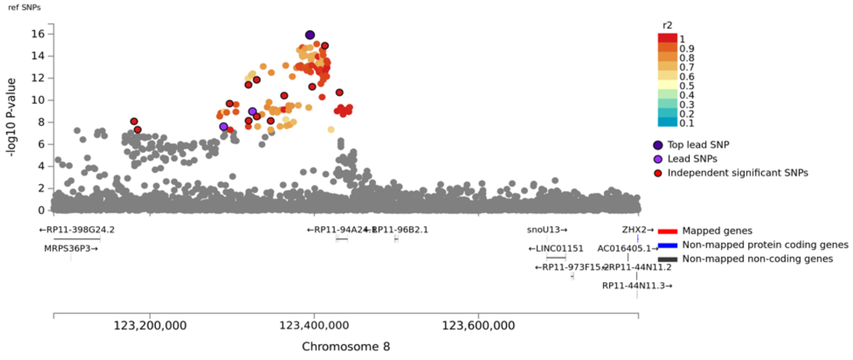

ref SNPs
-log10 P-value
r2
Top lead SNP
Lead SNPs
Independent significant SNPs
Mapped genes
Non-mapped protein coding genes
Non-mapped non-coding genes
←RP11-398G24.2
MRPS36P3→
snoU13→
ZHX2→
←LINC01151
RP11-44N11.3→
123,200,000
123,400,000
123,600,000
Chromosome 8


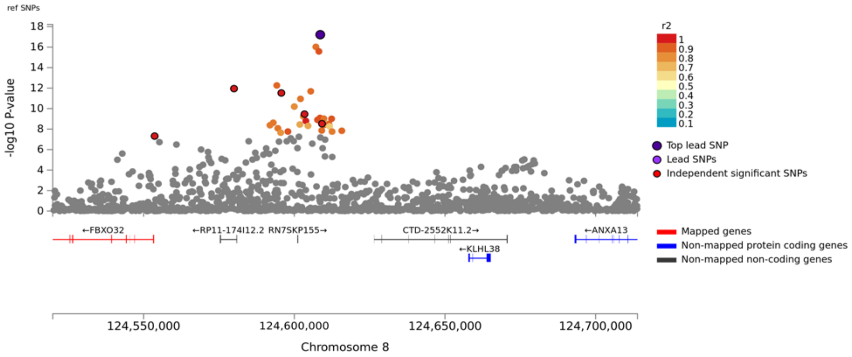

ref SNPs
-log10 P-value
r2
Top lead SNP
Lead SNPs
Independent significant SNPs
Mapped genes
Non-mapped protein coding genes
Non-mapped non-coding genes
←FBXO32
←RP11-174I12.2
RN7SKP155→
CTD-2552K11.2→
←KLHL38
←ANXA13
124,550,000
124,600,000
124,650,000
124,700,000
Chromosome 8


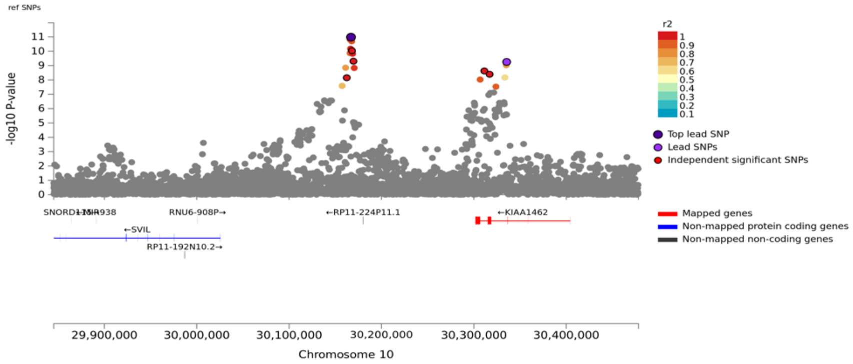

ref SNPs
-log10 P-value
r2
Top lead SNP
Lead SNPs
Independent significant SNPs
Mapped genes
Non-mapped protein coding genes
Non-mapped non-coding genes
RNU6-908P→
←RP11-224P11.1
←KIAA1462
←SVIL
RP11-192N10.2→
29,900,000
30,000,000
30,100,000
30,200,000
30,300,000
30,400,000
Chromosome 10

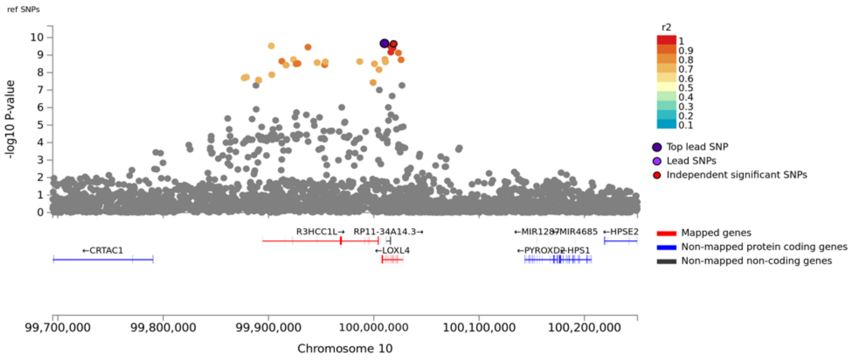
ref SNPs
-log10 P-value
r2
Top lead SNP
Lead SNPs
Independent significant SNPs
R3HCC1L→
RP11-34A14.3→
←CRTAC1
←LOXL4
←MIR1287
MIR4685
←HPSE2
←PYROXD2
HPS1
Mapped genes
Non-mapped protein coding genes
Non-mapped non-coding genes
99,700,000
99,800,000
99,900,000
100,000,000
100,100,000
100,200,000
Chromosome 10

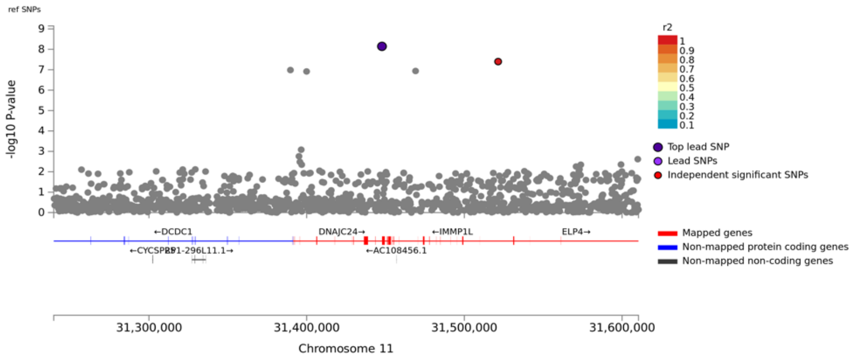
ref SNPs
-log10 P-value
r2
Top lead SNP
Lead SNPs
Independent significant SNPs
←DCDC1
DNAJC24→
←IMMP1L
ELP4→
←CYCSP26
RP11-296L11.1→
←AC108456.1
Mapped genes
Non-mapped protein coding genes
Non-mapped non-coding genes
31,300,000
31,400,000
31,500,000
31,600,000
Chromosome 11

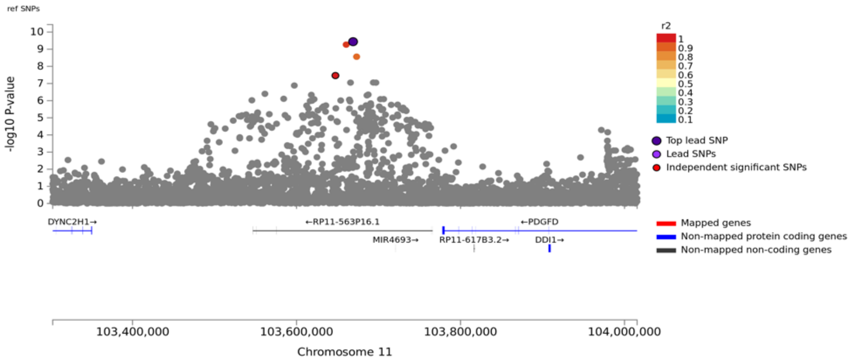
ref SNPs
-log10 P-value
r2
Top lead SNP
Lead SNPs
Independent significant SNPs
DYNC2H1→
←RP11-563P16.1
←PDGFD
MIR4693→
RP11-617B3.2→
DDI1→
Mapped genes
Non-mapped protein coding genes
Non-mapped non-coding genes
103,400,000
103,600,000
103,800,000
104,000,000
Chromosome 11

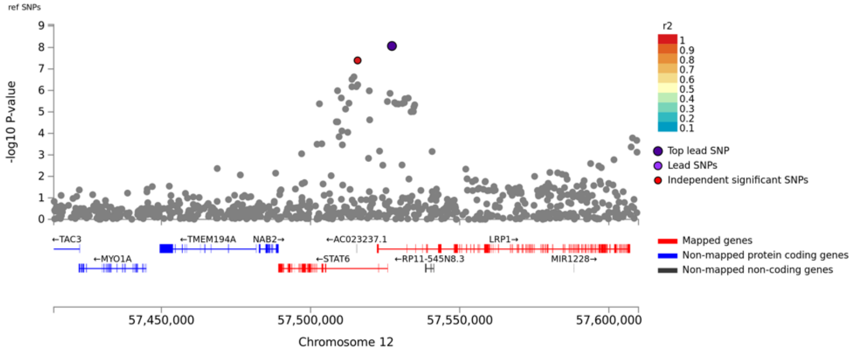
ref SNPs
-log10 P-value
r2
Top lead SNP
Lead SNPs
Independent significant SNPs
←TAC3
←TMEM194A
NAB2→
←AC023237.1
LRP1→
←MYO1A
←STAT6
←RP11-545N8.3
MIR1228→
Mapped genes
Non-mapped protein coding genes
Non-mapped non-coding genes
57,450,000
57,500,000
57,550,000
57,600,000
Chromosome 12

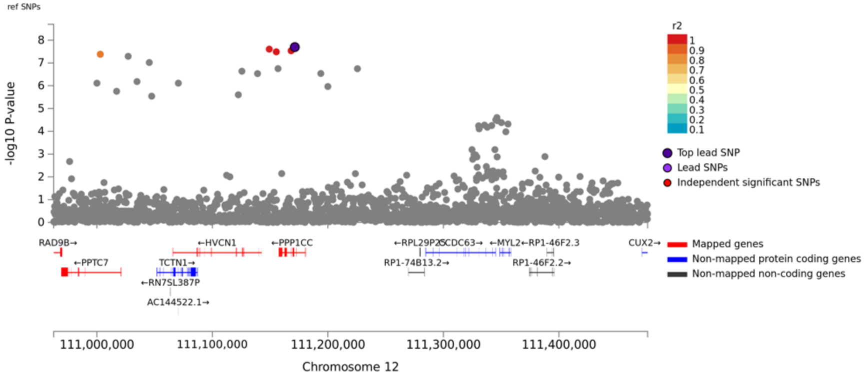
ref SNPs
-log10 P-value
r2
Top lead SNP
Lead SNPs
Independent significant SNPs
RAD9B→
←PPTC7
←HVCN1
TCTN1→
←RN7SL387P
AC144522.1→
←PPP1CC
←RPL29P25
CDC63→
←MYL2
←RP1-46F2.3
RP1-74B13.2→
RP1-46F2.2→
CUX2→
Mapped genes
Non-mapped protein coding genes
Non-mapped non-coding genes
111,000,000
111,100,000
111,200,000
111,300,000
111,400,000
Chromosome 12

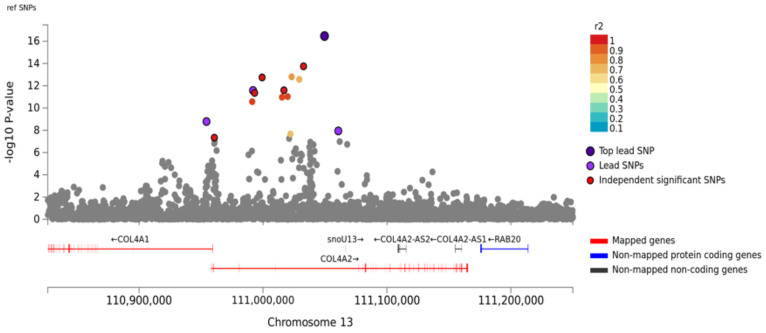
ref SNPs
-log10 P-value
r2
Top lead SNP
Lead SNPs
Independent significant SNPs
←COL4A1
snoU13→
←COL4A2-AS2
←COL4A2-AS1
←RAB20
COL4A2→
Mapped genes
Non-mapped protein coding genes
Non-mapped non-coding genes
110,900,000
111,000,000
111,100,000
111,200,000
Chromosome 13

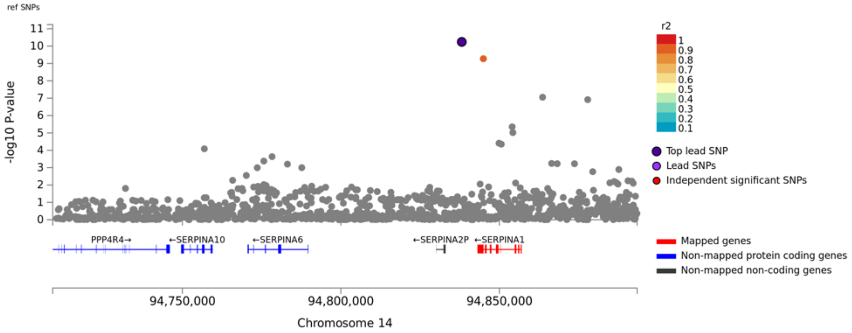
ref SNPs
-log10 P-value
r2
Top lead SNP
Lead SNPs
Independent significant SNPs
PPP4R4→
←SERPINA10
←SERPINA6
←SERPINA2P
←SERPINA1
Mapped genes
Non-mapped protein coding genes
Non-mapped non-coding genes
94,750,000
94,800,000
94,850,000
Chromosome 14

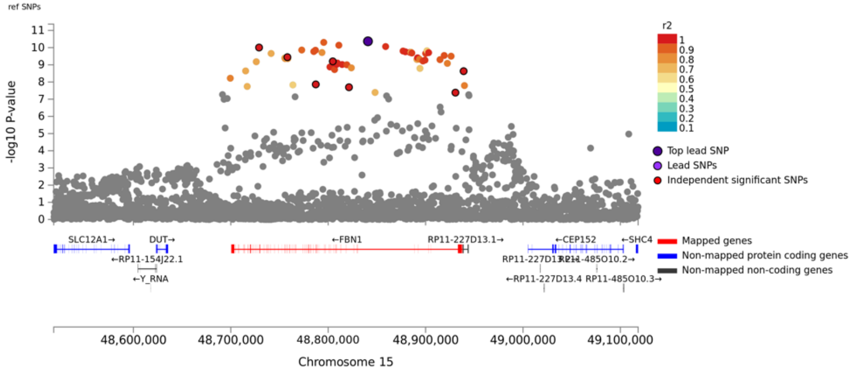
ref SNPs
-log10 P-value
r2
Top lead SNP
Lead SNPs
Independent significant SNPs
SLC12A1→
DUT→
←RP11-154J22.1
←Y_RNA
←FBN1
RP11-227D13.1→
←CEP152
←SHC4
←RP11-227D13.4
RP11-485O10.3→
RP11-485O10.2→
Mapped genes
Non-mapped protein coding genes
Non-mapped non-coding genes
48,600,000
48,700,000
48,800,000
48,900,000
49,000,000
49,100,000
Chromosome 15

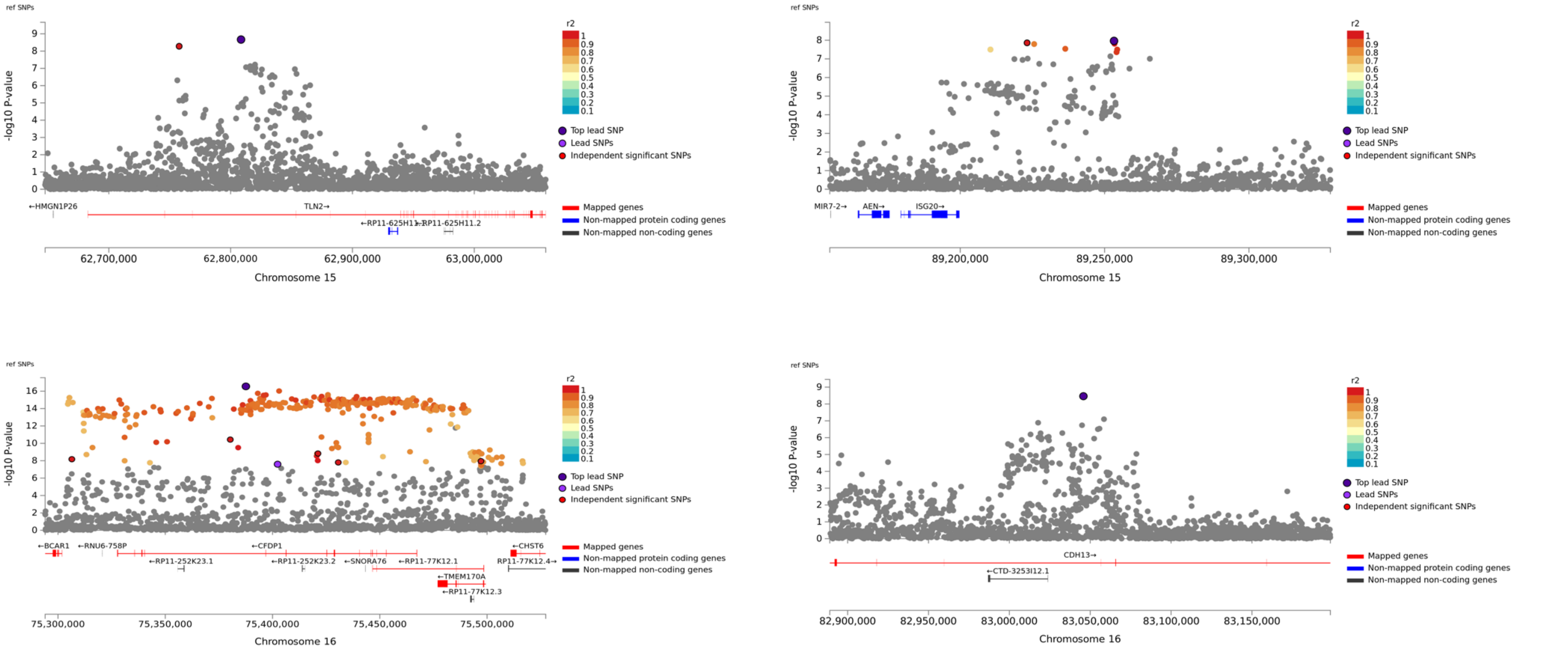
ref SNPs
-log10 P-value
r2
Top lead SNP
Lead SNPs
Independent significant SNPs
Mapped genes
Non-mapped protein coding genes
Non-mapped non-coding genes
←HMGN1P26
TLN2→
62,700,000
62,800,000
62,900,000
63,000,000
Chromosome 15
MIR7-2→
AEN→
ISG20→
89,200,000
89,250,000
89,300,000
Chromosome 15
←BCAR1
←RNU6-758P
←CFDP1
←CHST6
←SNORA76
←TMEM170A
75,300,000
75,350,000
75,400,000
75,450,000
75,500,000
Chromosome 16
CDH13→
82,900,000
82,950,000
83,000,000
83,050,000
83,100,000
83,150,000
Chromosome 16

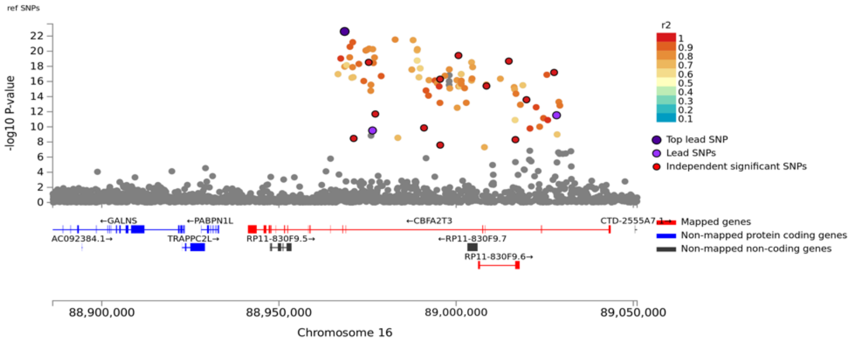
ref SNPs
-log10 P-value
r2
Top lead SNP
Lead SNPs
Independent significant SNPs
←GALNS
←PABPN1L
←CBFA2T3
AC092384.1→
TRAPPC2L→
RP11-830F9.5→
←RP11-830F9.7
RP11-830F9.6→
Mapped genes
Non-mapped protein coding genes
Non-mapped non-coding genes
88,900,000
88,950,000
89,000,000
89,050,000
Chromosome 16

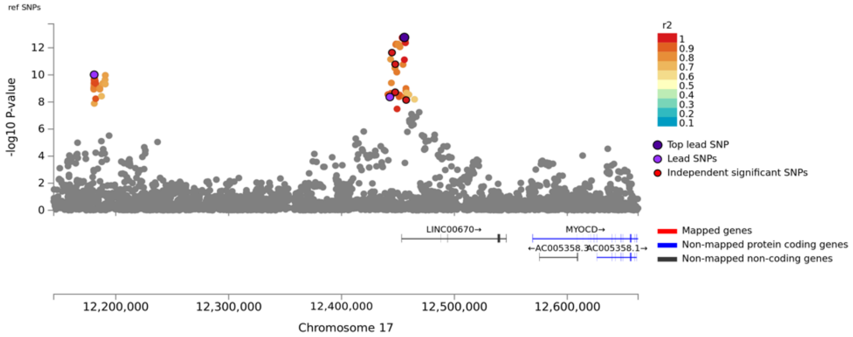
ref SNPs
-log10 P-value
r2
Top lead SNP
Lead SNPs
Independent significant SNPs
LINC00670→
MYOCD→
Mapped genes
Non-mapped protein coding genes
Non-mapped non-coding genes
12,200,000
12,300,000
12,400,000
12,500,000
12,600,000
Chromosome 17

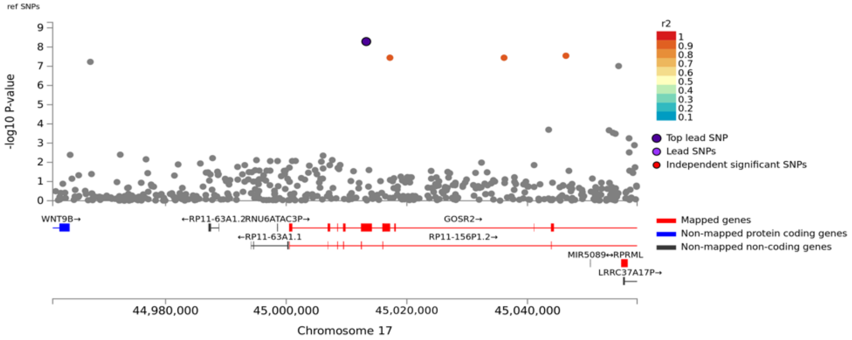
ref SNPs
-log10 P-value
r2
Top lead SNP
Lead SNPs
Independent significant SNPs
WNT9B→
GOSR2→
←RP11-63A1.1
RP11-156P1.2→
Mapped genes
Non-mapped protein coding genes
Non-mapped non-coding genes
44,980,000
45,000,000
45,020,000
45,040,000
Chromosome 17

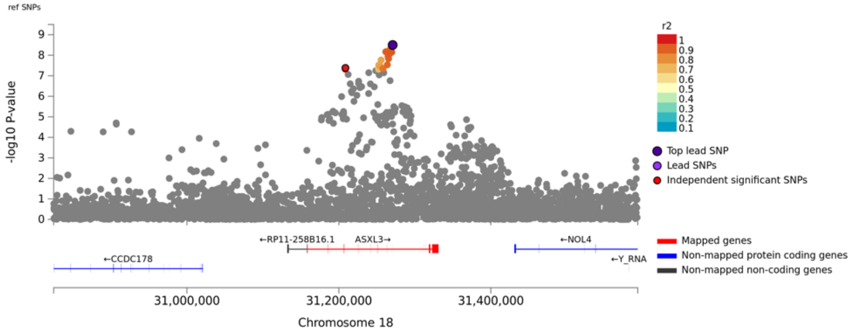
ref SNPs
-log10 P-value
r2
Top lead SNP
Lead SNPs
Independent significant SNPs
←RP11-258B16.1
ASXL3→
←NOL4
←CCDC178
←Y_RNA
Mapped genes
Non-mapped protein coding genes
Non-mapped non-coding genes
31,000,000
31,200,000
31,400,000
Chromosome 18

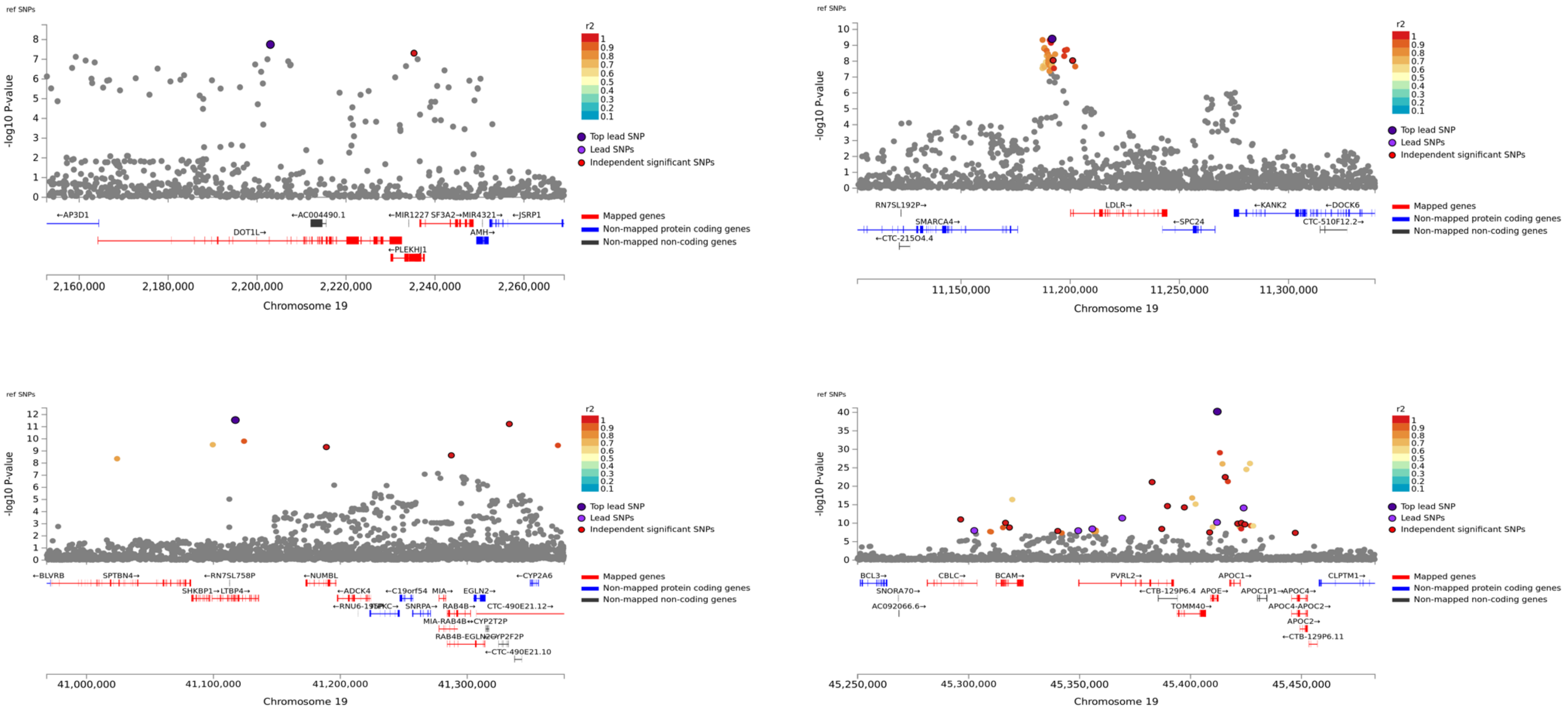
ref SNPs
-log10 P-value
r2
Top lead SNP
Lead SNPs
Independent significant SNPs
Mapped genes
Non-mapped protein coding genes
Non-mapped non-coding genes
←AP3D1
←AC004490.1
←MIR1227 SF3A2→MIR4321→
←JSRP1
DOT1L→
AMH→
←PLEKHJ1
2,160,000
2,180,000
2,200,000
2,220,000
2,240,000
2,260,000
Chromosome 19
RN7SL192P→
LDLR→
←KANK2
←DOCK6
SMARCA4→
←SPC24
CTC-510F12.2→
←CTC-215O4.4
11,150,000
11,200,000
11,250,000
11,300,000
Chromosome 19
←BLVRB
SPTBN4→
←RN7SL758P
←NUMBL
←CYP2A6
SHKBP1→LTBP4→
←ADCK4
←C19orf54
MIA→
EGLN2→
SNRPA→
RAB4B→
CTC-490E21.12→
MIA-RAB4B
←CYP2T2P
←CTC-490E21.10
41,000,000
41,100,000
41,200,000
41,300,000
Chromosome 19
BCL3→
CBLC→
BCAM→
PVRL2→
APOC1→
CLPTM1→
SNORA70→
←CTB-129P6.4
APOE→
APOC1P1→
←APOC4→
AC092066.6→
TOMM40→
APOC4-APOC2→
APOC2→
←CTB-129P6.11
45,250,000
45,300,000
45,350,000
45,400,000
45,450,000
Chromosome 19

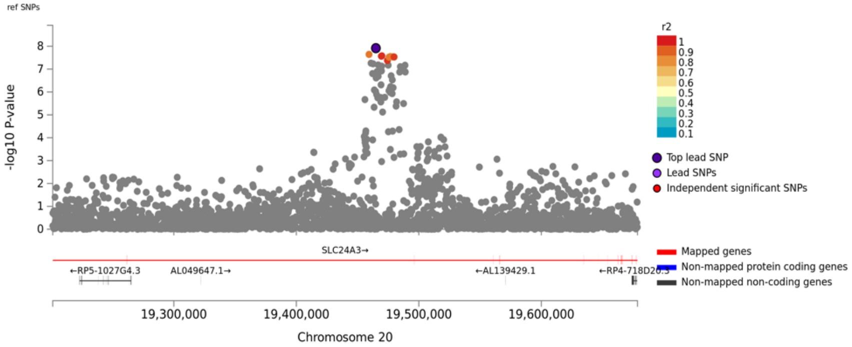

ref SNPs
-log10 P-value
r2
Top lead SNP
Lead SNPs
Independent significant SNPs
SLC24A3→
←RP5-1027G4.3
AL049647.1→
←AL139429.1
Mapped genes
Non-mapped protein coding genes
Non-mapped non-coding genes
19,300,000
19,400,000
19,500,000
19,600,000
Chromosome 20


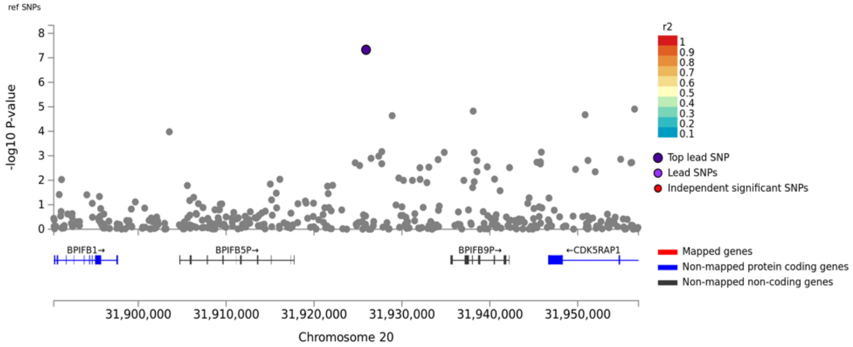

ref SNPs
-log10 P-value
r2
Top lead SNP
Lead SNPs
Independent significant SNPs
BPIFB1→
BPIFB5P→
BPIFB9P→
←CDK5RAP1
Mapped genes
Non-mapped protein coding genes
Non-mapped non-coding genes
31,900,000
31,910,000
31,920,000
31,930,000
31,940,000
31,950,000
Chromosome 20


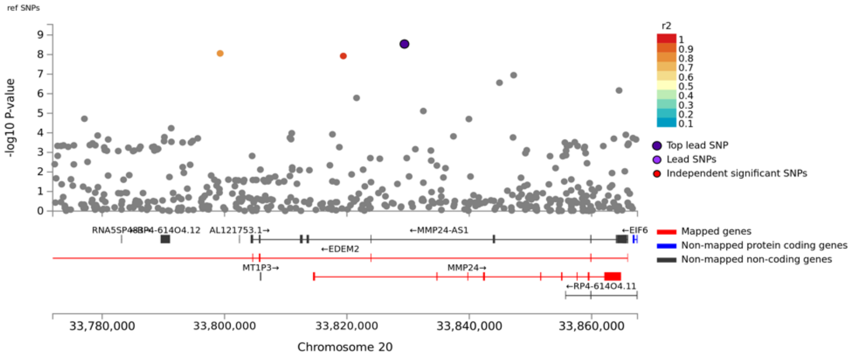

ref SNPs
-log10 P-value
r2
Top lead SNP
Lead SNPs
Independent significant SNPs
AL121753.1→
←MMP24-AS1
←EIF6
←EDEM2
MT1P3→
MMP24→
←RP4-614O4.11
Mapped genes
Non-mapped protein coding genes
Non-mapped non-coding genes
33,780,000
33,800,000
33,820,000
33,840,000
33,860,000
Chromosome 20


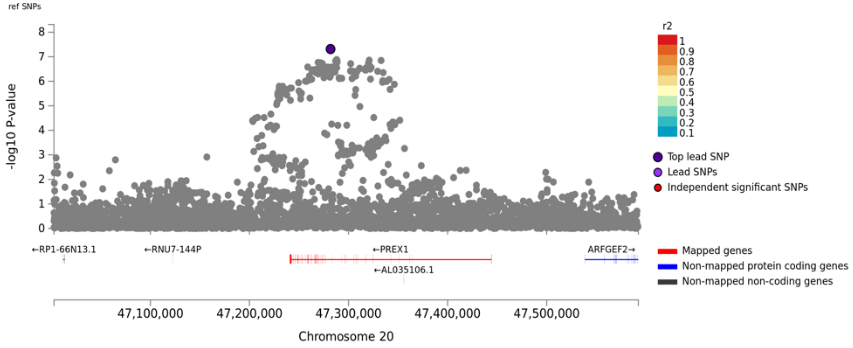

ref SNPs
-log10 P-value
r2
Top lead SNP
Lead SNPs
Independent significant SNPs
←RP1-66N13.1
←RNU7-144P
←PREX1
←AL035106.1
ARFGEF2→
Mapped genes
Non-mapped protein coding genes
Non-mapped non-coding genes
47,100,000
47,200,000
47,300,000
47,400,000
47,500,000
Chromosome 20

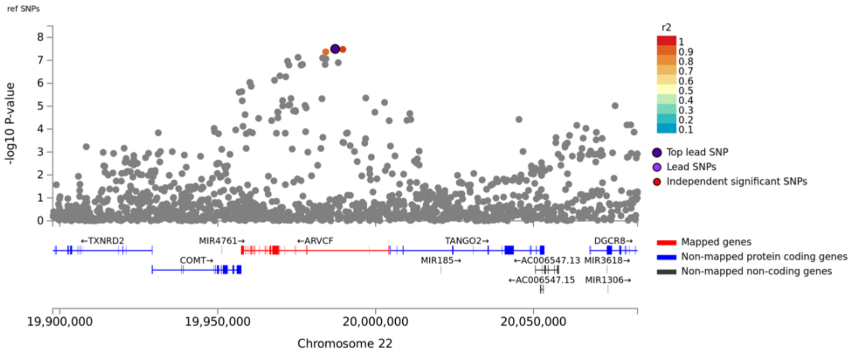
ref SNPs
-log10 P-value
r2
Top lead SNP
Lead SNPs
Independent significant SNPs
←TXNRD2
MIR4761→
←ARVCF
TANGO2→
DGCR8→
COMT→
MIR185→
←AC006547.13
MIR3618→
←AC006547.15
MIR1306→
Mapped genes
Non-mapped protein coding genes
Non-mapped non-coding genes
19,900,000
19,950,000
20,000,000
20,050,000
Chromosome 22

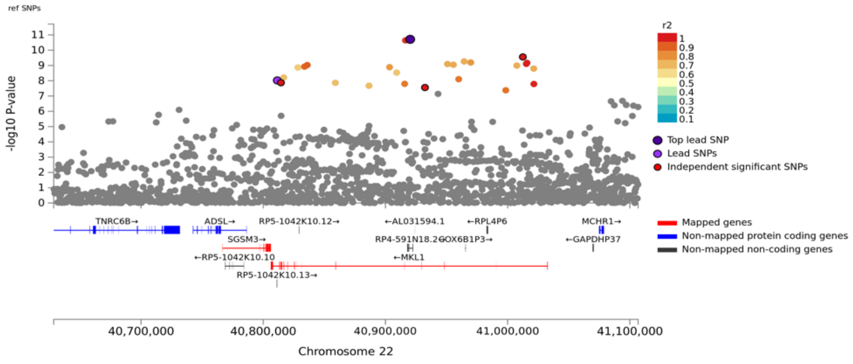
ref SNPs
-log10 P-value
r2
Top lead SNP
Lead SNPs
Independent significant SNPs
TNRC6B→
ADSL→
RP5-1042K10.12→
←AL031594.1
←RPL4P6
MCHR1→
SGSM3→
RP4-591N18.2
COX6B1P3→
←GAPDHP37
←RP5-1042K10.10
←MKL1
RP5-1042K10.13→
Mapped genes
Non-mapped protein coding genes
Non-mapped non-coding genes
40,700,000
40,800,000
40,900,000
41,000,000
41,100,000
Chromosome 22

**Supplementary Figure 5.** Forest plots of 51 loci significantly associated with cIMT

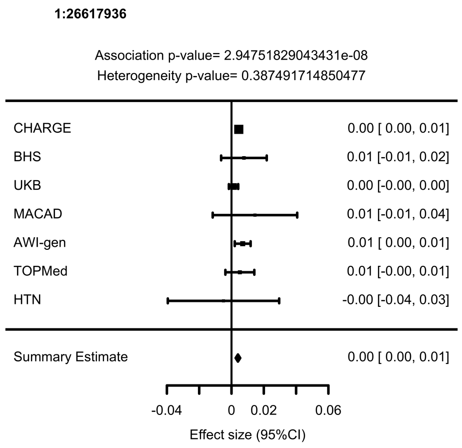


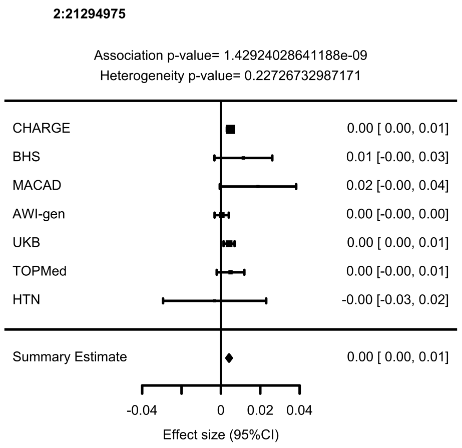


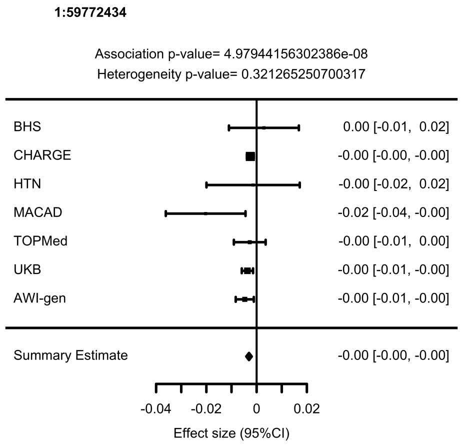


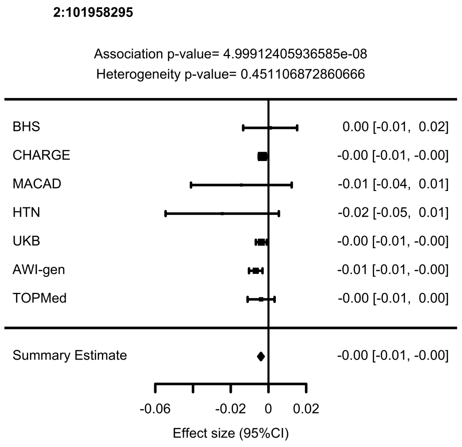

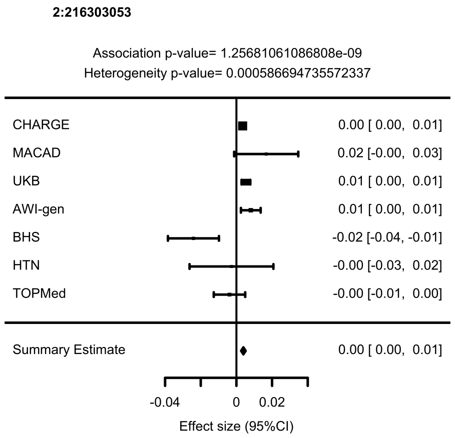
2:216303053
Association p-value= 1.25681061086808e-09
Heterogeneity p-value= 0.000586694735572337
CHARGE 0.00 [ 0.00, 0.01]
MACAD 0.02 [-0.00, 0.03]
UKB 0.01 [ 0.00, 0.01]
AWI-gen 0.01 [ 0.00, 0.01]
BHS -0.02 [-0.04, -0.01]
HTN -0.00 [-0.03, 0.02]
TOPMed -0.00 [-0.01, 0.00]
Summary Estimate 0.00 [ 0.00, 0.01]
-0.04
0
0.02
Effect size (95%CI)

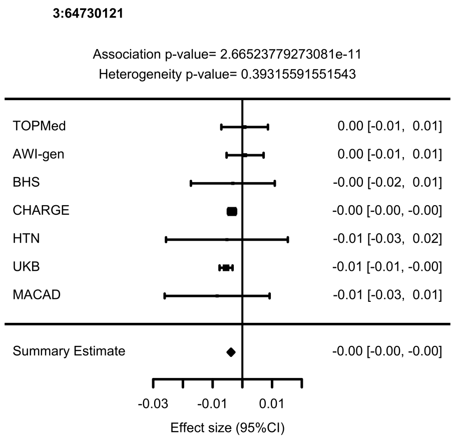
3:64730121
Association p-value= 2.66523779273081e-11
Heterogeneity p-value= 0.39315591551543
TOPMed 0.00 [-0.01, 0.01]
AWI-gen 0.00 [-0.01, 0.01]
BHS -0.00 [-0.02, 0.01]
CHARGE -0.00 [-0.00, -0.00]
HTN -0.01 [-0.03, 0.02]
UKB -0.01 [-0.01, -0.00]
MACAD -0.01 [-0.03, 0.01]
Summary Estimate -0.00 [-0.00, -0.00]
-0.03
-0.01
0.01
Effect size (95%CI)

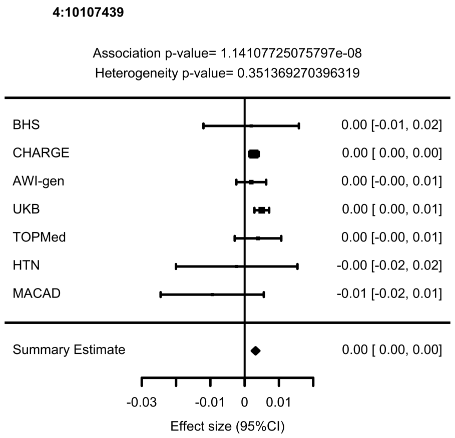
4:10107439
Association p-value= 1.14107725075797e-08
Heterogeneity p-value= 0.351369270396319
BHS 0.00 [-0.01, 0.02]
CHARGE 0.00 [ 0.00, 0.00]
AWI-gen 0.00 [-0.00, 0.01]
UKB 0.00 [ 0.00, 0.01]
TOPMed 0.00 [-0.00, 0.01]
HTN -0.00 [-0.02, 0.02]
MACAD -0.01 [-0.02, 0.01]
Summary Estimate 0.00 [ 0.00, 0.00]
-0.03
-0.01
0
0.01
Effect size (95%CI)

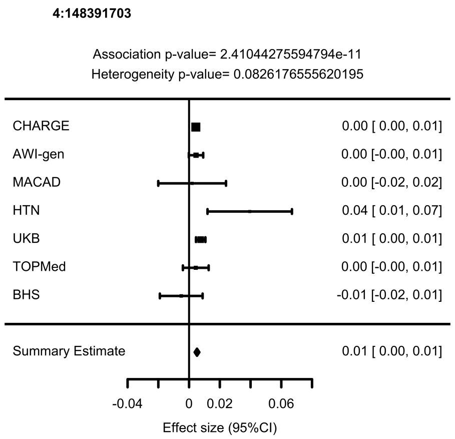
4:148391703
Association p-value= 2.41044275594794e-11
Heterogeneity p-value= 0.0826176555620195
CHARGE 0.00 [ 0.00, 0.01]
AWI-gen 0.00 [-0.00, 0.01]
MACAD 0.00 [-0.02, 0.02]
HTN 0.04 [ 0.01, 0.07]
UKB 0.01 [ 0.00, 0.01]
TOPMed 0.00 [-0.00, 0.01]
BHS -0.01 [-0.02, 0.01]
Summary Estimate 0.01 [ 0.00, 0.01]
-0.04
0
0.02
0.06
Effect size (95%CI)

**4:174656889**

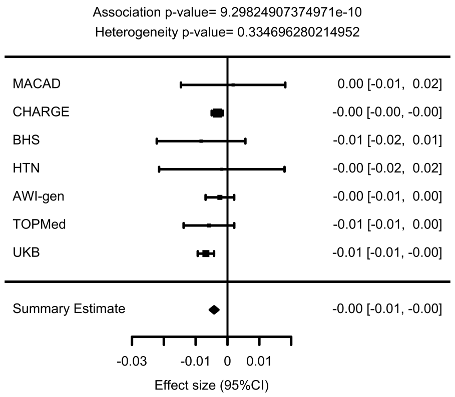


**5:82891343**

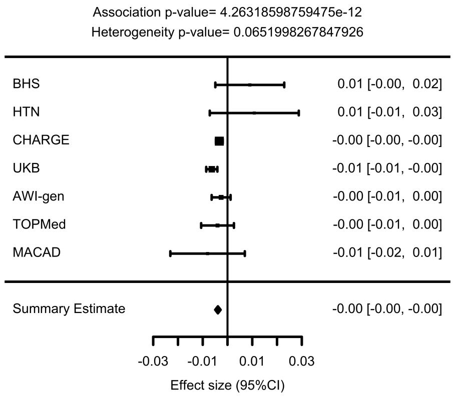


**5:81683739**

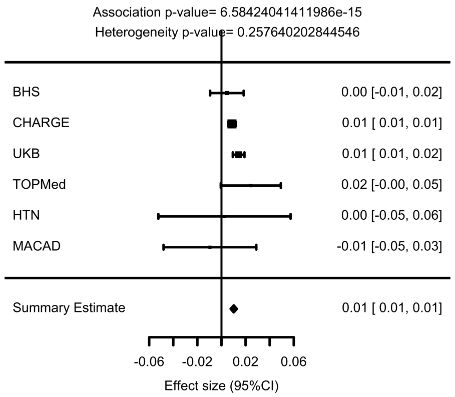


**5:95539330**

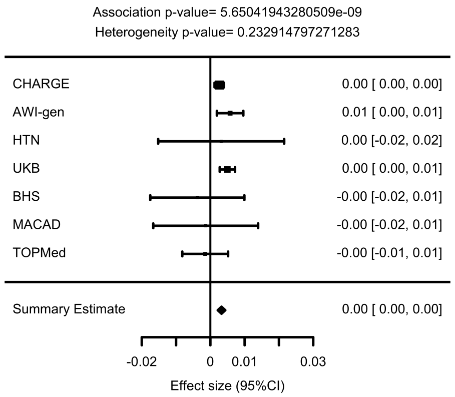

**5:108110324**

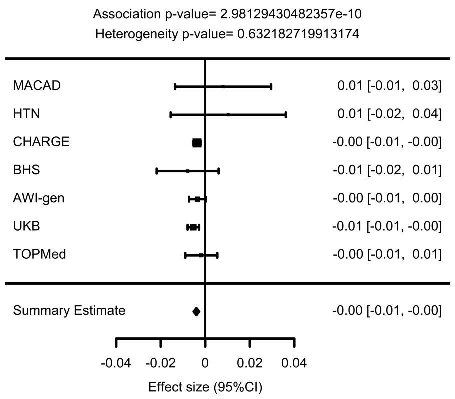


**6:29929253**

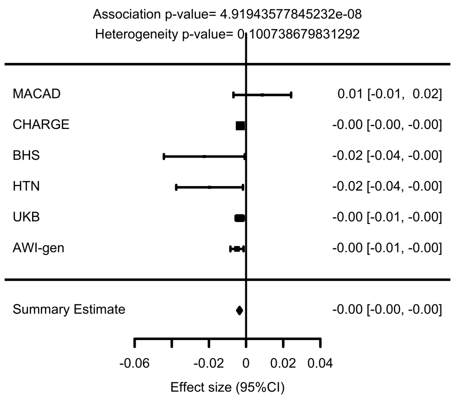


**6:1976723**

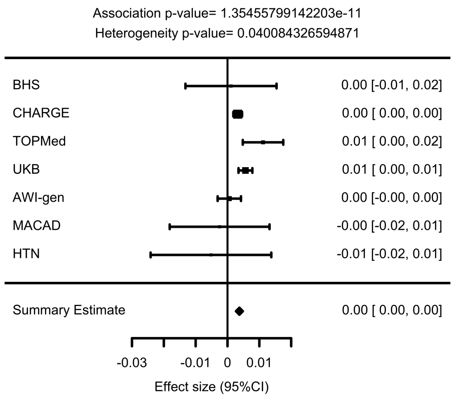


**6:85220342**

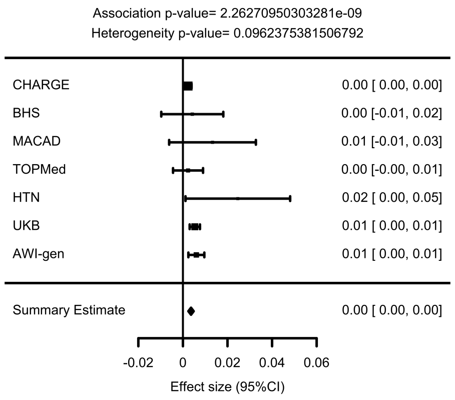

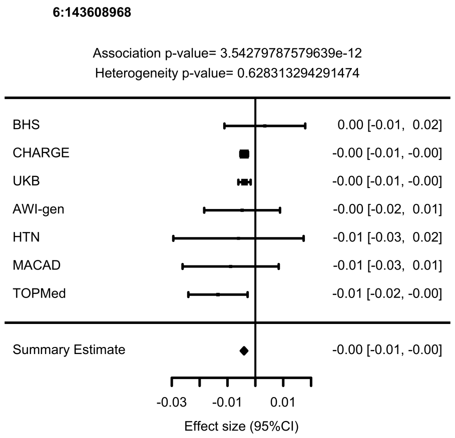
6:143608968
Association p-value= 3.54279787579639e-12
Heterogeneity p-value= 0.628313294291474
BHS 0.00 [-0.01, 0.02]
CHARGE -0.00 [-0.01, -0.00]
UKB -0.00 [-0.01, -0.00]
AWI-gen -0.00 [-0.02, 0.01]
HTN -0.01 [-0.03, 0.02]
MACAD -0.01 [-0.03, 0.01]
TOPMed -0.01 [-0.02, -0.00]
Summary Estimate -0.00 [-0.01, -0.00]
-0.03
-0.01
0.01
Effect size (95%CI)

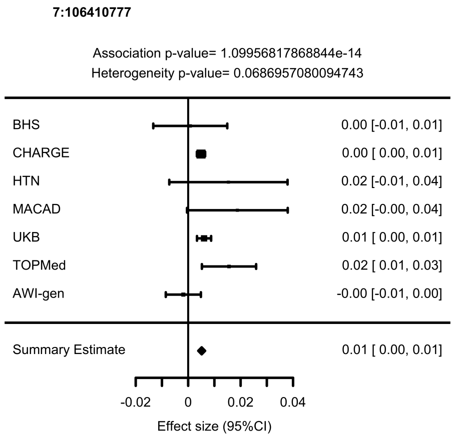
7:106410777
Association p-value= 1.09956817868844e-14
Heterogeneity p-value= 0.0686957080094743
BHS 0.00 [-0.01, 0.01]
CHARGE 0.00 [ 0.00, 0.01]
HTN 0.02 [-0.01, 0.04]
MACAD 0.02 [-0.00, 0.04]
UKB 0.01 [ 0.00, 0.01]
TOPMed 0.02 [ 0.01, 0.03]
AWI-gen -0.00 [-0.01, 0.00]
Summary Estimate 0.01 [ 0.00, 0.01]
-0.02
0
0.02
0.04
Effect size (95%CI)

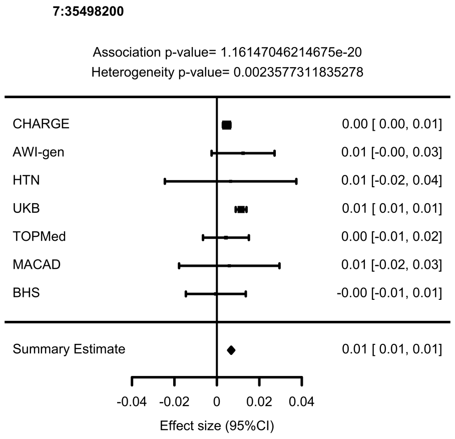
7:35498200
Association p-value= 1.16147046214675e-20
Heterogeneity p-value= 0.0023577311835278
CHARGE 0.00 [ 0.00, 0.01]
AWI-gen 0.01 [-0.00, 0.03]
HTN 0.01 [-0.02, 0.04]
UKB 0.01 [ 0.01, 0.01]
TOPMed 0.00 [-0.01, 0.02]
MACAD 0.01 [-0.02, 0.03]
BHS -0.00 [-0.01, 0.01]
Summary Estimate 0.01 [ 0.01, 0.01]
-0.04
-0.02
0
0.02
0.04
Effect size (95%CI)

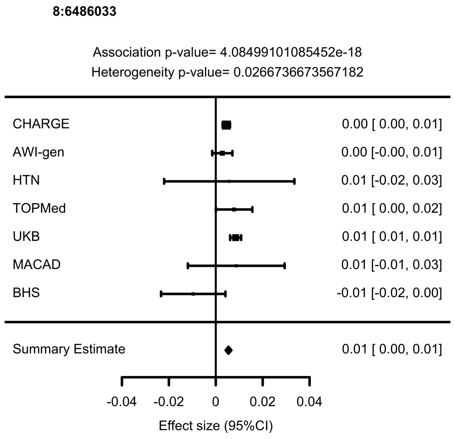
8:6486033
Association p-value= 4.08499101085452e-18
Heterogeneity p-value= 0.0266736673567182
CHARGE 0.00 [ 0.00, 0.01]
AWI-gen 0.00 [-0.00, 0.01]
HTN 0.01 [-0.02, 0.03]
TOPMed 0.01 [ 0.00, 0.02]
UKB 0.01 [ 0.01, 0.01]
MACAD 0.01 [-0.01, 0.03]
BHS -0.01 [-0.02, 0.00]
Summary Estimate 0.01 [ 0.00, 0.01]
-0.04
-0.02
0
0.02
0.04
Effect size (95%CI)

**8:10634132**

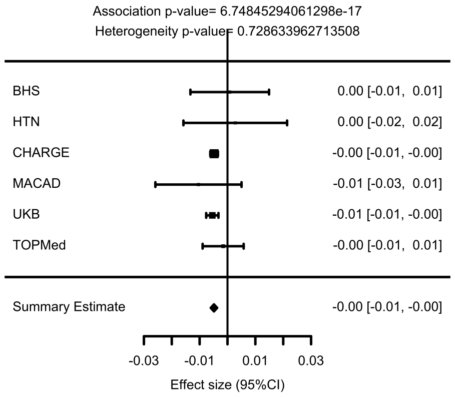


**8:123394850**

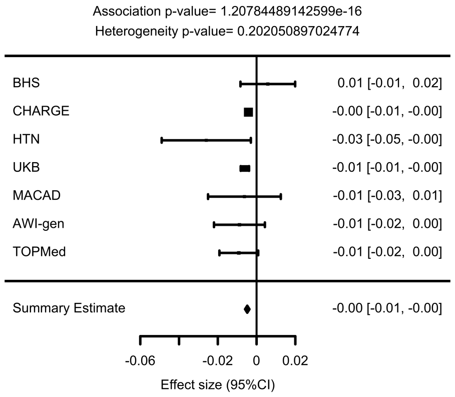


**8:81267236**

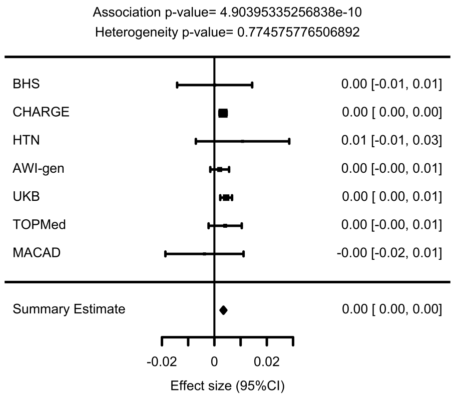


**8:124608614**

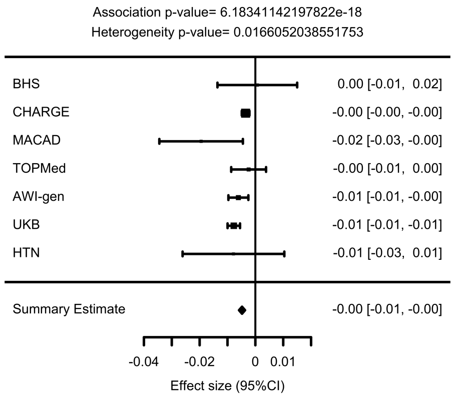

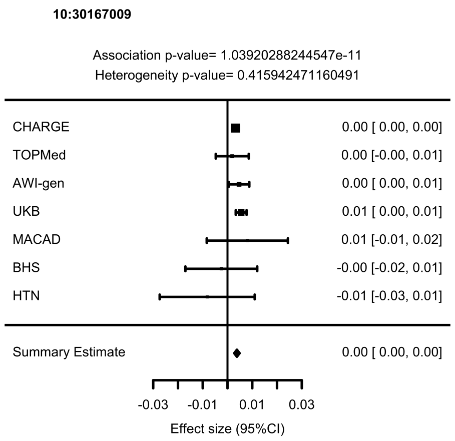
10:30167009
Association p-value= 1.03920288244547e-11
Heterogeneity p-value= 0.415942471160491
CHARGE 0.00 [ 0.00, 0.00]
TOPMed 0.00 [-0.00, 0.01]
AWI-gen 0.00 [ 0.00, 0.01]
UKB 0.01 [ 0.00, 0.01]
MACAD 0.01 [-0.01, 0.02]
BHS -0.00 [-0.02, 0.01]
HTN -0.01 [-0.03, 0.01]
Summary Estimate 0.00 [ 0.00, 0.00]
-0.03 -0.01 0.01 0.03
Effect size (95%CI)

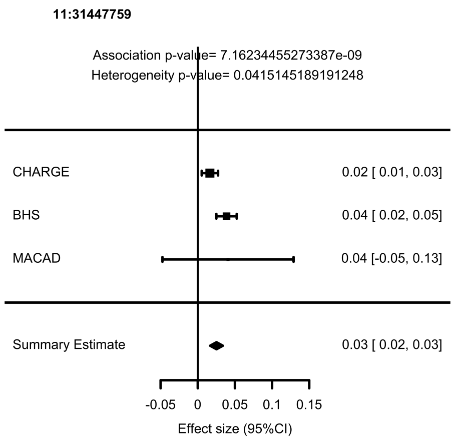
11:31447759
Association p-value= 7.16234455273387e-09
Heterogeneity p-value= 0.0415145189191248
CHARGE 0.02 [ 0.01, 0.03]
BHS 0.04 [ 0.02, 0.05]
MACAD 0.04 [-0.05, 0.13]
Summary Estimate 0.03 [ 0.02, 0.03]
-0.05 0 0.05 0.1 0.15
Effect size (95%CI)

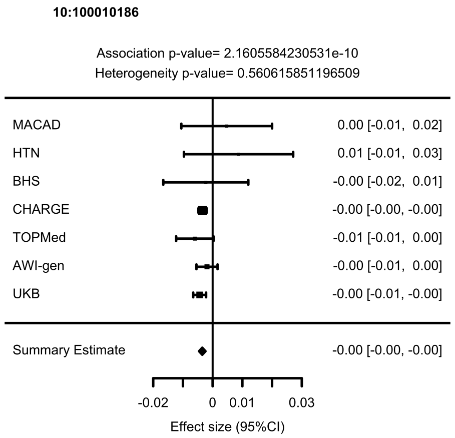
10:100010186
Association p-value= 2.1605584230531e-10
Heterogeneity p-value= 0.560615851196509
MACAD 0.00 [-0.01, 0.02]
HTN 0.01 [-0.01, 0.03]
BHS -0.00 [-0.02, 0.01]
CHARGE -0.00 [-0.00, -0.00]
TOPMed -0.01 [-0.01, 0.00]
AWI-gen -0.00 [-0.01, 0.00]
UKB -0.00 [-0.01, -0.00]
Summary Estimate -0.00 [-0.00, -0.00]
-0.02 0 0.01 0.03
Effect size (95%CI)

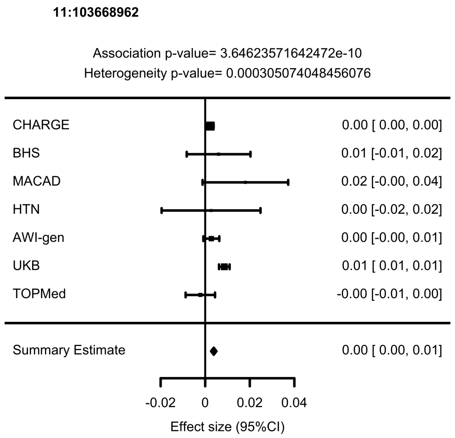
11:103668962
Association p-value= 3.64623571642472e-10
Heterogeneity p-value= 0.000305074048456076
CHARGE 0.00 [ 0.00, 0.00]
BHS 0.01 [-0.01, 0.02]
MACAD 0.02 [-0.00, 0.04]
HTN 0.00 [-0.02, 0.02]
AWI-gen 0.00 [-0.00, 0.01]
UKB 0.01 [ 0.01, 0.01]
TOPMed -0.00 [-0.01, 0.00]
Summary Estimate 0.00 [ 0.00, 0.01]
-0.02 0 0.02 0.04
Effect size (95%CI)

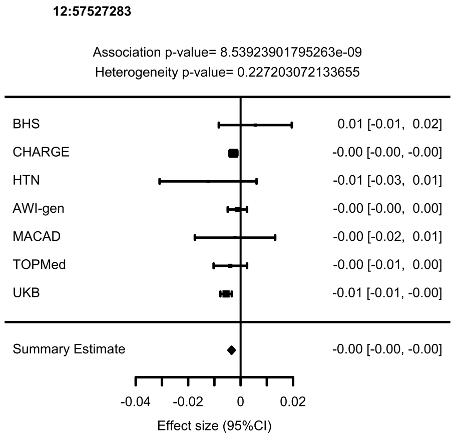
12:57527283
Association p-value= 8.53923901795263e-09
Heterogeneity p-value= 0.227203072133655
BHS 0.01 [-0.01, 0.02]
CHARGE -0.00 [-0.00, -0.00]
HTN -0.01 [-0.03, 0.01]
AWI-gen -0.00 [-0.00, 0.00]
MACAD -0.00 [-0.02, 0.01]
TOPMed -0.00 [-0.01, 0.00]
UKB -0.01 [-0.01, -0.00]
Summary Estimate -0.00 [-0.00, -0.00]
-0.04 -0.02 0 0.02
Effect size (95%CI)

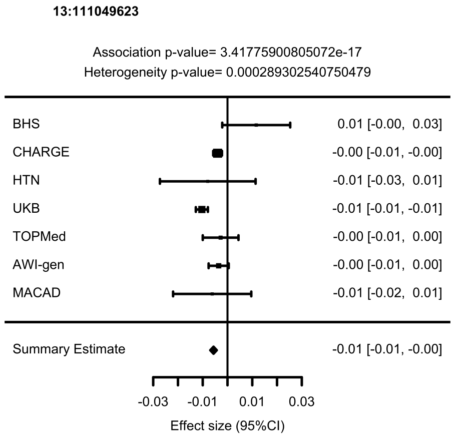
13:111049623
Association p-value= 3.41775900805072e-17
Heterogeneity p-value= 0.000289302540750479
BHS 0.01 [-0.00, 0.03]
CHARGE -0.00 [-0.01, -0.00]
HTN -0.01 [-0.03, 0.01]
UKB -0.01 [-0.01, -0.01]
TOPMed -0.00 [-0.01, 0.00]
AWI-gen -0.00 [-0.01, 0.00]
MACAD -0.01 [-0.02, 0.01]
Summary Estimate -0.01 [-0.01, -0.00]
-0.03 -0.01 0.01 0.03
Effect size (95%CI)

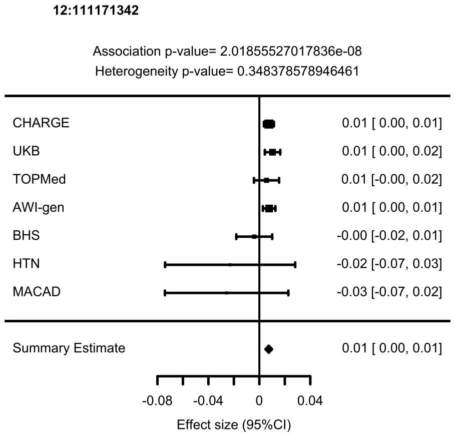
12:111171342
Association p-value= 2.01855527017836e-08
Heterogeneity p-value= 0.348378578946461
CHARGE 0.01 [ 0.00, 0.01]
UKB 0.01 [ 0.00, 0.02]
TOPMed 0.01 [-0.00, 0.02]
AWI-gen 0.01 [ 0.00, 0.01]
BHS -0.00 [-0.02, 0.01]
HTN -0.02 [-0.07, 0.03]
MACAD -0.03 [-0.07, 0.02]
Summary Estimate 0.01 [ 0.00, 0.01]
-0.08 -0.04 0 0.04
Effect size (95%CI)

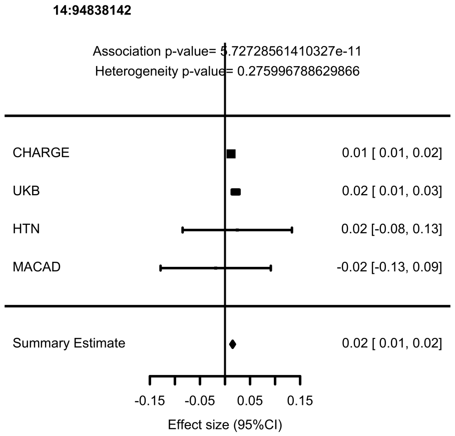
14:94838142
Association p-value= 5.72728561410327e-11
Heterogeneity p-value= 0.275996788629866
CHARGE 0.01 [ 0.01, 0.02]
UKB 0.02 [ 0.01, 0.03]
HTN 0.02 [-0.08, 0.13]
MACAD -0.02 [-0.13, 0.09]
Summary Estimate 0.02 [ 0.01, 0.02]
-0.15 -0.05 0.05 0.15
Effect size (95%CI)

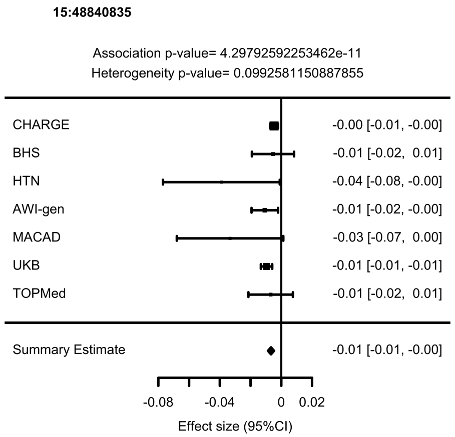
15:48840835
Association p-value= 4.29792592253462e-11
Heterogeneity p-value= 0.0992581150887855
CHARGE -0.00 [-0.01, -0.00]
BHS -0.01 [-0.02, 0.01]
HTN -0.04 [-0.08, -0.00]
AWI-gen -0.01 [-0.02, -0.00]
MACAD -0.03 [-0.07, 0.00]
UKB -0.01 [-0.01, -0.01]
TOPMed -0.01 [-0.02, 0.01]
Summary Estimate -0.01 [-0.01, -0.00]
-0.08 -0.04 0 0.02
Effect size (95%CI)

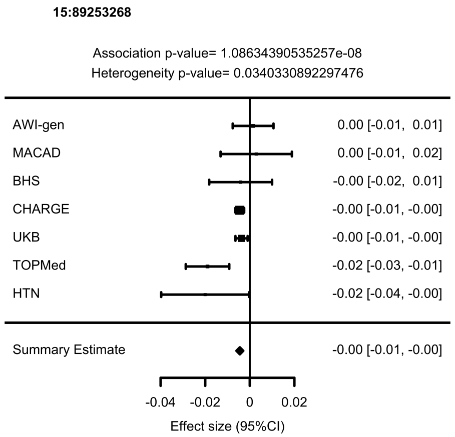
15:89253268
Association p-value= 1.08634390535257e-08
Heterogeneity p-value= 0.0340330892297476
AWI-gen 0.00 [-0.01, 0.01]
MACAD 0.00 [-0.01, 0.02]
BHS -0.00 [-0.02, 0.01]
CHARGE -0.00 [-0.01, -0.00]
UKB -0.00 [-0.01, -0.00]
TOPMed -0.02 [-0.03, -0.01]
HTN -0.02 [-0.04, -0.00]
Summary Estimate -0.00 [-0.01, -0.00]
-0.04 -0.02 0 0.02
Effect size (95%CI)

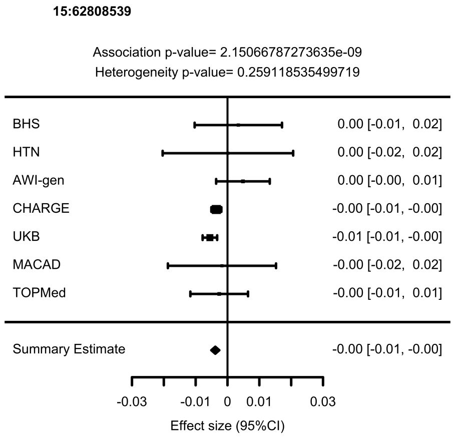
15:62808539
Association p-value= 2.15066787273635e-09
Heterogeneity p-value= 0.259118535499719
BHS 0.00 [-0.01, 0.02]
HTN 0.00 [-0.02, 0.02]
AWI-gen 0.00 [-0.00, 0.01]
CHARGE -0.00 [-0.01, -0.00]
UKB -0.01 [-0.01, -0.00]
MACAD -0.00 [-0.02, 0.02]
TOPMed -0.00 [-0.01, 0.01]
Summary Estimate -0.00 [-0.01, -0.00]
-0.03 -0.01 0 0.01 0.03
Effect size (95%CI)

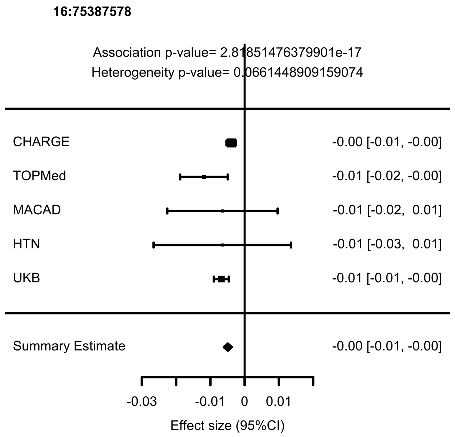
16:75387578
Association p-value= 2.81851476379901e-17
Heterogeneity p-value= 0.0661448909159074
CHARGE -0.00 [-0.01, -0.00]
TOPMed -0.01 [-0.02, -0.00]
MACAD -0.01 [-0.02, 0.01]
HTN -0.01 [-0.03, 0.01]
UKB -0.01 [-0.01, -0.00]
Summary Estimate -0.00 [-0.01, -0.00]
-0.03 -0.01 0 0.01
Effect size (95%CI)

**16:83045790**

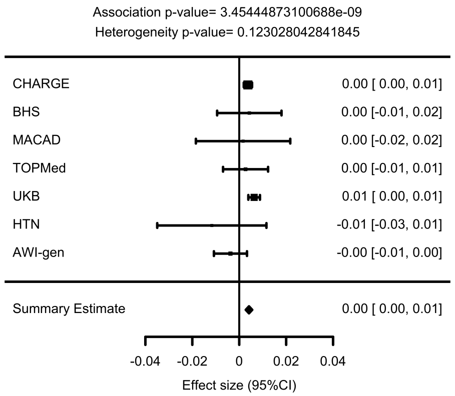


**16:88968540**

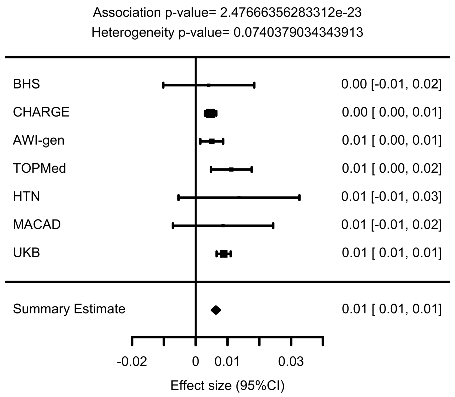


**17:12455692**

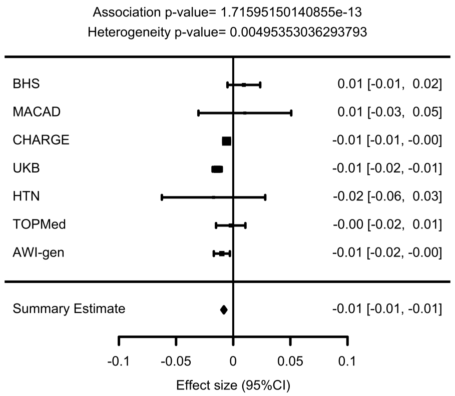


**17:45013271**

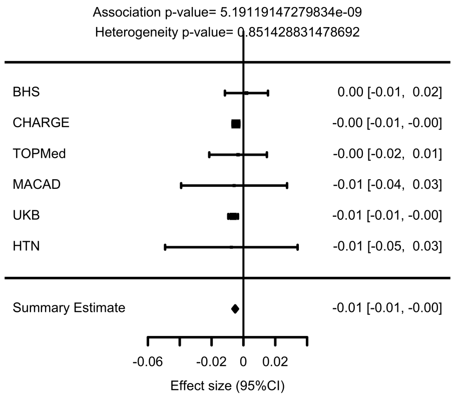

18:31270819

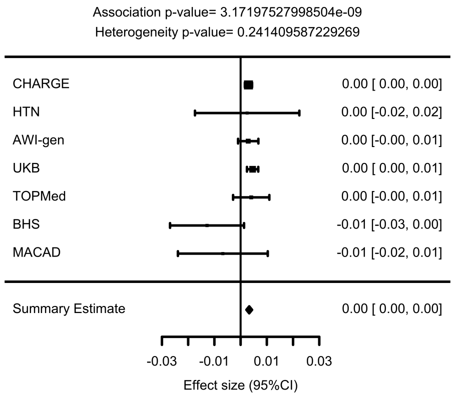


19:11191729

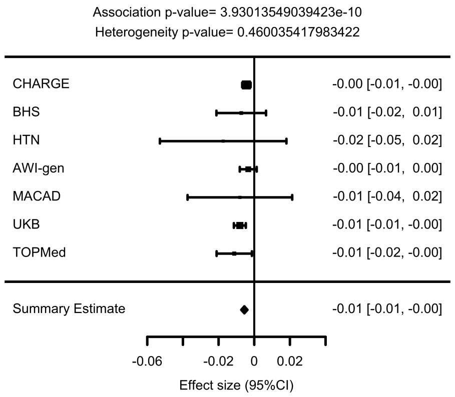


19:2202992

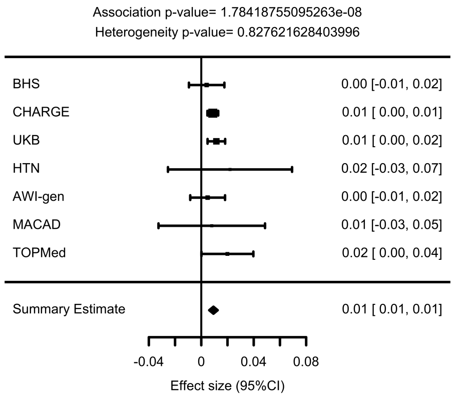


19:41117300

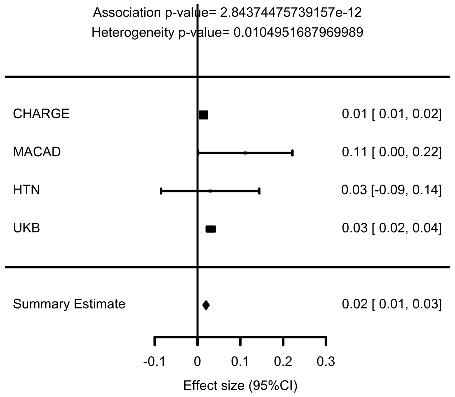

**19:45412079**

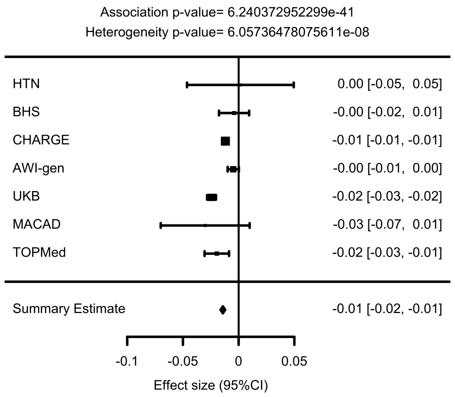


**20:31925918**

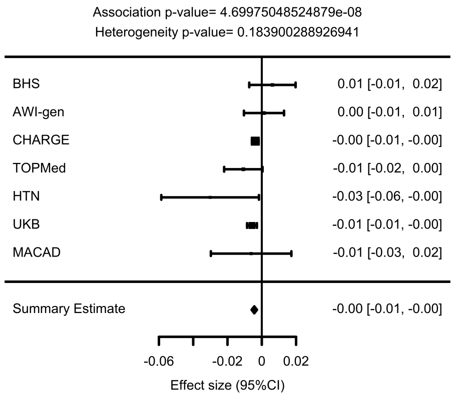


**20:19464926**

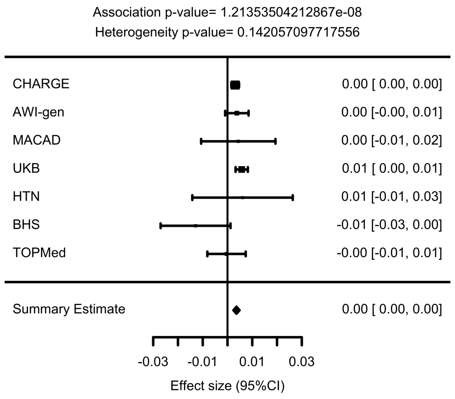


**20:33829406**

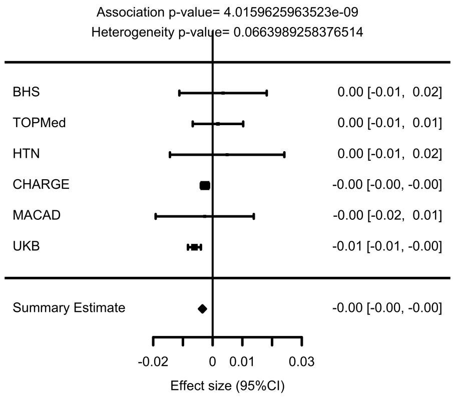

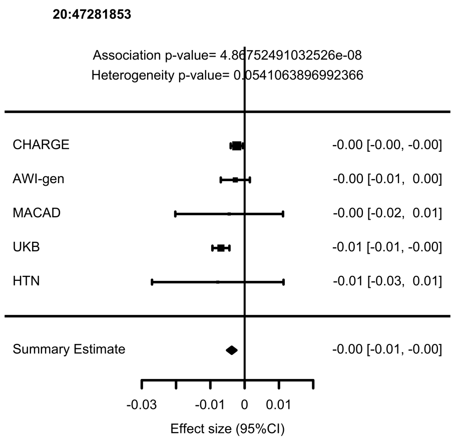
20:47281853
Association p-value= 4.86752491032526e-08
Heterogeneity p-value= 0.0541063896992366
CHARGE -0.00 [-0.00, -0.00]
AWI-gen -0.00 [-0.01, 0.00]
MACAD -0.00 [-0.02, 0.01]
UKB -0.01 [-0.01, -0.00]
HTN -0.01 [-0.03, 0.01]
Summary Estimate -0.00 [-0.01, -0.00]
-0.03
-0.01
0
0.01
Effect size (95%CI)

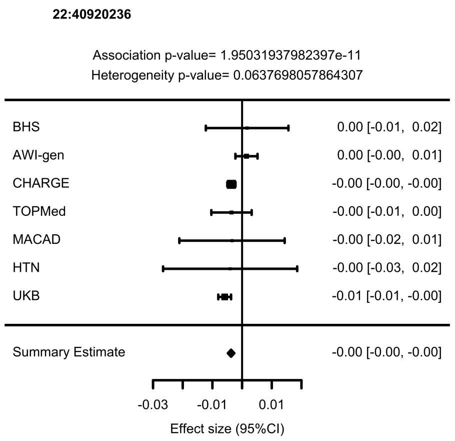
22:40920236
Association p-value= 1.95031937982397e-11
Heterogeneity p-value= 0.0637698057864307
BHS 0.00 [-0.01, 0.02]
AWI-gen 0.00 [-0.00, 0.01]
CHARGE -0.00 [-0.00, -0.00]
TOPMed -0.00 [-0.01, 0.00]
MACAD -0.00 [-0.02, 0.01]
HTN -0.00 [-0.03, 0.02]
UKB -0.01 [-0.01, -0.00]
Summary Estimate -0.00 [-0.00, -0.00]
-0.03
-0.01
0.01
Effect size (95%CI)

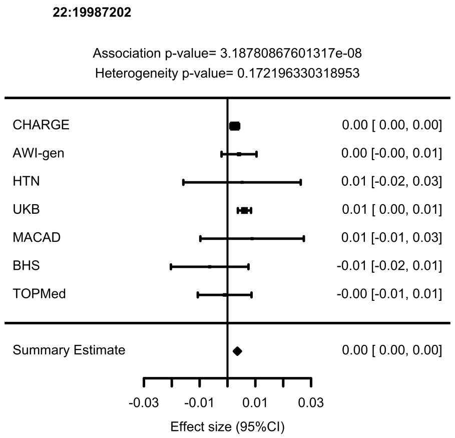
22:19987202
Association p-value= 3.18780867601317e-08
Heterogeneity p-value= 0.172196330318953
CHARGE 0.00 [ 0.00, 0.00]
AWI-gen 0.00 [-0.00, 0.01]
HTN 0.01 [-0.02, 0.03]
UKB 0.01 [ 0.00, 0.01]
MACAD 0.01 [-0.01, 0.03]
BHS -0.01 [-0.02, 0.01]
TOPMed -0.00 [-0.01, 0.01]
Summary Estimate 0.00 [ 0.00, 0.00]
-0.03
-0.01
0.01
0.03
Effect size (95%CI)

**Supplementary Figure 6**. Circos plots showing cIMT genomic risk loci from the multi-ancestry metanalysis and genes prioritised by *eQTL* and *chromatin interaction.* The dark blue areas indicate cIMT genomic risk loci. Genes in green, orange and red were mapped by *eQTL*, *chromatin interactions*, and both, respectively.

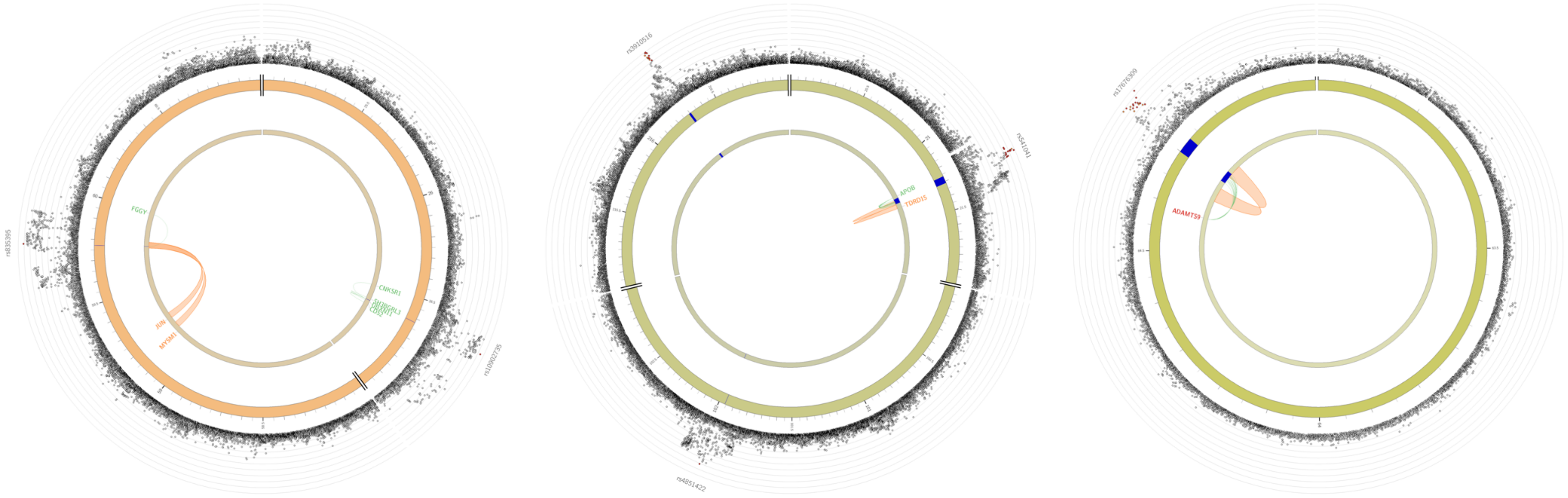

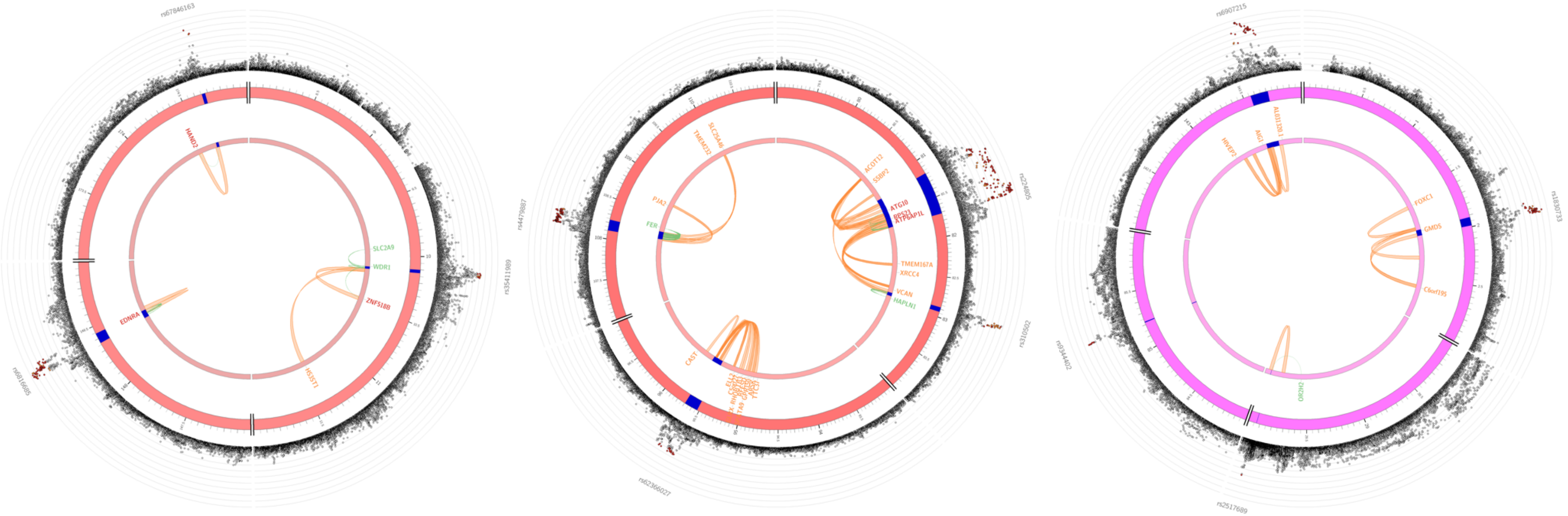
HAND2
SLC2A9
WDR1
ZNF518B
EDNRA
HS3ST1
SLC25A46
TMEM232
PJA2
FER
CAST
ACOT12
SSBP2
ATG10
RPS23
ATP6AP1L
TMEM167A
XRCC4
VCAN
HAPLN1
FOXC1
GMDS
C6orf195

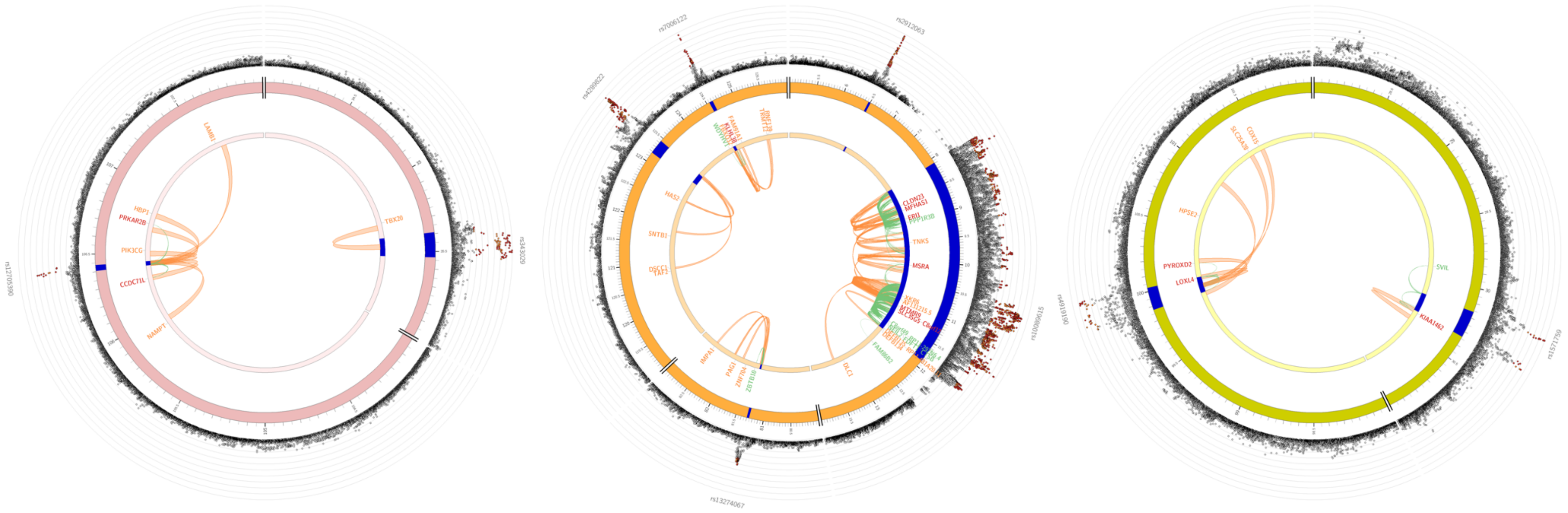

LAMB1
HBP1
PRKAR2B
PIK3CG
CCDC71L
NAMPT
TBX20
HAS2
SNTB1
CLDN23
TNKS
MSRA
IMPA1
DLC1
FAM86B2
HPSE2
PYROXD2
LOXL4
SVIL
KIAA1462

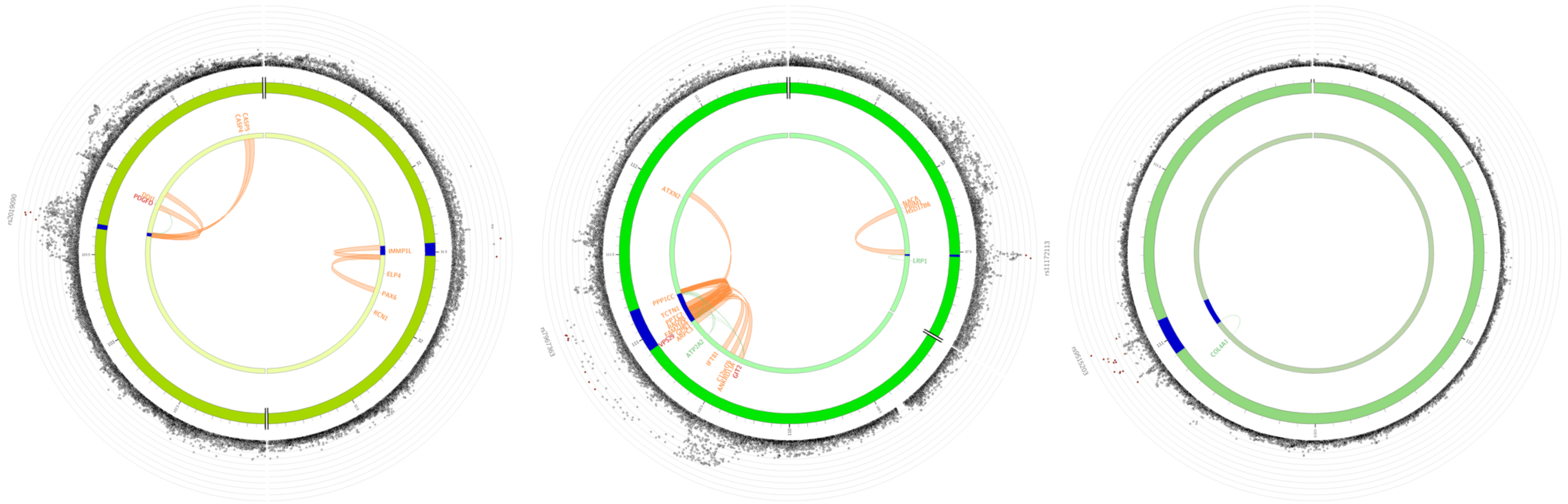
rs2019090
CASP5
CASP4
DDI1
PDGFD
IMMP1L
ELP4
PAX6
RCN1
rs7967363
ATXN2
PPP1CC
TCTN1
VPS29
ATP2A2
IFT81
GIT2
NACA
HSD17B6
LRP1
rs11172113
rs9515203
COL1A1

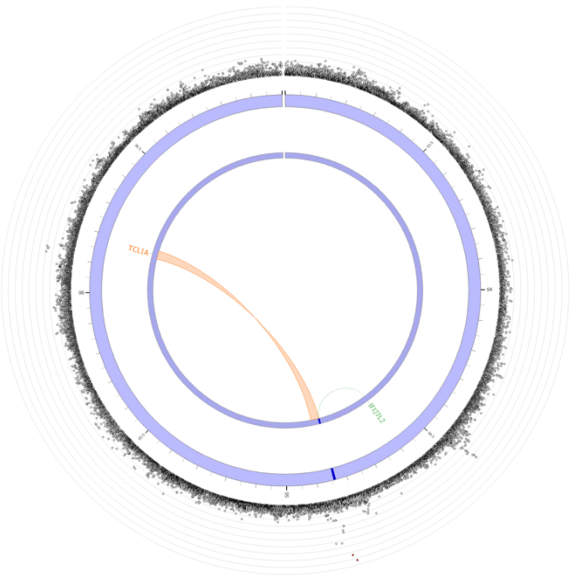
TCL1A

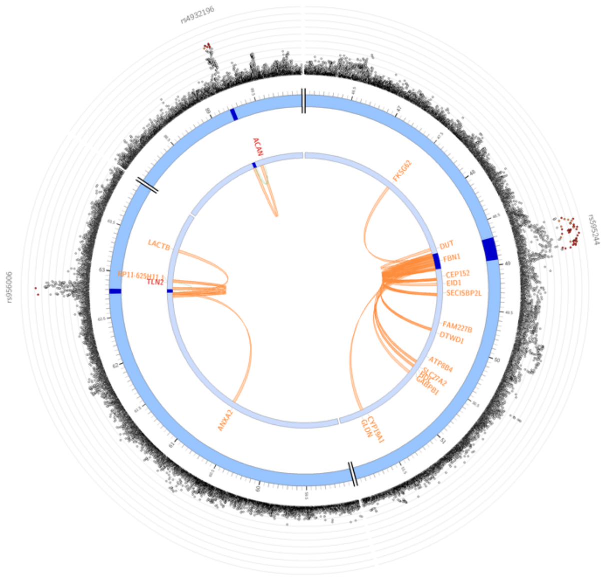
ACAN
LACTB
ANXA2
DUT
FBN1
CEP152
EID1
SECISBP2L
FAM227B
DTWD1
ATP8B4

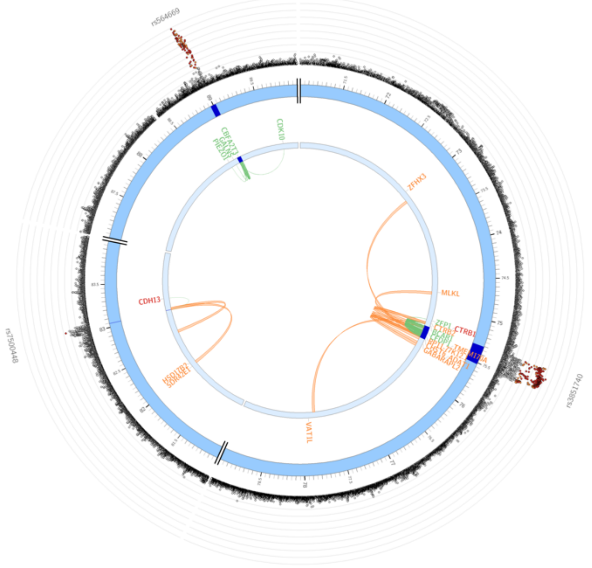
CDK10
CDH13
MLKL
VAT1L

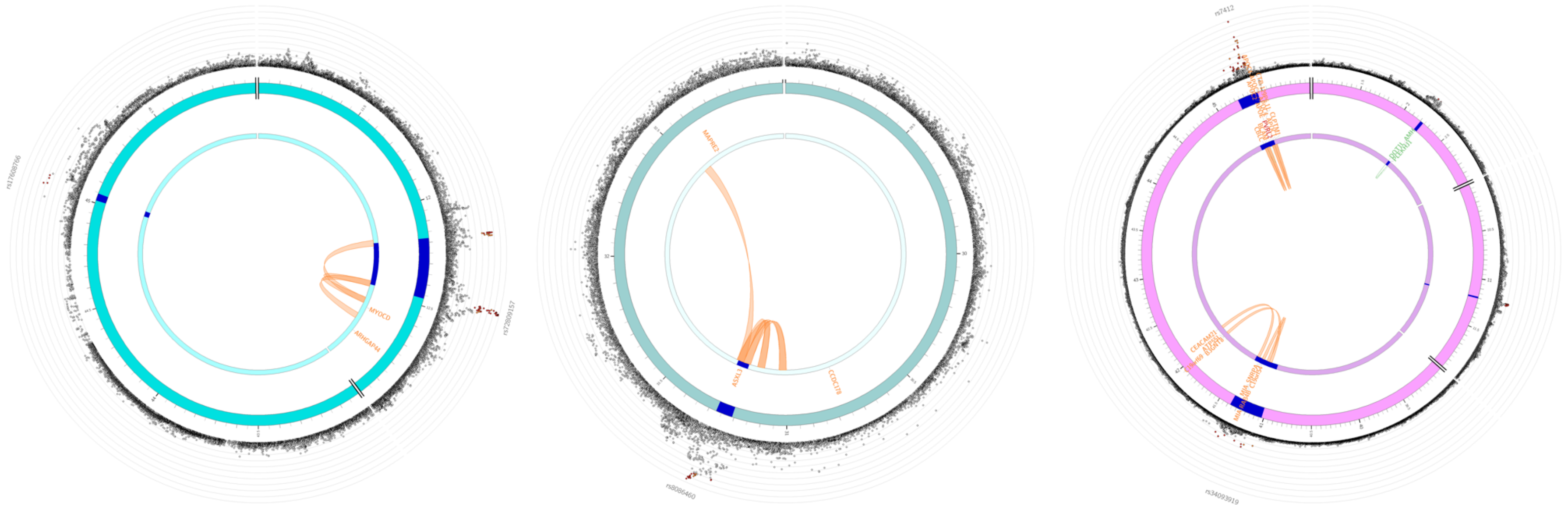
rs17608766
rs72809157
MYOCD
ARHGAP44
MAPRE2
ASXL1
CCDC178
rs8086460
rs7412
rs34093919

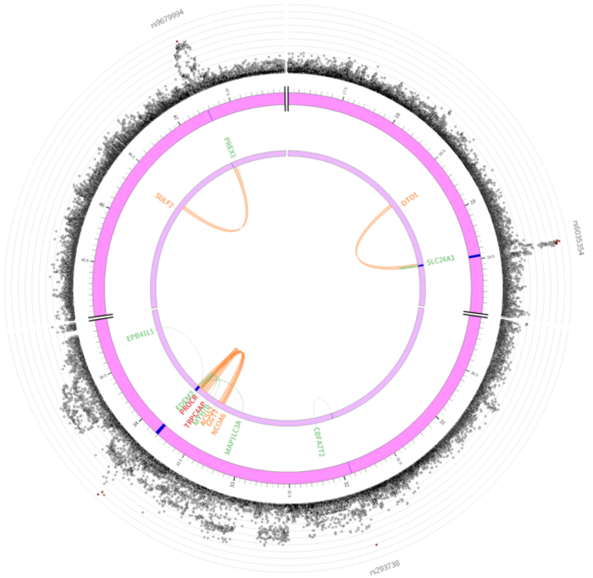
SULF2
PREX1
SLC24A3
EPB41L1
MAP1LC3A
CBFA2T2

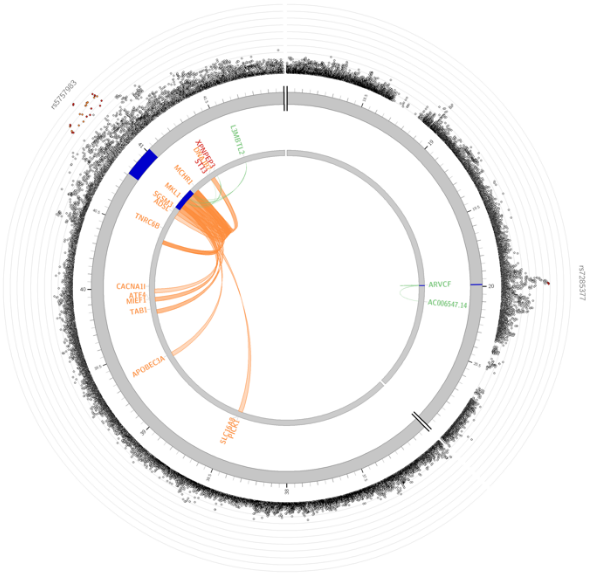
L3MBTL2
TNRC6B
CACNA1I
TAB1
APOBEC3A
ARVCF

**Supplementary Figure 7**. Miami plots for GWAS of cIMT in UK Biobank **a)** $IMT_{mean\text{-}max}$; **b)** $IM_{mean}$; **c)** $IMT_{mean\text{-}max}$ (women); and **d)** $IMT_{mean\text{-}max}$ (men). The plots show p-values ($-log_{10}$ transformed) presented on the y-axis and chromosomal position on the x- axis. The horizontal blue and red lines indicate the threshold for suggestive ($P<1\times10^{-5}$) and genome-wide ($P<5\times10^{-8}$) significance, respectively.

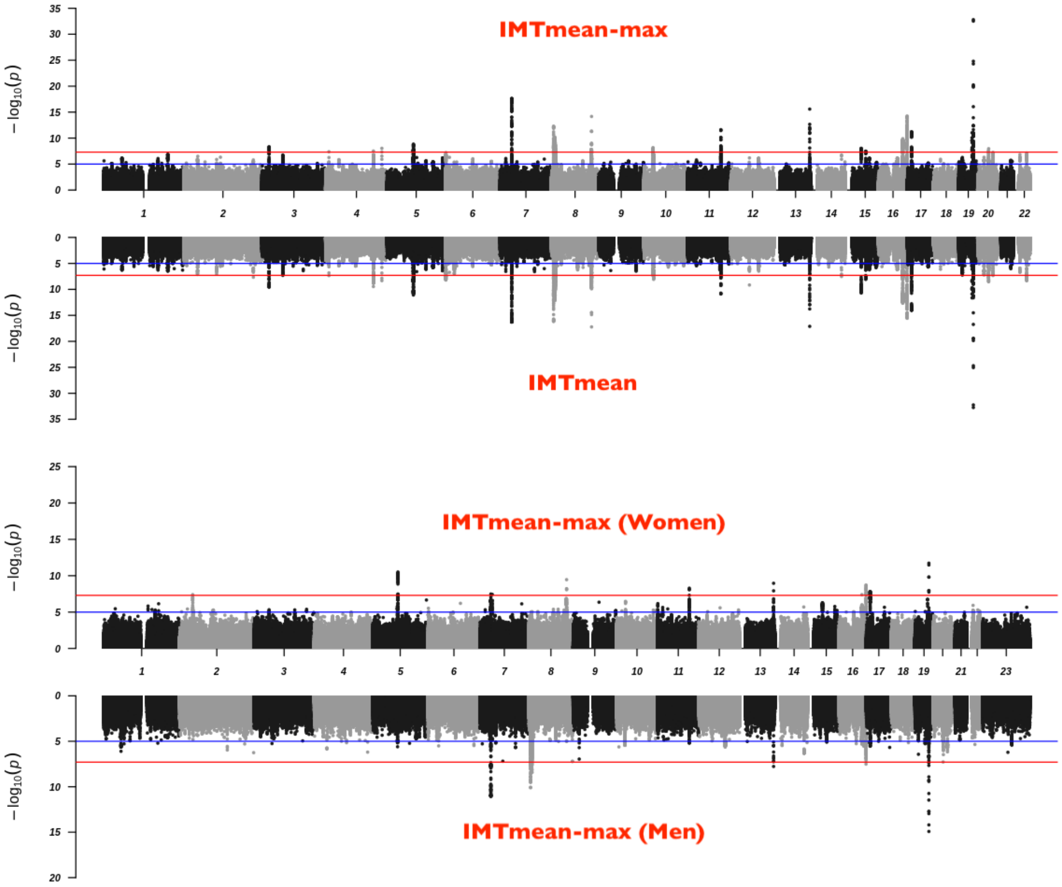

**Supplementary Figure 8.** Comparison of effect size estimates (beta) and p-values from the UKBB cIMT GWAS. **a)** effect size estimates from $IMT_{mean}$ and $IMT_{mean\text{-}max}$ GWAS, **b)** p-values ($-log_{10}$ transformed) from $IMT_{mean}$ and $IMT_{mean\text{-}max}$ GWAS, **c)** effect size estimates from sex-stratified $IMT_{mean\text{-}max}$ GWAS, and **d)** p-values ($-log_{10}$ transformed) from sex-stratified $IMT_{mean\text{-}max}$ GWAS. r, Pearson correlation coefficient.

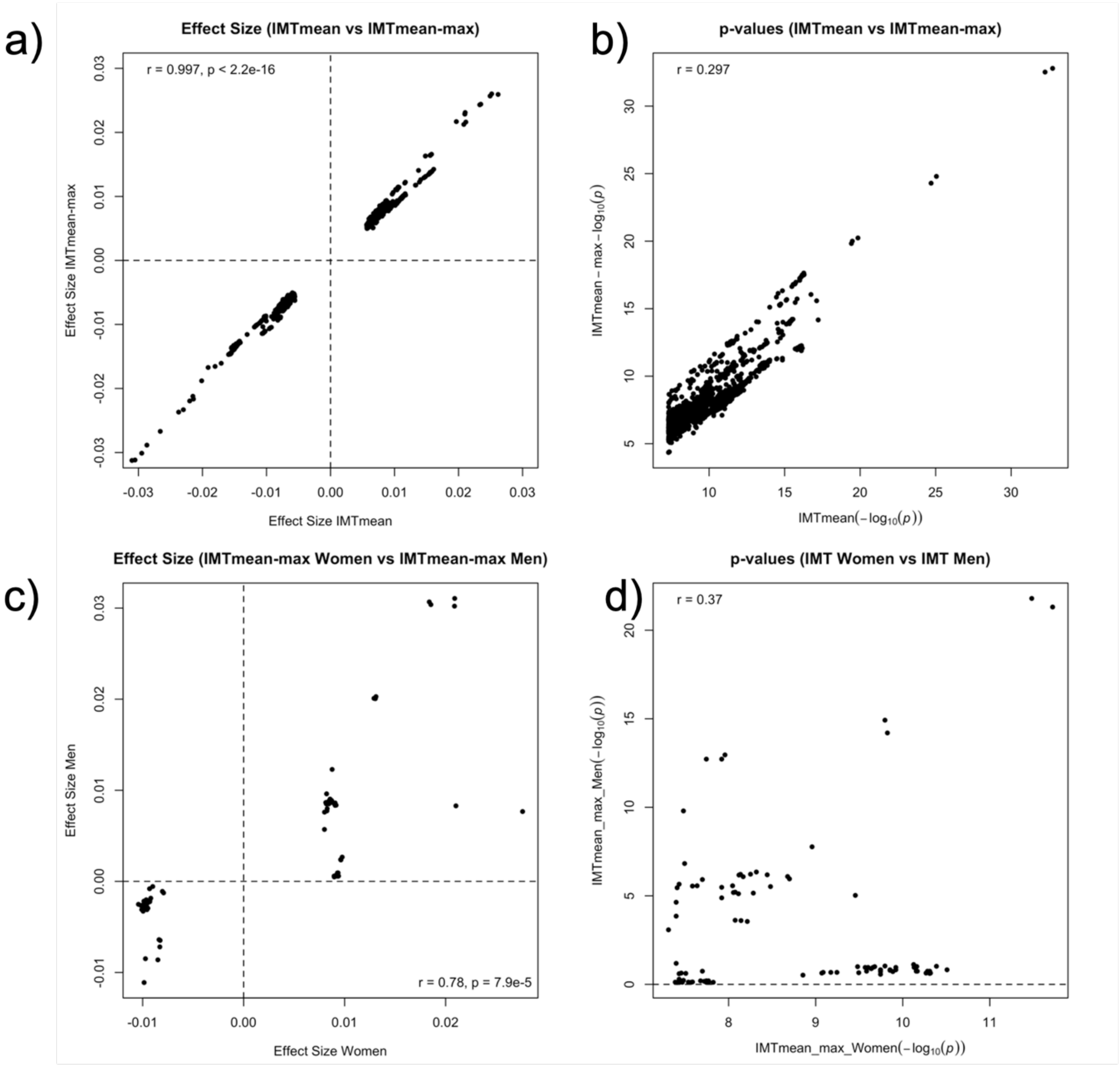

**Supplementary Figure 9**. Bar plot showing the number of SNPs in 95% credible sets at 39 loci identified in the European-only cIMT GWAS versus multi-ancestry cIMT GWAS. The x- and y-axis show 39 loci, and SNPs in the 95% credible sets, respectively. Eur, data from the European-only cIMT GWAS; Multi-Ethnic, data from the cIMT multi-ethnic GWAS.

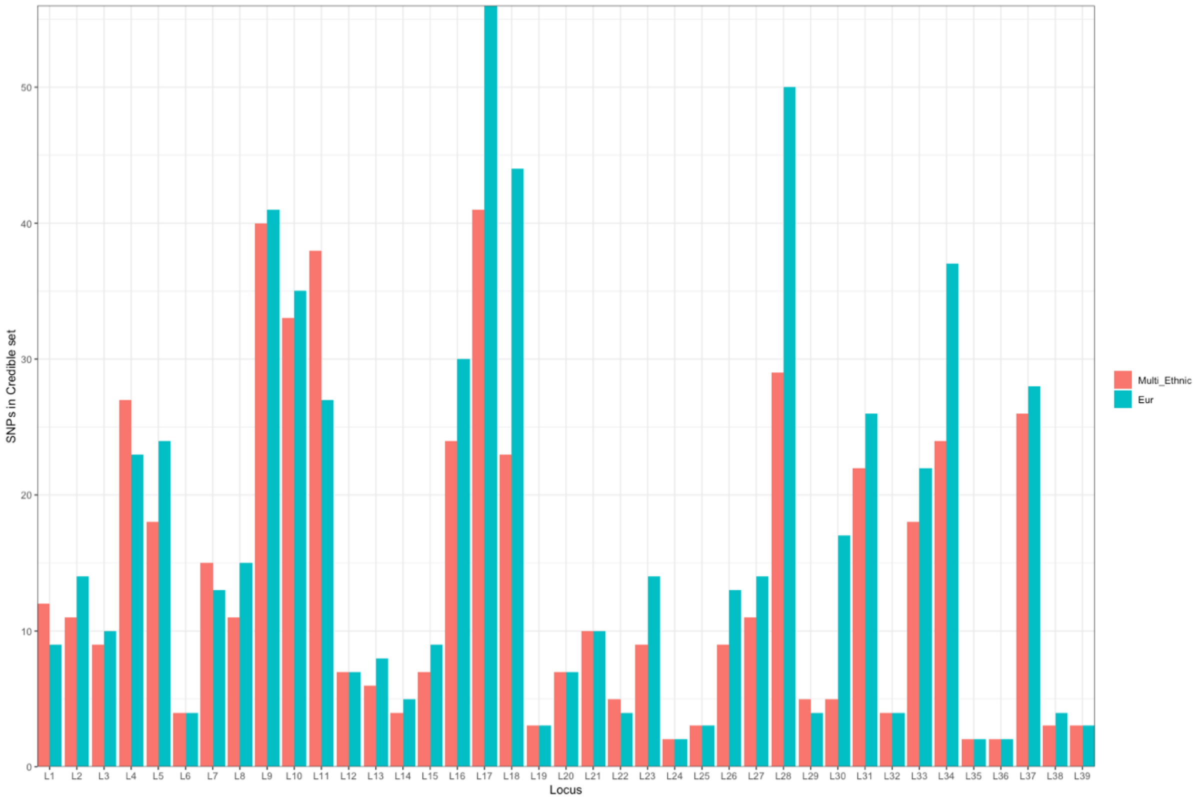
