## Supplementary Text for "Genome-wide association study and multi-ancestry meta-analysis identify common variants associated with carotid artery intima-media thickness"

**Study Descriptions and Methodology**


The Trans-Omics for Precision Medicine (TOPMed) program ……………………….…….. 2-15

The Africa Wits-INDEPTH partnership for Genomic Studies (AWI-Gen) …………………….. 15-17

The Mexican-American Coronary Artery Disease (MACAD) and the Hypertension-Insulin Resistance Family Study (HTN-IR) ………………………………………………………………… 17-20

The Baependi Heart Study (BHS) ……………………………………………...……......... 20-21

Athero-Express Biobank Study …………………….…………………………………...…. 21-24

Acknowledgements ………………………………………………………………………..25-29

References…………………………………………………………………………………..30-32

### Study Specific Descriptions and Methodology

#### Trans-Omics for Precision Medicine (TOPMed) program

**Introduction:** We leveraged whole genome sequencing (WGS) data from the from the Trans-Omics for Precision Medicine (TOPMed) program to perform a traditional GWAS as well as aggregate analyses to identify both common and rare loci in a multi-ethnic cohort. A total of 24,953 individuals from 9 different studies and 4 different ancestry groups were available for analyses.

**Methods:**

**Whole Genome Sequencing:** Within TOPMed, WGS was conducted at a mean depth of >30X using Illumina HiSeq X Ten instruments at five sequencing centers. Variant discovery and genotype calling for freeze 8 were conducted jointly across the 9 discovery studies, as well as additional studies not included in our current analysis, using the GotCloud pipeline by the TOPMed Informatics Research Center. Quality control of genetic variants performed by the TOPMed Information Resource Center consisted of the removal of variants failing the support vector machine filter, with excess heterozygosity or Mendelian inconsistencies, or overlapping centromeric or other low complexity regions[1]. Quality control of samples performed by the TOPMed Data Coordinating Center consisted of the removal of duplicate samples pertaining to the same individual, samples with discrepancies between genetic and reported sex, samples with discrepancies between genetically inferred and reported pedigrees, and samples with poor quality based on concordance of WGS and genotyping array data.

**Quantification of Carotid Intima Media Thickness:** Carotid intima-media thickness (IMT) was determined from carotid ultrasound measurements. Two primary measures of IMT were presented. The first measure, IMT-mean,mean, was calculed by taking the mean of the mean left common carotid artery far wall and the mean right common carotid artery far wall. The second measure, IMT-mean,max, was calculated by taking the mean for four measurements of the common carotid, the maximum values from left near and far wall and the maximum values from right near and far wall. Phenotype harmonization was performed by the TOPMed Data Coordinating Center[2]. Further detail regarding the measurement of CAC in the included studies can be found in the Supplementary Methods.

**TopMED Single Variant and Aggregate Analyses**

**Single Variant Association Tests:** Genome-wide tests for single variant association with ln(IMT-mean,mean) and ln(IMT-mean,max) were performed using linear mixed models. The first step of this procedure was to fit the 'null model' under the null hypothesis of no individual genetic variant associations (i.e. without any individual genotype terms in the model). Fixed effect covariates in the null model included age at IMT measurement, sex, study, and the first eleven PC-AiR[3] principal components (PCs) of genetic ancestry. A 4th degree sparse empirical kinship matrix computed with PC-Relate[4] was included to account for genetic relatedness among participants. We also allowed for heterogeneous residual variances across combined study by race/ethnicity groups, as it has previously been shown that this can improve control of genomic inflation[5]. To improve power and control of false positives with a non-normally distributed phenotype, we implemented a fully-adjusted two-stage procedure for rank-normalization when fitting the null model. First, we fit a linear mixed model using ln(IMT-mean,mean) and ln(IMT-mean,max) as the outcome, with the fixed effect covariates, sparse kinship matrix, and heterogeneous residual variance model as described above. We then perform a rank-based inverse-normal transformation of the marginal residuals, and subsequently re-scale by their original variance. This re-scaling allows for clearer interpretation of estimated genotype effect sizes from the association tests. Second, we fit a second linear mixed model using the rank-normalized and re-scaled residuals as the outcome, with the same fixed effect covariates, sparse kinship matrix, and heterogeneous residual variance model used in the first stage. The output of this second null model was used to perform genome-wide score tests of genetic association for all individual genetic variants with minor allele count≥50. All association testing analyses were performed using the GENESIS software and assumed an additive genetic model.[6]

**Gene-based Aggregate Rare Variant Association Tests:** Multi-variant association tests were also performed genome-wide to assess the cumulative association of rare variants in gene-centric aggregation units based on the GENCODE v28 gene models[7] with ln(IMT-mean,mean) and ln(IMT-mean,max). Three annotation-based aggregation and filtering strategies were implemented. The first strategy focused primarily on variants in protein-coding regions and included variants which were high-confidence loss-of-function variants according to Ensembl Variant Effect Predictor[8]. The second strategy included the above variants along with deleterious missense variants as predicted by SIFT 4G[9], PolyPhen-2_HumDIV, PolyPhen-2_HumVar[10], LRT_pred, or inframe insertions, inframe deletions with fathmm_XF_coding_score>0.5.[11].The third and final strategy included variants from the first two strategies as well as synonymous variants with fathmm_XF_coding score >0.5 and regulatory variants either proximal to a gene or predicted to be regulating the gene. Specifically, we retained variants within the upstream 5 Kb region (putative promoters) of a gene or in GeneHancers[12] (putative enhancers) which were labelled by Ensembl regulatory build annotation[13] as promoters, promoter flanking regions, enhancers, CTCF binding sites, transcription factor binding sites, or open chromatin regions.

The annotation based variant filtering and gene-centric aggregation was performed using a local MySQL database built from annotations generated by the Whole Genome Sequence Annotator version v0.[14] and formatted using WGSAParsr version 6.3.8. After performing annotation-based aggregation and filtering, variants were further filtered to those that were non-monomorphic with MAF < 1% among study participants.The aggregate association testing was performed using the Efficient Variant-Set Mixed Model Association Test (SMMAT).[15] The SMMAT test used the same null model as was fit for the single variant association tests. For each aggregation unit, SMMAT efficiently combines a burden test P-value with an asymptotically independent adjusted SKAT test P-value using Fisher's method. This testing approach is more powerful than either a burden or SKAT test alone, and is computationally more efficient than the SKAT-O test.[16] Wu weights[17] based on the variant MAF were used to upweight rarer variants in the aggregation units.

Statistical significance was determined using a Bonferroni threshold of $2.2 \times 10^{-6}$ for both outcomes [ln(IMT-mean,mean) and ln(IMT-mean,max)] corresponding to the 22,272 aggregation units tested in the third and largest filtering strategy.

**Overview of TOPMed Studies:** Genetics of Cardiometabolic Health in the Amish (Amish): The Amish Complex Disease Research Program includes a set of large community-based studies focused largely on cardiometabolic health carried out in the Old Order Amish community of Lancaster County, Pennsylvania starting in 1995. The Amish cohort participating in the TOPMed Consortium comprises subjects 18 years and older from large multigenerational families who were recruited for specific protocols between 2001 and 2006.

**Carotid ultrasound measurement:** High-resolution B-mode ultrasound was carried out to image the right and left common carotid arteries. CIMT was measured between lumen intima and media-adventitia interfaces of the far wall of the common carotid arteries (the 1-cm segment proximal to the bifurcation) by a single reader using an automated edge detection system.

*Selection criteria for sequencing:* All Amish subjects were utilized in single and aggregate analyses.

**Atherosclerosis Risk in Communities (ARIC):** The ARIC study is a population-based prospective cohort study of cardiovascular disease sponsored by the National Heart, Lung, and Blood Institute (NHLBI). ARIC included 15,792 individuals, predominantly European American and African American, aged 45-64 years at baseline (1987-89), chosen by probability sampling from four US communities. Cohort members completed three additional triennial follow-up examinations, a fifth exam in 2011-2013, a sixth exam in 2016-2017, and a seventh exam in 2018-2019. The ARIC study has been described in detail previously.[18]

**Carotid ultrasound measurement:** As described previously,[19] a Biosound 2000IISA system was used to perform ultrasound imaging of the carotid artery at the baseline examination. Images were processed centrally by trained readers at the ARIC Ultrasound Reading Center. cIMT of the distal common carotid (1 cm proximal to dilation of the carotid bulb) was assessed using 11 attempted measurements of the far wall (in 1 mm increments). Trained readers adjudicated plaque presence or absence if 2 of the following 3 criteria were met: abnormal wall thickness (defined as cIMT >1.5mm), abnormal shape, and abnormal wall texture.[19]

**Selection criteria for sequencing:** ARIC participants were selected for whole genome sequencing as part of the TOPMed venous thromboembolism (VTE) and AFGEN projects. Additional ARIC participants were selected for whole genome sequenced as a part of the National Human Genome Research Institute (NHGRI) Center for Common Disease Genomics (CCDG) program. Genotypes of the ARIC participants sequenced through CCDG were jointly called with TOPMed.

ARIC participants of European Ancestry were previously included in CHARGE analyses. Aggregate analyses included all ARIC participants. Single variant analysis was restricted to individuals of African Ancestry.

**Coronary Artery Risk Development in Young Adult (CARDIA):** CARDIA is a multicenter, prospective study of longitudinal cardiovascular risk development in Black and White participants recruited as young adults. In 1985-1986, a total of 5,115 CARDIA Black and White participants 18–30 years of age were recruited from Birmingham, AL; Chicago, IL; Minneapolis, MN; and Oakland, CA for the baseline visit. Study participants were recruited to include approximately equal numbers by race (52% Black), sex (54% women), education, and age at each of the four field centers. The recruitment strategy and study objectives have been described in detail.[20] Participants were re-examined in the clinic after 2, 5, 7, 10, 15, 20, 25, and 30 years, with retention rates of 91%, 86%, 81%, 79%, 74%, 72%, 72%, and 71%, respectively.

**Carotid ultrasound measurements:** Carotid cIMT was measured at CARDIA Y20 in 3258 (92%) of participants using a standard protocol across all centers previously described in detail.[21] Images were acquired at end-diastole using a GE-Logiq-700 (Issaquoah, Illinois) equipped with a high-resolution M12L transducer at 13MHz frequency for the common carotid artery and 9 MHz frequency for the bifurcation and internal carotid arteries. Certified sonographers selected the image with the lowest arterial diameter and saved images on super VHS-videotape. On both the left and right sides, two images were acquired in the common carotid, just below the bifurcation, in the carotid bulb, and in the proximal 2 cm of the internal carotid artery. The first image was taken at an angle of 45 degrees to the horizontal and the second was at more a vertical angle of 20–25 degrees.

Certified readers digitized images on an image analysis workstation. Readers traced lumen-intima and media-adventitia interfaces over a 1 cm of artery using a Wacom imaging tablet for each carotid artery segment. IMT was calculated for the far and near walls on each image. Plaques were included as part of the intima-media and the extent of any stenoses. The mean-maximum wall thickness for each artery segment was defined as the mean of the mean near and far wall thickness for each of the images on the left and right side, thus IMT was derived from 4 measures of the common segment and 8 measures each for the bulb and internal segments. Pearson correlation coefficients based on 58 replicate studies were 0.86 for the common segment, 0.72 for the bulb and 0.88 for the internal carotid.

**Selection criteria for sequencing:** CARDIA participants who consented to participate in GWAS studies, had available DNA samples, and cIMT measures were included in the study.

CARDIA participants of European Ancestry were previously included in CHARGE analyses. Aggregate analyses included all CARDIA participants. Single variant analysis was restricted to individuals of African Ancestry.

**Cardiovascular Health Study (CHS):** The Cardiovascular Health Study (CHS) is a population-based cohort study initiated by the National Heart, Lung and Blood Institute (NHLBI) in 1987 to determine the risk factors for development and progression of cardiovascular disease (CVD) in older adults, with an emphasis on subclinical measures. The study recruited adults aged 65 or older at entry in four U.S.

communities (Sacramento, CA; Hagerstown, MD; Winston-Salem, NC; Pittsburgh, PA). and conducted extensive annual clinical exams between 1989-1999 along with semi-annual phone calls, events adjudication, and subsequent data analyses and publications. Additional data are collected by studies ancillary to CHS. In June 1990, four Field Centers Blood samples were drawn from all participants at their baseline examination and during follow-up clinic visits and DNA was subsequently extracted from available samples. CHS analyses were limited to participants with available DNA who consented to genetic studies. The baseline examinations consisted of a home interview and a clinic examination that assessed not only traditional risk factors but also measures of subclinical disease, including carotid ultrasound, echocardiography, electrocardiography, and pulmonary function.

**Carotid ultrasound measurements:** Carotid high-resolution B-mode ultrasonography was used to assess the average maximal thickness of the common and internal carotid arteries.[22] The maximal intimal–medial thickness of the common carotid artery and of the internal carotid artery was defined as the mean of the maximal intimal–medial thickness of the near and far wall on both the left and right sides. The number of measurements that were available for averaging ranged from 1 to 4 for the common carotid artery and 1 to 12 for the internal carotid artery.[23]

**Selection criteria for sequencing:** TOPMed CHS participants were included if they had appropriate consent, available DNA, and met any of the following criteria: (a) had an adjudicated idiopathic VTE event during follow-up, (b) were African American, (c) had an incident MI or definite fatal coronary heart disease (CHD) event during follow-up, (d) had a probable fatal CHD event during follow-up, (e) had an incident stroke during follow-up, (f) had a prevalent MI or stroke at baseline, or (g) were part of a random sample of "healthy elderly" participants who survived free of an MI or stroke.

Selection of participants proceeded in a hierarchical manner from criterion (a) to (g) without replacement. Each participant is thus assigned to one, and only one of the seven groups.

CARDIA participants of European ancestry were previously included in CHARGE analyses. Aggregate analyses included all CARDIA participants. Single variant analysis was restricted to individuals of African Ancestry.

**Diabetes Heart Study (DHS):** The Diabetes Heart Study (DHS) family of studies is novel in its focus on CVD in a type 2 diabetes (T2D) enriched population. The DHS objectives were to study the genetic and epidemiological origins of CVD in families affected with T2D. DHS recruited and phenotyped individuals from European American and African American families with multiple T2D-affected members. Ascertainment of families was based on at least two siblings concordant for T2D (defined as a clinical diagnosis of diabetes after the age of 34 years, in the absence of historical evidence of diabetic ketoacidosis). Unaffected siblings, similar in age to the siblings with T2D, were also invited to participate, as were any additional diabetes-affected siblings. African Americans made up 15.4% of the original participants. Recruiting was based upon family structure with no inclusions/exclusions based on prevalent CVD at the time of recruitment. The only individuals excluded were those with serious health conditions, e.g., advanced nephropathy (prior serum creatinine concentration >2 mg/dL) or active malignancy. Participants with prior heart attack or stroke were included. Thus, DHS represents a cross section of the T2D population. This study was approved by the Institutional Review Board of the Wake Forest University School of Medicine (Winston-Salem, NC). All participants provided written informed consent.

The African American-Diabetes Heart Study (AA-DHS) started after DHS and enrolled unrelated African Americans. AA-DHS objectives are to improve understanding of ethnic differences in coronary artery calcification (CAC) and calcified atherosclerotic plaque (CP) in populations of African and European ancestry. T2D in African Americans was diagnosed after the age of 30 years in the absence of diabetic ketoacidosis. Individuals who underwent prior coronary artery bypass surgery or coronary artery angioplasty and/or stent placement were not included in the analyses, because CAC scores could have been impacted by the procedures. Those with prior myocardial infarction (MI) or stroke were included. The only individuals excluded were those with serious health conditions, e.g., advanced nephropathy (prior serum creatinine concentration $\geq$2 mg/dL) or active malignancy. The study was approved by the WFSM Institutional Review Board, and all participants provided written informed consent.

**Carotid ultrasound measurements:** In DHS, high-resolution B-mode carotid ultrasonography was performed using a 7.5-MHz transducer and a Biosound Esaote (AU5, Indiannapolis, Ind) machine. Scans were performed of the near and far walls of the distal 10-mm portion of the common carotid artery at 5 predefined interrogation angles on each side.

*Selection criteria for sequencing*: TOPMed DHS participants were included if they had appropriate consent, available DNA, existing genetic data (e.g. GWAS, exome chip, linkage, etc.), measures of cIMT.

**Framingham Heart Study (FHS):** The Framingham Offspring cohort participants were recruited in 1971, and comprised the children of participants enrolled in the Original cohort and their spouses.[24] In 2002, the Framingham Third generation cohort was enrolled and comprised the children of the Offspring cohort participants (i.e., the grandchildren of the Original cohort participants).[25]

**Carotid ultrasound measurements:** Ultrasonographic images of the right and left common and internal carotid arteries were acquired using high-resolution B-mode ultrasound in the FHS Offspring cohort. Continual intima-media interface lines were manually traced by a certified reader Common carotid IMT was measured over a 1-centimeter segment located ~0.5 cm below the carotid-artery bulb that did contain any plaque. The internal carotid artery IMT was measured on a segment extending from the carotid bulb to 1 cm above the carotid sinus. IMT values were calculated based on the average of measurements on each side.[26]

**Selection criteria for sequencing:** FHS participants of European ancestry and with available DNA and informed consent contributed to whole genome sequencing; a subset of individuals with available measures of subclinical atherosclerosis contributed to the present investigation.

FHS participants were previously included in CHARGE analyses. These participants were included in TOPMed aggregate analyses and excluded from single variant analysis.

**Jackson Heart Study (JHS):**The JHS is a longitudinal investigation of genetic and environmental risk factors associated with cardiovascular disease in African Americans. The JHS represents an expansion of the Jackson Field Center of the Atherosclerosis Risk in Communities (ARIC) study to broaden data collection in an African American population and to increase access to and participation of African American populations and scientists in biomedical research and professions.[27] The JHS recruited 5306 African American residents living in the Jackson, Mississippi, metropolitan area of Hinds, Madison, and Rankin Counties.[28] The age at enrollment for the unrelated cohort was 35-84 years; the family cohort included related individuals >21 years old. Participants provided extensive medical and social history, had an array of physical and biochemical measurements and diagnostic procedures during a baseline examination (2000-2004) and two follow-up examinations (2005-2008 and 2009-2013).[29] Samples for genomic DNA were collected during the first two examinations.[30] Consent for genetic studies and broad sharing of genetic data was provided by 3,482 participants; after all quality control procedures, whole genome sequence data are available for 3,406 participants. Annual follow-up interviews and cohort surveillance for cardiovascular events and mortality are ongoing.[31]

**Carotid ultrasound measurements:** In the JHS, an electrocardiography-gated, B-mode, and spectral steered Doppler with an integrated recorder ultrasound machine was used to obtain the carotid artery images in a 7.5 MHz linear-array transducer at the baseline visit.[29] Images were obtained bilaterally at the far and near walls on three segments of the carotid artery: the common carotid artery (CCA), bifurcation of the carotid artery, and internal carotid artery. All segments were imaged from the optimal angle (the angle of interrogation that most clearly shows the separation of the internal and external carotid arteries and the tip of the flow divider). The observed values were obtained for each segment, side, and wall. For each segment, sequences of 150 consecutive frames over approximately five cardiac cycles were digitized. The widest diameter frame during systole was then selected for measurement based on visualization of arterial interfaces. Maximum likelihood estimates were calculated by adjusting for missing data in the collecting, processing, and reading of carotid images. CCA intima media thickness (CIMT) represented a maximum likelihood estimate of the average values across the right and left CCA far wall.

**Selection criteria for sequencing:** All JHS participants were included with CIMT data and who consented for genetic studies were sequenced through TOPMed.

**Multi-Ethnic Study of Atherosclerosis (MESA):** The MESA contribution to TOPMed consisted of the parent MESA study, as well as the MESA Family Study. The MESA is a study of the characteristics of subclinical cardiovascular disease (disease detected non-invasively before it has produced clinical signs and symptoms) and the risk factors that predict progression to clinically overt cardiovascular disease or progression of the subclinical disease. MESA researchers study a diverse, population-based sample of 6,814 asymptomatic men and women aged 45-84. Thirty-eight percent of the recruited participants are white, 28 percent African-American, 22 percent Hispanic, and 12 percent of Chinese descent.

Participants were recruited from six field centers across the United States: Wake Forest University, Columbia University, Johns Hopkins University, University of Minnesota, Northwestern University and University of California - Los Angeles. DNA has been extracted and lymphocytes cryopreserved (for possible immortalization) for study of candidate genes and possibly, genome-wide scanning, expression, and other genetic techniques. Participants are being followed for identification and characterization of cardiovascular disease events, including acute myocardial infarction and other forms of coronary heart disease (CHD), stroke, and congestive heart failure; for cardiovascular disease interventions; and for mortality.

In addition to the six Field Centers, MESA involves a Coordinating Center, a Central Laboratory, and Central Reading Centers for Computed Tomography (CT), Magnetic Resonance Imaging (MRI), Ultrasound, and Electrocardiography (ECG). Protocol development, staff training, and pilot testing were performed in the first 18 months of the study. The first examination took place over two years, from July 2000 - July 2002. It was followed by six examination periods that were 17-20 months in length. Participants have been contacted every 9 to 12 months throughout the study to assess clinical morbidity and mortality.

**MESA Family Study**, an ancillary study to MESA funded by a grant from NHLBI, is to apply modern genetic analysis and genotyping methodologies to delineate the genetic determinants of early atherosclerosis within families in two large non-majority US populations, i.e. African Americans and Hispanic Americans. This is being accomplished by utilizing all the current organizational structures of the Multi-Ethnic Study of Atherosclerosis (MESA) and Genetic Centers at Cedars-Sinai Medical Center and University of Virginia.

**Carotid ultrasound measurements:** Trained technicians in each field center performed B-mode ultrasonography of the right and left near and far walls of the internal carotid and common carotid arteries.[32] They used the Logiq 700 ultrasound device (General Electric Medical Systems, Waukesha, Wisconsin) to record images. An ultrasound reading center (Department of Radiology, Tufts–New England Medical Center, Boston, Massachusetts) measured maximal IMT of the internal and common carotid sites as the mean of the maximum IMT of the near and far walls of the right and left sides. In addition, for this manuscript, we created a composite *z* score for overall maximal IMT by summing the values of the 2 carotid IMT sites (if both were measured) after standardization (subtraction of the mean and division by the SD of each measure) and then dividing by the SD of the sum. If only 1 of the 2 measures was available, it was used. The resulting variable, hereafter referred to as *z* score maximum IMT, has a mean of 0 and an SD of 1. Each participant and his or her physicians were notified whether an accompanying Doppler assessment suggested significant carotid stenosis (≥50%), but no recommendation was made about treatment.

**Selection criteria for sequencing:** MESA classic participants who were consented to MESA Genetics participation, have sufficient DNA volume in central lab repository (≥ 40 mcg DNA), and have maximum relevant phenotype data available.

Whole genome sequencing was completed on 4,619 MESA Classic participants

MESA Family Ancillary study also provided DNA samples for TOPMed WGS via the study/designation of "AACAC" (African-American Coronary Artery Calcium consortium study). Sample selection in this case was defined as having consented to MESA Family Ancillary Study (which required genetics consent), not already sequenced as a MESA participant, and having self-reported Black or African-American race.

**San Antonio Family Study (SAFS):** The SAFS results from the amalgamation of two San Antonio-based genetic studies. The first is the longitudinal San Antonio Family Heart Study which began in 1991 and was designed to primarily investigate the genetics of cardiovascular disease and its risk factors in Mexican Americans. The SAFHS included 1,431 individuals in 42 extended families at baseline.[33] With some additional recruiting, it has now been expanded to 1,662 individuals in 47 families. Ascertainment occurred by way of the random selection of an adult Mexican American proband, without regard to presence or absence of disease and almost exclusively from Mexican American census tracts in San Antonio. The second component study is the San Antonio Family Gall Bladder Study (Dr. Duggirala, PI) which included 907 individuals from 39 families ascertained similarly to the SAFHS but with the requirement that the original proband also be diabetic.[34] This is a very weak form of ascertainment in Mexican Americans, where lifetime prevalence of diabetes approaches 30%. In fact, 20 years after the initiation of the SAFS, the prevalence for major diseases such as heart disease, diabetes, and obesity are not significantly different between these two component studies. Finally, we have expanded these pedigrees in recent years by examining a set of 498 children than are part of these families. This expansion was part of Dr. Duggirala’s San Antonio Family Assessment of Metabolic Risk Factors in Youth (SAFARI) study. Additionally, we have seen 112 of these children subsequently as adults. Combined, these studies have 3,099 individuals primarily from 73 families. Our study is a mixed longitudinal design. Subjects have been seen between 1 and 4 times with an average of 1.95 examinations.

**Carotid ultrasound measurements:** For this study, analysis was limited to 772 individuals in 29 families who participated in the second visit of the San Antonio Family Heart Study, occurring between 1996 and 1999 and whom had B-mode ultrasound information available.[33] IMT measurements has been previously described.[35] In brief, B-mode ultrasound evaluations were completed on bilateral segments of extracranial carotid arteries applying protocols identical to those in the Cardiovascular Health Study.[32] Ultrasound images were recorded on super-VHS tapes and sent monthly to central carotid ultrasound readers, where one reader reads each participant's image. The scanning protocol required sonographers to obtain, on the right and left sides, one lateral view of the common carotid artery and three views of the internal carotid artery in different fixed planes. The common carotid artery was defined as the 10-mm segment of the carotid artery immediately proximal to the origin of the bulb, where the near and far walls of the artery were parallel. Three views of internal carotid artery segments were centered on the site of maximum wall thickness within the carotid artery bulb or where the sonographer considered the artery to be normal (without evidence of plaque) on the initial 10mm of the arterial segment. Pulsed-wave Doppler was recorded at the point of maximum velocity in each carotid artery. Ultrasound readers measured the near and far wall IMT of common carotid view and three different views (near and far wall, plus carotid arterial plaque) of the internal carotid artery centered on sites of maximum thickness.[36] Measurements between the near and far walls for both the common and internal carotid arteries were also averaged to provide a composite measure of each artery.

**Selection criteria for sequencing:** All 3,099 SAFS individuals underwent whole genome sequencing.

**The Africa Wits-INDEPTH partnership for Genomic Studies (AWI-Gen)**

The Africa Wits-INDEPTH partnership for Genomic Studies (AWI-Gen) is a National Institutes of Health (NIH) funded Collaborative Centre of the Human Heredity and Health in Africa (H3Africa) Consortium investigating the genomic and environmental risk factors for cardiometabolic diseases in Africans. The study was cross-sectional in nature and was investigating populations from six sub-Saharan African sites in four countries (Burkina Faso, Ghana, Kenya and South Africa) in East, West and Southern Africa[37]. Study participants provided informed consent and were enrolled for data collection from 2012 to 2016. Over 12 000 sub-Saharan Africans were enrolled form rural and urban settings and aged from 40 to 60 years with a parity between male and female individuals[37]. All participants completed a questionnaire with questions on demography, health history and behaviour. Anthropometric measurements were taken and fasting venous blood samples were collected for genotyping (H3Africa array) and phenotyping (biomarkers) purposes as described in the AWI-Gen resource paper and the detailed AWI-Gen methods paper[37,38]. Ultrasound scans were performed to assess the right and left common carotid intima-media thickness (cIMT).

cIMT was measured from Dual B-mode ultrasound images of the carotid tree as a typical double line of the arterial wall. The far wall of both the left and right common carotid artery were imaged using a linear-array 12L-RS transducer with a GE Healthcare B-mode LOGIQe ultrasound machine (GE, Healthcare, CT, USA). The participant was in a supine position for the measurements, head turned towards the left at a 45-degree angle to measure the right carotid. Operators used anatomical landmarks to identify the common carotid artery (CCA) on a longitudinal plane and the image was frozen. The operator then identified a continuous 1cm segment (10 mm) of the CCA far wall. The operator then placed a cursor between two points (10 mm apart) on this segment with the proximal starting point 1cm from the bulb of the CCA. The inbuilt software then automatically detected the intima-lumen and the media-adventitia interfaces and calculated the minimum, maximum and mean common cIMT in mm and to three decimal places. To measure the left carotid, the participant's head was turned to the opposite side, and the process was repeated. Mannheim Consensus criteria, for use of cIMT as continuous variable in population-based studies, were applied to QC the data. Mean cIMT was defined as the average of the mean right cIMT and mean left cIMT measurements. Mean Max cIMT was calculated as the average of the maximum cIMT from the left and right and used for the GWAS analysis.

The H3Africa genotyping array, designed as an African-common-variant-enriched GWAS array (Illumina) with ~2.3 million SNPs (https://www.h3abionet.org/h3africa-chip), was used to genotype genomic DNA using the Illumina FastTrack Service. The following pre-imputation QC steps were applied to the entire AWI-Gen genotype data set. Individuals with a missing SNP calling rate greater than 0.05 were removed. SNPs with a genotype missingness greater than 0.05, MAF less than 0.01 and failing Hardy-Weinberg equilibrium (HWE) at a P-value less than 0.0001 were removed. Non-autosomal and mitochondrial SNPs, and ambiguous SNPs that did not match the GRCh37 references alleles or strands were also removed. Imputation was performed on the cleaned dataset (with 1,729,661 SNPs and 10,903 individuals) using the Sanger Imputation Server and the African Genome Resource as reference panel. EAGLE2 [39] was used for pre-phasing and the default PBWT algorithm was used for imputation. After imputation, poorly imputed SNPs with info scores less than 0.6, MAF less 0.01, and HWE P-value less than 0.00001 were excluded. The final QC-ed imputed data had 13.98 M SNPs, and only participants with both good quality cIMT and genotyping data (n = 7895) were used for the GWAS analyses. Association was tested using fastGWA with adjustment for age, sex, site and 4 PC [40]. A GRM for relatedness was built with a subsample of 1014126 SNPs without in low LD with each other. The option --fastGWA-mlm-exact was used to perform an exact MLM-based association analysis without the GRAMMAR-GAMMA approximation, and run the analysis was run using the automated Nextflow pipeline h3abionet/h3gwas [41].

The AWI-Gen study was approved by the Wits Human Research Ethics Committee (Medical) under clearance number is M121026, and this sub-study under clearance number M1706110. All enrolled participants signed informed consent and study procedures were compliant with the Declaration of Helsinki.

### **<u>The Mexican-American Coronary Artery Disease (MACAD) and the Hypertension-Insulin Resistance Family Study (HTN-IR)</u>**

#### **Study Description**

**MACAD:** The Mexican-American Coronary Artery Disease (MACAD) Study was designed to examine the genetic basis of coronary artery disease and insulin resistance using a family-based design [42]. Family members of Mexican-American probands with documented coronary artery disease were recruited from the Los Angeles area. The GUARDIAN Consortium genotyping included 772 individuals from 208 families.

**HTN-IR:** The Hypertension-Insulin Resistance Family Study (HTN-IR) was designed to examine the genetic basis of hypertension and insulin resistance using a family-based design [43]. Family members of Mexican American probands with documented hypertension were recruited from the Los Angeles area. The GUARDIAN Consortium genotyping included 708 individuals from 156 families.

**Carotid ultrasound measurements**: Ultrasound image acquisition and carotid artery intima-media thickness (CIMT) measurement were conducted with standardized procedures and technology developed specifically for longitudinal measurement of atherosclerosis (Patents 2005, 2006, 2011) (Hodis HN, Mack WJ, Henderson VW, Shoupe D, Budoff MJ, Hwang-Levine J, Li Y, Feng M, Dustin L, Kono N, Stanczyk FZ, Selzer RH, Azen SP for the ELITE Research Group. Vascular effects of early versus late postmenopausal treatment with estradiol. N Engl J Med 2016;374:1221–1231.). Briefly, using a Toshiba SSH-140A ultrasound system with a linear array 7.5 MHz transducer, high-resolution B-mode ultrasound images of the jugular vein stacked above the carotid artery were transversely and longitudinally acquired along with internal anatomical landmarks for reproducible transducer angulation. The baseline carotid artery image for each individual was used as an online guide for longitudinal image acquisition. All instrumentation settings, including depth of field, gain, ultrasound input power, dynamic range and monitor intensity settings used at baseline were maintained for follow-up examinations establishing instrument setup standardization encompassing the full dynamic range of the ultrasound echo across all examinations within the same individual. Ultrasound images and single lead electrocardiogram (ECG) were simultaneously recorded. These standardized procedures result in reproducible imaging and processing of the same portion of the arterial wall at each examination necessary for accurately tracking atherosclerosis change. Using automated computerized edge detection software and sequential frame averaging, far wall

CIMT was measured at sub-pixel resolution (Patents 2005, 2006, 2011) (PMID: 7840805) (PMID: 11137099). Just proximal to the carotid artery bulb along a 1-cm length at the same point of the cardiac cycle, CIMT was determined as the average of 70 to 100 measurements between the intima-lumen and media-adventitia interfaces. This method standardizes the location and distance during the same cardiac cycle over which CIMT is measured, ensuring comparability within and across participants. This CIMT method is correlated with the change in coronary artery disease assessed by quantitative coronary angiography (PMID: 10856529) and is predictive of clinical coronary events (PMID: 9471928). The coefficient of variation of repeated CIMT measurements is typically <3% and often approaches 1%.

**GWAS Genotyping:** All samples were genotyped on the Illumina HumanOmniExpress BeadChip, and alleles were called using GenomeStudio software (Illumina, San Diego, CA) [44 Shen, 2005 #11]. Samples with call rates >0.98, single nucleotide polymorphisms (SNPs) with call rates >0.99, and minor allele frequency (MAF) >0.001 passed laboratory quality control, with 22,000 additional SNPs manually reviewed for clustering accuracy. Samples were removed from analysis if the overall call rate was <0.98, if the samples were genetic outliers for sex and admixture proportions, if the samples were monomorphic, or if there was inconsistent fingerprinting from existing SNP data [45]. The primary inferential SNPs did not exhibit differential missingness by trait, had a SNP call rate >98%, and did not depart from Hardy- Weinberg equilibrium expectations. Pedigrees were examined for consistency of stated family structure. Each SNP was examined for Mendelian inconsistencies using PedCheck (Program for Detecting Marker Typing Incompatibilities in Pedigree Data, http://watson.hgen.pitt.edu/register/docs/pedcheck.html), and inconsistencies were converted to missing. Population substructure was estimated using ADMIXTURE version 1.21 (http://www.genetics.ucla.edu/software/admixture) based on SNPs that passed quality control. Data from the HapMap Project, including Mexican ancestry, were used as reference populations [45]. Admixture proportions were included as covariates in the tests of association with liver enzymes.

**Acknowledgement:** This research was supported by the GUARDIAN Study DK-085175 from the National Institute of Diabetes and Digestive and Kidney Diseases (NIDDK) and from the National Institutes of

Health, National Heart, Lung, and Blood Institute in collaboration with the Mexican-American Coronary Artery Disease Project (MACAD) HL-088457, and the Hypertension in Insulin Resistance (HTN-IR) study HL-0697974, as well as DK-079888.

### The Baependi Heart Study (BHS)

The Baependi Heart Study is an epidemiological study in Baependi, a city in a rural area (752 Km2, 18,307 inhabitants at the 2010 census) located in Minas Gerais State, Brazil (21.95°S, 44.88°W). The initial data collection phase occurred between December 2005/January 2006, and one hundred and nine families were selected, corresponding to 1,627 individuals of both genders. In 2010 during the first follow-up visit, 2,239 individuals from the same families participated in the protocol. The current analysis used a cross-sectional analysis of data collected at the second evaluation visit (from 2010 to 2015) on subjects that underwent carotid ultrasonography. Individuals that presented angina, infarct, cardiac insufficiency and revascularization were removed from our analysis. The technique used to measure and calculate cIMT was to measure a double line with the definition of the light-intima and media-adventitia interfaces of the vessel. The distance between the two acoustic interfaces was considered the cIMT measure. Measurements with reference to the light-intima and media-adventitia interfaces of the vessel were standardized using the Philips Envisor HD7 ultrasound equipment with a linear 7.5MHz transducer. Three measurements of the cIMT were performed on each side, starting 1.0 cm below the upper limit of the image (1.0 cm below the carotid bifurcation), using the posterior (distal) wall of the common carotid artery, with a 5 mm spacing between them. For each side, we performed the arithmetic mean of the measurements. cIMT was calculated by the mean of the three measurements performed on the walls of the distal carotid, using Osirix ™ software. In addition, carotid bifurcation was studied at 4.0 cm for plaques. Images of common carotids acquired and documented in a 4.0 cm length starting at the carotid bifurcation. When plaques occurred that did not allow the measurement, 1.0 cm below the upper limit of the image, the measurement was performed immediately after the plaque. The atheromatous plaque was defined as a focal structure that extends at least 0.5 mm to the vessel lumen or measures more than 50% of the adjacent cIMT measurement value or a measurement greater than 1.5 mm. BHS DNA samples for genotyped using Axiom_PMRA.r3 array (ThermoFisher) and genotypes annotated using the Axiom_PMRA.na35.annot.db provided at the ThermoFisher site. Genotype calling was performed using Affymetrix Power Tools. Initial VCF file contained 850483 variants that fulfilled all quality criteria. Imputation was performed using the Haplotype Reference Consortium Michigan Imputation Server using the TOPMED reference haplotype panel as reference. After imputation data were exported in the standard PLINK format and downstream QC procedures and statistical analysis were conducted using the latest PLINK (http://pngu.mgh.harvard.edu/_purcell/plink) and R software packages (http://www.r-project.org/), installed on a Linux based computation resource. After imputation markers were kept if R2 > 0.3, and minor allele frequency (MAF) > 0.01. A HWE p-value <1 × 10−20 was used to control for potential genotyping clustering problems.

**Athero-Express Biobank Study**

Patient population: Atherosclerotic plaques were obtained from patients undergoing a carotid endarterectomy (CEA) procedure and included in the Athero-Express Biobank Study (AE, www.atheroexpress.nl), an ongoing biobank study at the University Medical Centre Utrecht (Utrecht, The Netherlands) and the St. Antonius Hospital (Nieuwegein, The Netherlands) [46]. This study complies with the Declaration of Helsinki, and all participants provided informed consent. The medical ethical committees of the respective hospitals approved this study which was registered under number TME/C-01.18. The study design of the AE was described before[46], but in brief: during surgery blood and plaques are obtained, stored at -80°C and plaque material is routinely used for histological analysis[46].

**Histological phenotyping**

We described the standardized (immuno)histochemical analysis protocols used in the AE before[1,2]. In short, 10-micron cross-sections of the paraffin-embedded segments were cut using a microtome and examined under a microscope. We quantitatively scored the number of macrophages (CD68) and smooth muscle cells

(SMCs, α-actin), as percentage of the microscopy field area by computerized analysis usingAnalySIS 3.2 software (Soft Imaging Systems GmbH, Münster, Germany). Intraplaque vessel density (CD34) was assessed as the average number per 3 hotspots. Intraplaque hemorrhage (IPH) was scored as no/yes using a hematoxylin and eosin staining (HE). Intraplaque fat was defined as less or more than 40% fat per total plaque area using HE. The amount of calcification (using HE) and collagen (picrosirius red) were binary scored as no/minor vs. moderate/heavy staining. All histological observations were performed by the same dedicated technician and interobserver analyses have been reported previously[3].

**DNA isolation, genotyping, and imputation**

**DNA isolation and genotyping**

We genotyped the AE in three separate, but consecutive experiments. In short, DNA was extracted from EDTA blood or (when no blood was available) plaque samples of 1,858 consecutive patients from the Athero-Express Biobank Study and genotyped in 3 batches. For the Athero-Express Genomics Study 1 (AEGS1) 891 patients (602 males, 262 females, 27 unknown sex), included between 2002 and 2007, were genotyped (440,763 markers) using the Affymetrix Genome-Wide Human SNP Array 5.0 (SNP5) chip (Affymetrix Inc., Santa Clara, CA, USA) at Eurofins Genomics(https://www.eurofinsgenomics.eu/, formerly known as AROS). For the Athero-Express Genomics Study 2 (AEGS2) 954 patients (640 makes, 313 females, 1 unknown sex), included between 2002 and 2013, were genotyped (587,351 markers) using the Affymetrix Axiom® GW CEU 1 Array (AxM) at the Genome Analysis Center (https://www.helmholtz-muenchen.de). The two first batches, AEGS1 and AEGS2, were described before[4]. For the Athero-Express Genomics Study 3 (AEGS3) 658 patients (448 males, 203 females, 5 unknown sex), included between 2002 and 2016, were genotyped (693,931 markers) using the Illumina GSA MD v1 BeadArray (GSA) at Human Genomics Facility, HUGE-F (http://glimdna.org/index.html). All experiments were carried out according to OECD standards. We used the genotyping calling algorithms as advised by Affymetrix (AEGS1 and AEGS2) and Illumina (AEGS3): BRLMM-P, AxiomGT1, and Illumina GenomeStudio respectively.

**Quality control after genotyping**

After genotype calling, we adhered to community standard quality control and assurance (QCA) procedures of the genotype data from AEGS1, AEGS2, and AEGS3[4,5]. Samples with low average genotype calling and sex discrepancies (compared to the clinical data available) were excluded. The data was further filtered on 1) individual (sample) call rate > 97%, 2) SNP call rate > 97%, 3) minor allele frequencies (MAF) > 3%, 4) average heterozygosity rate ± 3.0 s.d., 5) relatedness (pi-hat > 0.20), 6) Hardy–Weinberg Equilibrium (HWE p < 1.0×10−3), and 7) Monomorphic SNPs (< 1.0×10−6). After QCA 2,493 samples remained, 108 of non-European descent/ancestry, and 156 related pairs. These comprise 890 samples and 407,712 SNPs in AEGS1, 869 samples and 534,508 SNPs in AEGS2, and 649,954 samples and 534,508 SNPs in AEGS3 remained.

**Imputation**

Before phasing using SHAPEIT2, data was lifted to genome build b37 using the liftOver tool from UCSC (https://genome.ucsc.edu/cgi-bin/hgLiftOver). Finally, data was imputed with 1000G phase 3, version 5 and HRC release 1.1 as a reference using the Michigan Imputation Server (https://imputationserver.sph.umich.edu/)[6]. These results were further integrated using QCTOOL v2, where HRC imputed variants are given precedence over 1000G phase 3 imputed variants.

**RNA isolation and single-cell RNA sequencing**

Atherosclerotic plaques were collected from 35 individuals and processed for single-cell RNA sequencing as described before [47]. In short, time between surgical removal and plaque processing did not exceed 10 minutes; note that the inclusion of a small medial layer in the dissected tissue could not be excluded during the surgical procedure. The remainder of the plaque was washed in RPMI and minced into small pieces with a razor blade. The tissue was then digested in RPMI 1640 containing 2.5 mg/mL Collagenase IV (ThermoFisher Scientific), 0.25 mg/mL DNAse I (Sigma), 2.5 mg/mL Human Albumin Fraction V (MP Biomedicals) and 1 mM Flavopiridol (Selleckchem) at 37oC for 30 minutes. Subsequently, the plaque cell suspension was filtered through a 70 μm cell strainer and washed with RPMI 1640. Cells were kept in

RPMI 1640 with 1% Fetal Calf Serum until subsequent staining for fluorescence-activated cell sorting. Remaining, unstained cells were cryostored in liquid nitrogen.

We adapted a CELseq2-protocol for single-cell RNA sequencing and processed the data as we described before [47]. Analyses were performed using Seurat (version 3.xx). Prior to processing, reads were filtered for mitochondrial and ribosomal genes, MALAT1, KCNQ1OT1, UGDH-AS1, and EEF1A. In order to omit doublets and low-quality cells, only cells expressing between 500 and 10.000 genes and genes expressed in at least 3 cells were used for further analysis. Data was log-normalized and scaled with the exclusion of unique molecular identifiers (UMIs). Top variable genes for all samples were used to combine samples into one object using Seurat function RunMultiCCA(), after which samples were aligned using AlignSubspace() with reduction.type=CCA and grouping.var=”plate”. Subsequently, canonical correlation analysis (CCA) reduction was performed with a resolution of 1.2 for 15 dimensions to identify clusters and to perform t-distributed stochastic neighbor embedding (tSNE). Cell types were assigned to cell clusters by evaluating gene expression of individual cell clusters using differential gene expression (Wilcoxon rank sum test) and analysis with SingleR8 against BLUEPRINT reference data.

### Acknowledgements


#### Study Specific Acknowledgements

Acknowledgements for each of the study cohorts follow below:

**CHARGE-UCLEB**: Infrastructure for the CHARGE Consortium is supported in part by the National Heart, Lung, and Blood Institute grant R01HL105756.

**TOPMed**

**Genetics of Cardiometabolic Health in the Amish (Amish):** The TOPMed component of the Amish Research Program was supported by National Institutes of Health (NIH) grants R01 HL121007, U01 HL072515, and R01 AG18728. The Amish studies upon which these data are based were supported by NIH grants R01 AG18728, U01 HL072515, R01 HL088119, R01 HL121007, and P30 DK072488. See publication: PMID: 18440328. WGS for "NHLBI TOPMed: Genetics of Cardiometabolic Health in the Amish" (phs000956) was performed at the Broad Institute of MIT and Harvard (HHSN268201500014C).

**Atherosclerosis Risk in Communities (ARIC):** The Atherosclerosis Risk in Communities study has been funded in whole or in part with Federal funds from the NHLBI, NIH, Department of Health and Human Services (contract numbers HHSN268201700001I, HHSN268201700002I, HHSN268201700003I, HHSN268201700004I and HHSN268201700005I). The authors thank the staff and participants of the ARIC study for their important contributions.

**Coronary Artery Risk Development in Young Adult (CARDIA)**: The Coronary Artery Risk Development in Young Adults Study (CARDIA) is conducted and supported by the NHLBI in collaboration with the University of Alabama at Birmingham (HHSN268201800005I & HHSN268201800007I), Northwestern University (HHSN268201800003I), University of Minnesota (HHSN268201800006I), and Kaiser Foundation Research Institute (HHSN268201800004I). The Y25 cardiac CT scans were supported by an NHLBI award to Vanderbilt University Medical Center (R01-HL098445). This manuscript has been reviewed by CARDIA for scientific content.

**Cardiovascular Health Study (CHS):** This research was supported by contracts HHSN268201200036C, HHSN268200800007C, HHSN268201800001C, N01HC55222, N01HC85079, N01HC85080, N01HC85081, N01HC85082, N01HC85083, N01HC85086, 75N92021D00006, and grants R01HL64587, U01HL080295, and U01HL130114 from the NHLBI, with additional contribution from the National Institute of Neurological Disorders and Stroke (NINDS). Additional support was provided by R01AG023629 from the National Institute on Aging (NIA). A full list of principal CHS investigators and institutions can be found at CHS-NHLBI.org. The content is solely the responsibility of the authors and does not necessarily represent the official views of the NIH.

**Diabetes Heart Study (DHS):** This work was supported by R01 HL92301, R01 HL67348, R01 NS058700, R01 AR48797, R01 DK071891, R01 AG058921, the General Clinical Research Center of the Wake Forest University School of Medicine (M01 RR07122, F32 HL085989), the American Diabetes Association, and a pilot grant from the Claude Pepper Older Americans Independence Center of Wake Forest University Health Sciences (P60 AG10484).

**Framingham Heart Study (FHS):** This project has been funded in whole or in part with Federal funds from the National Heart Lung and Blood Institute, National Institutes of Health, Department of Health and Human Services, under Contracts NO1-HC-25195, HHSN268201500001I and 75N92019D00031 and grant supplement R01 HL092577-06S1. We also acknowledge the dedication of the FHS study participants without whom this research would not be possible.

**Jackson Heart Study (JHS):** The Jackson Heart Study (JHS) is supported and conducted in collaboration with Jackson State University (HHSN268201800013I), Tougaloo College (HHSN268201800014I), the Mississippi State Department of Health (HHSN268201800015I) and the University of Mississippi Medical

Center (HHSN268201800010I, HHSN268201800011I and HHSN268201800012I) contracts from the NHLBI and the National Institute for Minority Health and Health Disparities (NIMHD). The authors also wish to thank the staff and participants of the JHS.

**Multi-Ethnic Study of Atherosclerosis (MESA):** MESA and the MESA SHARe projects are conducted and supported by the National Heart, Lung, and Blood Institute (NHLBI) in collaboration with MESA investigators. Support for MESA is provided by contracts 75N92020D00001, HHSN268201500003I, N01-HC-95159, 75N92020D00005, N01-HC-95160, 75N92020D00002, N01-HC-95161, 75N92020D00003, N01-HC-95162, 75N92020D00006, N01-HC-95163, 75N92020D00004, N01-HC-95164, 75N92020D00007, N01-HC-95165, N01-HC-95166, N01-HC-95167, N01-HC-95168, N01-HC-95169, UL1-TR-000040, UL1-TR-001079, UL1-TR-001420, UL1TR001881, DK063491, and R01HL105756. MESA Family is conducted and supported by the NHLBI in collaboration with MESA investigators. Support is provided by grants and contracts R01HL071051, R01HL071205, R01HL071250, R01HL071251, R01HL071258, R01HL071259, and by the National Center for Research Resources, Grant UL1RR033176.The authors thank the other investigators, the staff, and the participants of the MESA study for their valuable contributions. A full list of participating MESA investigators and institutes can be found at http://www.mesa-nhlbi.org.

**MCAD and HTN-IR**: Whole genome sequencing (WGS) for the Trans-Omics in Precision Medicine (TOPMed) program was supported by the National Heart, Lung and Blood Institute (NHLBI). WGS for "NHLBI TOPMed: Multi-Ethnic Study of Atherosclerosis (MESA)" (phs001416.v1.p1) was performed at the Broad Institute of MIT and Harvard (3U54HG003067-13S1). Centralized read mapping and genotype calling, along with variant quality metrics and filtering were provided by the TOPMed Informatics Research Center (3R01HL-117626-02S1, contract HHSN268201800002I). Phenotype harmonization, data management, sample-identity QC, and general study coordination, were provided by the TOPMed Data Coordinating Center (3R01HL-120393; U01HL-120393; contract HHSN268180001I). MESA and the MESA SHARe projects are conducted and supported by the National Heart, Lung, and Blood Institute

(NHLBI) in collaboration with MESA investigators. MESA and the MESA SHARe projects are conducted and supported by the National Heart, Lung, and Blood Institute (NHLBI) in collaboration with MESA investigators. Support for MESA is provided by contracts 75N92020D00001, HHSN268201500003I, N01-HC-95159, 75N92020D00005, N01-HC-95160, 75N92020D00002, N01-HC-95161, 75N92020D00003, N01-HC-95162, 75N92020D00006, N01-HC-95163, 75N92020D00004, N01-HC-95164, 75N92020D00007, N01-HC-95165, N01-HC-95166, N01-HC-95167, N01-HC-95168, N01-HC-95169, UL1-TR-000040, UL1-TR-001079, UL1-TR-001420. Funding for SHARe genotyping was provided by NHLBI Contract N02-HL-64278. Genotyping was performed at Affymetrix (Santa Clara, California, USA) and the Broad Institute of Harvard and MIT (Boston, Massachusetts, USA) using the Affymetrix Genome-Wide Human SNP Array 6.0. The provision of genotyping data was supported in part by the National Center for Advancing Translational Sciences, CTSI grant UL1TR001881, the National Institutes for Diabetes and Digestive and Kidney Diseases contract R01-HL151855-01 and contract R01HL146860, and the National Institute of Diabetes and Digestive and Kidney Disease Diabetes Research Center (DRC) grant DK063491 to the Southern California Diabetes Endocrinology Research Center. Infrastructure for the CHARGE Consortium is supported in part by the National Heart, Lung, and Blood Institute (NHLBI) grant R01HL105756. This research was also supported by the Mexican-American Coronary Artery Disease (MACAD) National Heart, Lung, and Blood Institute, contracts R01-HL088457, R01-HL-60030; Hypertension and Insulin Resistance (HTN-IR) contracts R01-HL067974, R01-HL-55005, R01-HL 067974, and contract HL-055798 (NIDDM-Athero).

**Athero-Express Biobank lookup**: Dr. Sander W. van der Laan is funded through grants from the Netherlands CardioVascular Research Initiative of the Netherlands Heart Foundation (CVON 2011/B019 and CVON 2017-20: Generating the best evidence-based pharmaceutical targets for atherosclerosis [GENIUS I&II]). We are thankful for the support of the ERA-CVD program 'druggable-MI-targets' (grant number: 01KL1802), the EU H2020 TO_AITION (grant number: 848146), and the Leducq Fondation 'PlaqOmics'.

**San Antonio Family Study (SAFS):** Collection of the San Antonio Family Study data was supported in part by National Institutes of Health (NIH) grants R01 HL045522, MH078143, MH078111 and MH083824; and whole genome sequencing of SAFS subjects was supported by U01 DK085524 and R01 HL113323. We are very grateful to the participants of the San Antonio Family Study for their continued involvement in our research programs.

**Million Veteran Program:** The MVP is funded by the Department of Veterans Affairs Office of Research and Development, Million Veteran Program Grant #MVP000. This publication does not represent the views of the Department of Veterans Affairs or the United States Government. MVP was also supported by three additional Department of Veterans Affairs awards (I01-01BX03340, I01-BX003362, and I01-CX001025).
